# Quantitative Trend Analysis of SARS-CoV-2 RNA in Municipal Wastewater Exemplified with Sewershed-Specific COVID-19 Clinical Case Counts

**DOI:** 10.1101/2022.03.13.22272304

**Authors:** Vince Pileggi, Jayson Shurgold, Jianxian Sun, Minqing Ivy Yang, Elizabeth Edwards, Hui Peng, Amir Tehrani, Kimberley Gilbride, Claire Oswald, Shinthuja Wijayasri, Dana Al-Bargash, Rebecca Stuart, Zeinab Khansari, Melanie Raby, Janis Thomas, Tim Fletcher, Albert Simhon

**Affiliations:** Ontario Ministry of the Environment, Conservation and Parks, Toronto, ON, Canada M4V 1M2; AMR Division, Public Health Agency of Canada, 130 Colonnade Rd, Ottawa, ON, Canada K1A 0K9; University of Toronto, 27 King’s College Cir, Toronto, ON, Canada M5S 3K1; Ryerson University, 350 Victoria St, Toronto, ON, Canada M5B 2K3; Toronto Public Health, 277 Victoria St, Toronto, ON, Canada M5B 1W2; Canadian Field Epidimiology Program, Public Health Agency of Canada, 100 Colonnade Rd, Ottawa, ON, Canada K2E 7ME

**Keywords:** SARS-CoV-2 RNA, wastewater surveillance, linear break-point trend-analysis, normalization, interpretation

## Abstract

We present and demonstrate a quantitative statistical linear trend analysis (*QTA*) approach to analyze and interpret SARS-CoV-2 RNA wastewater surveillance results concurrently with clinical case data. This demonstration is based on the work completed under the Ontario (Canada) Wastewater Surveillance Initiative (WSI) by two laboratories in four large sewersheds within the Toronto Public Health (TPH) jurisdiction. The sewersheds were sampled over a 9-month period and data were uploaded to the Ontario Wastewater Surveillance Data and Visualization Hub (*Ontario Dashboard*) along with clinical case counts, both on a sewershed-specific basis. The data from the last 5-months, representing a range of high and low cases, was used for this demonstration. The QTA was conducted on a sewershed specific approach using the recommendations for public health interpretation and use of wastewater surveillance data by the United States Centers for Disease Control and Prevention (US CDC). The interpretation of the QTA results was based on the integration of both clinical and wastewater virus signals using an integration matrix in an interim draft guide by the Public Health Agency of Canada (PHAC). The key steps in the QTA consisted of (i) the calculation of Pepper Mild Mottle Virus (PMMoV), flow and flow-PMMoV-normalized virus loads; (ii) computation of the linear trends including interval estimation to identify the key inflection points using a segmented linear regression method and (iii) integrated interpretations based on consideration of both the cases and wastewater signals, as well as end user actionability. This approach is considered a complementary tool to commonly used qualitative analyses of SARS-CoV-2 RNA in wastewater and is intended to directly support public health decisions using a systematic quantitative approach.

## 1. Introduction

As of August 1, 2021 the wastewater contributed by over 70% of the population of the province of Ontario (Canada) was surveilled for the presence of SARS-CoV2 RNA fragments in wastewater through the Ontario Wastewater Surveillance Initiative (WSI) led by the Ontario Ministry of the Environment, Conservation and Parks (MECP). A total of 86 centralized wastewater treatment plants (WWTP) within 34 Ontario public health units have been sampled up to five times per week, through a coordinated provincial effort. Since August 2020, 13 academic laboratories and one federal public health agency laboratory have conducted wastewater analyses by RT-qPCR. The data associated with the analysis of the wastewater samples has been centrally uploaded onto the Ontario Wastewater Surveillance Initiative Data and Visualization Hub (*Ontario Dashboard*). The data and visualization products are made available to participating public health units, municipalities and university laboratories. A review of recent relevant literature (below) provides the context and identifies a need for a quantitative integration and interpretation reporting for the combined clinical case data with wastewater surveillance trends.

The Water Research Foundation^®^ provided recommendations from a group of global experts following their April 2020 summit event, “International Water Research Summit on Environmental Surveillance for the Genetic Signal of SARS-CoV-2 in Sewersheds” on the use of wastewater surveillance for the tracking of COVID-19. Some of the key recommendations were related to communication of results to public health decision makers, the public and other key stakeholders. A key recommendation, categorized as ‘currently very feasible,’ was the general use of wastewater surveillance data to assess trends. These trends and changes in trends, could be used to determine occurrence and reemergence while tracking the impact of medical and social interventions using curves to designate an increase or decrease within a given sewershed. A second key recommendation, categorized as ‘currently may or may not be feasible’ was that the assessment of community prevalence or tracking disease prevalence in the community; identification of areas of concern, as well as areas that are not impacted by the virus; and estimation of the level of infection in a community. However, summit participants agreed that wastewater surveillance could offer valuable information to public health decision makers on the potential prevalence of community disease including: the identification of ‘hot spots’ within a community with appropriate spatial sampling approach; identifying areas that are not currently impacted by the virus and the occurrence of COVID-19 in specific confined sub-communities such as correctional facilities, long term care facilities or schools [1].

A recent review published by Naughton et al, 2021 [2] examined data from 195 countries of which 50 (as of March 8, 2021) had wastewater monitoring programs. The authors conclude that generally, the analysis of wastewater surveillance and wastewater-based epidemiology (WBE) data and analysis reported worldwide represents a qualitative assessment showing general trends with little in the way of rigorous quantification of the results needed to support public health decisions. Here we present a novel and rigorous *quantitative statistical linear trend analysis (QTA)* based on normalized concentrations of SARS-CoV-2 RNA found in municipal wastewater. Further, these wastewater trends are directly compared to the linear trends of sewershed specific COVID-19 cases by reported date provided by the Ontario Ministry of Health (MOH) for temporal comparison and decision making as described below.

Arabzadeh et al, 2021 [3] discussed the need for *data smoothing techniques* to address the issue of the intrinsic random fluctuations in the wastewater viral signal to reduce misleading conclusions on community prevalence. Some of the issues that contribute to the random fluctuations include dilution, fate and transport processes within the sewershed, variations in the number of persons discharging, excretion rates and differences in daily water consumption rates. Arabzadeh et al, 2021 [3] investigated 13 smoothing algorithms applied to the virus signals monitored in four WWTPs over an 8-month period in Austria and found that when considering various performance metrics, SPLINE, Generalized Additive Model and Friedman’s Super Smoothers provided superior methods and reduced the tendency to over-smoothing. Through a first analysis of these data sets they found an influence of catchment size for WBE as smaller communities reveal a “signal threshold before any relation with infection dynamics is visible and also a higher sensitivity towards infection clusters.” To address some of these issues related to wastewater signal variability associated with flow and populations dynamics, biomarker and flow normalization have been identified as useful methods [3].

A modeling framework applied to three Canadian cities, including Toronto’s four main sewershed (also considered in our case studies), is described in Nourbakhsh et al, 2021 [4]. It provides a detailed mechanistic approach to assess both the transmission of the SARS-CoV-2 at the population level and the associated RNA gene fragments within the wastewater after fecal shedding by infected individuals in the community. The integration and interpretation of both metrics concurrently, *clinical case counts* (CCC) data and *wastewater signals* (WVS), was implemented in the modeling framework. Some of the key factors included in the proposed model was the *transit time* and *decay rate* of the RNA within the sewer system and it was assessed that with a transit time of less than 3-days the decay rate of the wastewater SARS-CoV-2 signals (e.g., samples collected at the influent to the WWTP) is not likely affected and likely remains associated with fecal contributions of the virus from the community within the sewer system [4].

The work by D’Aoust et al, 2021 [5] from Ontario, Canada, examined the relationship between SARS-CoV-2 viral signals from wastewater samples collected from an upstream pumping station and from an access port at a downstream wastewater treatment lagoon with the community’s COVID-19 percent test positivity in a small, rural community in Ontario. They found that, samples collected at the upstream pumping station, particularly when normalized for fecal contributions using PMMoV, predicted an increase in SARS-CoV-2 viral signal in the wastewater approximately 10-14 days prior to an increase in community’s COVID-19 reported test percent positivity. This was in contrast to samples collected at lagoons which revealed weak SARS-CoV-2 viral signal in wastewater. Based on their findings they concluded that sampling at upstream sites of lagoons, at locations such as pumping stations, is preferred over downstream lagoon sites and that PMMoV normalized signal, compared to unnormalized signals, was more significantly correlated to new community COVID-19 cases.

Earlier work by D’Aoust et al, [6] focused on the Robert O. Pickard Environmental Centre, serving the city of Ottawa, ON, Canada, with a population of approximately 1 million residents. The sewershed is reported to have a sewage travel time in the range of 2 – 35 hours and mean of 12 hours (h) to the WWTP with a mean daily flow of 435 megaliters per day. A number of key observations relating to the value of wastewater-based surveillance were reported including: (1) the SARS-CoV-2 viral signal in primary clarified sludge was found to increase 48 h prior to increase in new COVID cases; (2) SARS-CoV-2 wastewater viral signal increased 96 h prior to increase in COVID-19 hospitalizations; (3) time-lagged correlations confirmed that increased wastewater signal predates new COVID cases and hospitalizations and (4) wastewater-based surveillance may be a better testing approach detecting surges in COVID-19 cases and providing a valuable complementary public health metric to clinical information.

Ai et al, 2021 [7] from the state of Ohio in the USA investigated nine WWTPs in central Ohio and considered normalization methods with PMMoV and Cross-Assembly phage (crAssphage) and also considered if wastewater surveillance can serve as a sentinel piece for detecting SARS-CoV-2 variants of concern within a community. SARS-CoV-2 RNA concentrations in wastewater and COVID-19 cases were found to correlate well when they were imputed using a 5-day moving day average. Normalization using the mean concentration in the community was investigated along with linear and polynomial models to improve correlations. Also, the sequencing results from wastewater samples showed agreement with the sequencing results from clinical nasal swab samples during an early period of new strain emergence. It was concluded that wastewater surveillance is ideal for fast tracking variant emergence and transmission within a community.

An additional metric introduced by Xiao et al, 2021 [8] which when combined with clinical case data trends may provide additional insight into the potential for under- or over-estimation of disease incidence in the community is the wastewater viral signal (WVS) to clinical case count (CCC) ratio or WCR. It was reported that an increase in the WCR would imply an increase in asymptomatic cases in the community; a decrease in the WC ratio would imply clinical testing may be over-estimating disease incidence when counting previously infected cases as new cases and a baseline or no change in the WC ratio, would imply there is sufficient public health capacity. Additionally it was recommended that the magnitude and range of the WC ratio be established for each sewershed to assess a population baseline where sufficient public health testing capacity indicated a relatively constant WC ratio. Furthermore the WCR was recommended as a useful metric to better detect short-term trends of asymptomatic disease transmission compared to the wastewater and clinical data independently [8].

Despite these significant advances in demonstrating a strong relationship between wastewater-based surveillance data and clinical cases for COVID there remains a gap in quantitative approaches to interpret these trends collectively. Here we report and demonstrate the utility of a novel approach for quantifying and interpreting sewershed-specific trends of both wastewater viral RNA signals (WVS) and clinical case counts (CCC) with Toronto as a case study.

## 2. Methods

### 2.1. Quantitative Statistical Linear Trend Analysis (QTA)

The QTA methodology is based on an implementation of the recommendations on public health interpretation and use of wastewater surveillance data from the United States Centers for Disease Control and Prevention (US CDC) [9] and the recommendations on the integration of clinical and wastewater surveillance signals in the interim draft guide by the Public Health Agency of Canada (PHAC) [10,11]. The following key steps were applied:

a. *Calculation of the normalized virus loads* per US CDC recommendations [9].
b. *Computation of the linear trends* as per US CDC recommendations [9] and using break-point linear regression [12].
c. *Interpretations* based on PHAC draft integration matrix and guidance [10,11].

The data used in this report was downloaded from the secure Ontario Dashboard. Two academic laboratories in their analysis of 24-hour wastewater composite field samples from the influent to WWTPs (post screening and prior to degritting) followed prescribed laboratory quality assessment and quality control (QA/QC) measures and field recommended practices referenced and described in the MECP documents [13,14]. The generated primary data included viral RNA quantified using RT-PCR with the N1 and N2 primers as well as PMMoV. Each lab collected and analyzed samples 3–5 days a week during the testing period. Key metadata included the mean daily flowrate over the 24-hour sample period provided by Toronto Water. This information was documented on the *Ontario Data Template* (ODT) (the Supplement describes the ODT) based on the data structure of the *Open Data Model (ODM) for Wastewater-Based Surveillance* [15,16], a framework for wastewater surveillance, before being uploaded to the Ontario Dashboard. An additional MECP document provides details about laboratory protocols and is intended to address the issue of dealing with ‘trace’ concentration levels or values measured at or below the method limit of detection (LOD) [17]. In our approach we considered all technical and biological replicate values as reported for integration into arithmetic mean values. Further, if the arithmetic mean average resulted in a zero-value, this was replaced by one-half of the ‘UJ’ value [17] within the reported data set. This final data adjustment was done for wastewater data only and to eliminate numerical issues associated with log-transformation of zero-values prior to break-point linear regression.

The case studies for this work focused on four large urban municipal sewersheds described in Table 1 and included the Ashbridges Bay (TAB), the Humber River (THR), the Highland Creek (THC) and the North Toronto (TNT) WWTP sewersheds. Some of the sewershed details have been provided in the published Toronto Water annual reports dedicated to each WWTP [18–21].

**Table 1.** Municipal sanitary sewershed system general information

| Sewershed ID | Collection System<sup>a,b</sup> | Population Served<sup>c</sup> | Area<sup>d</sup> (ha) | Median Flow<sup>e</sup> (CI) (ML/d) | Travel Time<sup>f</sup> (h) |
| --- | --- | --- | --- | --- | --- |
| TAB | Combined | 1,603,700 | 25,000 | 476 (464 - 484) | 10 |
| THR | Combined | 685,000 | 18,000 | 234 (231 - 239) | 7 |
| THC | Separated | 533,000 | 10,600 | 164 (160 - 165) | 5 |
| TNT | Combined | 55,000 | 5,000 | 18 (17 - 18) | 2 |

<sup>a</sup> Combined, means the sewershed includes some older 1-pipe sections that collect both sewage and stormwater with a higher degree of dilution during storm events <sup>b</sup> Separated, means the sewershed is predominantly a 2-pipe system; one collecting sewage and the other stormwater <sup>c</sup> Population, refers to the 2020 census <sup>d</sup> Area (ha), is an estimate of the extent of the sewershed in hectares <sup>e</sup> Median Flow (CI) (ML/d), means the median 5-month flow with 95 % confidence interval calculated using the adjusted bootstrap percentile (BCa) interval type (n = 82 - 117) <sup>f</sup> Travel Time (h) is the estimated mean wastewater travel time in hours taking into account the furthest distance in the sewershed to the WWTP and an assumed design scouring velocity for the sewers of 1 m/s.

The overall workflow in the QTA process is given in Figure 1 with a description of the data sets used and the related sample R-code provided in the Supplement. Details of the steps used in the QTA are described in the following sections.

**Figure 1.**
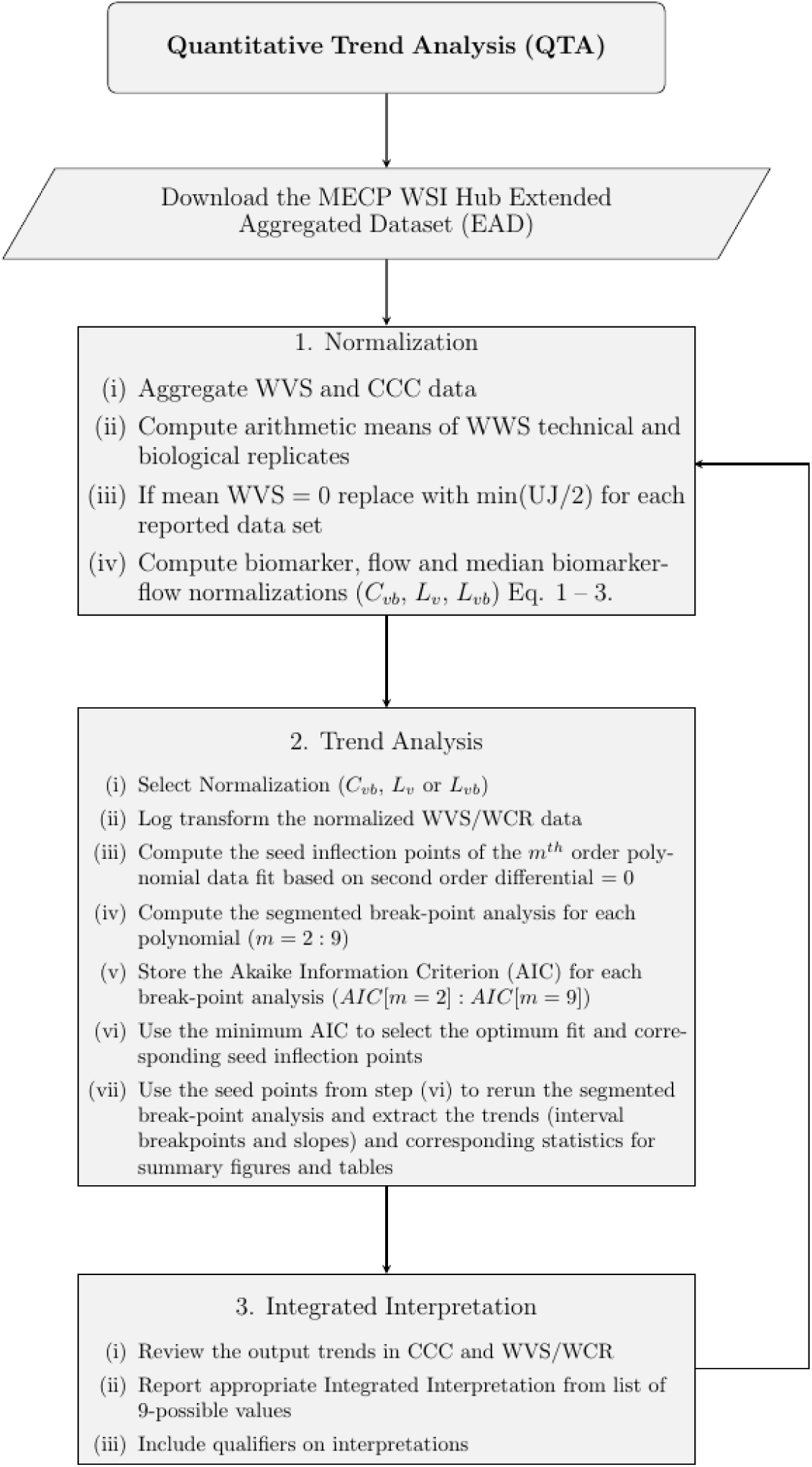
Quantitative trend analysis (QTA) flowchart showing the three major steps in the QTA process: Normalization, Trend Analysis and Integrated Interpretation with automated optimum inflection point selection (Note that segmented() refers to the main R-function used from the *segmented* R-package).

#### 2.1.1. Normalization

Three different normalizations of the wastewater SARS-CoV-2 RNA signal were considered and included: PMMoV normalized concentration with the 2020 census population per 10^5^ introduced as a factor *C*_*vb*_ in 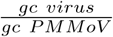 per 10^5^ population (Eq. 1); flow normalization viral flux *L*_*v*_ in 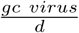 per 10^5^ population (Eq. 2) and the median biomarker-flow normalized flux *L*_*vb*_ in 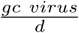 per 10^5^ population (Eq. 3):

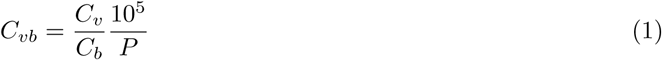

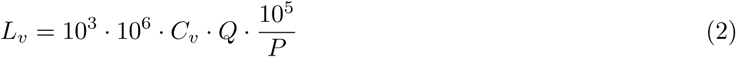

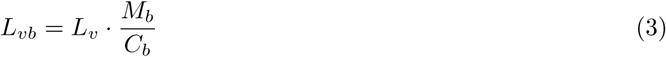

where:

i. *C*_*v*_ is the virus RNA gene concentration in gene copies per ml 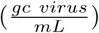.
ii. *C*_*b*_ is the fecal biomarker concentration in gene copies per ml 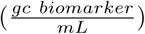 which takes into account the temporal variation of the population (*P*) in the sewershed [3].
iii. *Q* is the mean daily flowrate in megaliters per day 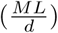 corresponding to the 24-hour composite sample collected of the influent wastewater at the wastewater sewage treatment plant (WWTP).
iv. *P* is the contributing sewershed 2020 population (*cap*).
v. *M*_*b*_ is the median biomarker gene counts per mL of 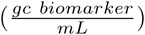 in the wastewater throughout the population during the complete sampling campaign [3].
vi. The 10^3^ is included for the unit conversion of *L* to *mL*.
vii. The 10^6^ is included for the unit conversion of ML to *L*.
viii. The 10^5^ is included to adjust the value per 10^5^ population (*cap*).

Eqs. 1, 2 and 3 apply equally for any virus RNA gene target including N1, N2 or N1N2 (average of N1 and N2). Further N1 and N2 represent the mean of technical replicates.

The virus biomarker normalized concentration *C*_*vb*_ in Eq. 1 may be interpreted as the virus concentration per fecal contribution from 10^5^ people within the specific sewershed being monitored. The viral flow normalized loading *L*_*v*_ in Eq. 2 may be interpreted as the daily virus load per flow contribution from 10^5^ people within the specific sewershed being monitored. The median biomarker-flow normalized loading *L*_*vb*_ in Eq. 3 may be interpreted as the total mass of viral RNA transported to the WWTP during the 24-h sampling period per contribution from 10^5^ people relative to the median PMMoV (*M*_*b*_) population contribution.

#### 2.1.2. Trend Analysis

The *computation of the linear trends* was based on a break-point linear analysis [12] of the arithmetic means of reported technical or biological replicates. The first step involved selecting the normalization mode (*C*_*vb*_, *L*_*v*_ or *L*_*vb*_) followed by an assessment of the ND reported values and log transformation of the wastewater data. Mean values of zero were replaced by the minimum UJ [17] value divided by 2 before log transforming the data. The case data was not transformed and a linear scale was maintained. An iterative automated process was used to find the optimum break-points and trends. This involved the following steps with sample R-code provided in the Supplemental: Function 1: (i) Fitting the complete data set to a liner regression model as the feed for the segmented routine (given a regression model, the segmented routine ‘updates’ it by adding one or more piece-wise linear relationships [12]); (2) Fitting the data sets (case data and log transformed wastewater data) to an *m*^*th*^ order polynomial (*m* = 2 − 9), iteratively, to find seed inflection points based on second derivative equal to zero; (iii) Applying the segmented routine to compute the break-point linear trend results; (iv) Computing the Akaike information criterion (AIC) for each break-point trend results for each polynomial fit (*m* = 2 − 9); (v) Using the minimum AIC as the criteria to select the optimum polynomial order and corresponding inflection seed points; and (vi) Selecting the optimum inflection points (minimum AIC) to rerun the segmented break-point analysis and extract the trends (break-points and interval slopes) with corresponding statistics for summary figures and tables.

The *segmented* R-package employs internal hypothesis testing about the existence of breakpoints and the use of sequential hypothesis testing to select the optimum number of breakpoints from the seed values. Various appropriate statistical methods are also integrated into the *segmented* routine to assess the confidence intervals and error margins associated with the break-points and linear trends [12].

The above analysis was applied to each sewershed specific data set and the results of the *segmented* routine, were extracted to determine the break-points identifying the dates of major changes in either the clinical case counts (CCC) or the wastewater signal (WVS) which were used to determine the *time intervals* and *duration* between significant changes. The *slope* of each segmented linear trend line was determined and associated with the slope standard errors (SE) and 95% confidence intervals (CI). Because the wastewater signal was log-transformed, the percent daily change (*PDC*) was computed per US CDC [9] using Eq. 4:

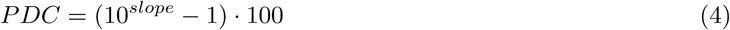

where:

i. *PDC* is the percent daily change (%) in wastewater signal.
ii. *slope* is the *log*_10_ transformed data regression line segment slope.

Based on the completed trend results customized figures and tables were generated to clearly identify the *time intervals* between transitions in clinical case counts and wastewater signals. The figures provide an estimate of the 95% confidence interval (CI) for the WVS (blue) and the CCC (red) about the best-fit linear trend line for each time interval. The error associated with the break-point was provided by the output from the *segmented* routine based on the the use of the smoothed score-like statistic and score-based interval estimators [12]. To identify the values corresponding to the values below the reported LOD values these points were color-coded green to distinguish them from other point values with reported values above the LOD. Summary tables are used to report the results with corresponding error analysis results. The tables include the time interval duration (start and end dates) with the corresponding daily changes in cases and the PDC in the normalized wastewater signal with the corresponding SE and 95 % CI. Color coded icons are added to facilitate quick identification directions 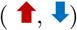 or no change 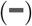 associated with the signals for each break point segment.

#### 2.1.3. Integrated Interpretations

The *interpretations* for each time interval were determined based on a comparison and integration of trend directions for both the case and wastewater signals using PHAC led collaborative recommendations derived from an *interpretation integration matrix* [10]. A total of 9 potential permutations were considered with each of the clinical case counts and wastewater signal having three potential outcomes for each slope or trend: **increasing, decreasing** or **baseline**. Table 2 provides the general interpretations of the integrated signals. Note that baseline is taken to mean that clinical case counts (CCC) or the wastewater signal (WVS) is near zero or at the limit of quantification, respectively. Additionally, general interpretations are provided to enable local public health epidemiology, recognizing that each scenario requires additional context that may be influencing either signal.

**Table 2.** General interpretation of integrated CCC and WVS trends

| Signal Trends |  | Trends Interpretation |
| --- | --- | --- |
| CCC | WVS | Integration of Clinical Case Counts (CCC) and Wastewater Signal (WVS) |
| ↑ | ↑ | **Both** the CCC and WVS are **trending up**. Strong evidence of escalating community transmission. |
| ↑ | ↓ | The CCC **trending up** and the WVS is **trending down**. Weak evidence of escalating community transmission. |
| ↑ | — | The CCC is **trending up** and the WVS is **baseline**. Some evidence of escalating community transmission. |
| ↓ | ↑ | The CCC is **trending down** and the WVS is **trending up**. Weak evidence of escalating community transmission. |
| ↓ | ↓ | **Both** the CCC and WVS signals are **trending down**. No evidence of escalating community transmission. |
| ↓ | — | The CCC is **trending down** and the WVS signal is **baseline**. No evidence of escalating community transmission. |
| — | ↑ | The CCC is **baseline** and the WVS is **trending up**. Some evidence of escalating community transmission. |
| — | ↓ | The CCC is **baseline** and the WVS is **trending down**. No evidence of escalating community transmission. |
| — | — | **Both** the CCC and WVS signals are **baseline**. No evidence of escalating community transmission. |

For example, even a scenario with both the CCC and WVS trending up, could be a result of local factors (e.g., an outbreak among elderly residents at a long-term care facility combined with WWTP sewershed maintenance operations that results in increased sediments in the collected wastewater sample which would not provide any real evidence of increased community transmission). Further, now that more than 78% of the population aged 12 and older are fully vaccinated in Ontario, these interpretations might be more applicable to urban centers. However, there are still many caveats to consider. The QTA approach alone may not be informative and interpretations may be confounding without a fuller consideration of the local field conditions. For example: (i) There is currently no definitive understanding of how viral RNA shedding patterns differ for vaccinated and unvaccinated infections; (ii) There may be increased uncertainty of the true incidence of SARS-CoV-infection given an increase in the frequency of mild cases in vaccinated populations; (iii) The stratification of the case counts by vaccination status is not fully understood and may change as a function of waning immunity; and (iv) As mass clinical testing is discontinued, the clinical signal may increasingly rely on hospitalization data as the only remaining robust and high-quality source of clinical information. Further, would a spike in the wastewater signal and an increase in hospitalizations trigger investigations for variants of concern (VOC) and outbreak management decisions? [10,11].

The core of the interpretation message is that, in most cases, wastewater provides a source of high-quality information that is independent from clinical surveillance (and associated pit-falls) that ultimately allows epidemiologists to better interpret local data in order to make appropriate decisions. Essentially, WVS that are baseline or trending down contribute public health intelligence that there is less cause for concern, while WVS that are trending up signify increasing concern [10,11].

The wastewater to cases ratio (WCR) [8] was also considered and integrated as part of an extension to the base QTA report intended to demonstrate the versatility of the QTA methodology to integrate new metrics for additional trend analysis and insight. Table 3 provides an integration of the PHAC and WC recommended interpretations intended to supplement the CCC and WVS trend interpretations [10,11].

**Table 3.** General interpretation of integrated Clinical Case Counts (CCC) and Wastewater Signal (WVS)

| Signal Trends |  | Trends Interpretation |
| --- | --- | --- |
| CCC | WC | Integration of CCC and WC Ratio |
| ↑ | ↑ | **Both** the CCC and WC Ratio **are trending up**. Strong evidence of escalating community transmission and increase in asymptomatic cases in the community. |
| ↑ | ↓ | The CCC **trending up** and the WC Ratio is **trending down**. Weak evidence of escalating community transmission and clinical testing may be over-estimating disease incidence when counting previously infected cases as new cases. |
| ↑ | — | The CCC is **trending up** and the WC Ratio is **baseline**. Some evidence of escalating community transmission and there is sufficient public health capacity. |
| ↓ | ↑ | The CCC is **trending down** and the WC Ratio is **trending up**. Weak evidence of escalating community transmission and increase in asymptomatic cases in the community. |
| ↓ | ↓ | **Both** the CCC and WC Ratio signals are **trending down**. No evidence of escalating community transmission and clinical testing may be over-estimating disease incidence when counting previously infected cases as new cases. |
| ↓ | — | The CCC is **trending down** and the WC Ratio signal is **baseline**. No evidence of escalating community transmission and there is sufficient public health capacity. |
| — | ↑ | The CCC is **baseline** and the WC Ratio is **trending up**. Some evidence of escalating community transmission and increase in asymptomatic cases in the community. |
| — | ↓ | The CCC is **baseline** and the WC Ratio is **trending down**. No evidence of escalating community transmission and clinical testing may be over-estimating disease incidence when counting previously infected cases as new cases. |
| — | — | **Both** the CCC and WC Ratio signals are **baseline**. No evidence of escalating community transmission and there is sufficient public health capacity. |

#### 2.1.4. Reporting the QTA Findings

The QTA results were provided in a succinct 2-page report considered a template model report for different metrics under considerations and for direct use by public health officials. In addition to the trend analysis results the QTA report also includes a statistical error analysis of each break point and trend slope for the end user to better assess the confidence of the reported trends.

The results of the QTA were summarized in various Figures and Tables generated by R-code using *ggplot* [22] and *tidyverse* [23] functionality along with additional R-packages including: *lubridate* [24]; *reshape2* [25]; *scales* [26]; *gridExtra* [27]; *cowplot* [28] and *kowr* [29]. The complete analysis and typesetting were generated within the R-Studio IDE [30] Version 1.4.1106, using *rmarkdown* [31], *bookdown* [32] and integrating LATEX[33] typesetting with a *pdf* output option. The complete analysis and manuscript typesetting was integrated into a single *rmarkdown* with customized functions all based on the *literate programming* paradigm to promote reproducibility and open science through a *reproducible workflow* [34]. Additional material was provided in the Supplement Document (*Supplement*) generated by a separate *Rmarkdown-scripts* and referenced throughout this document.

## 3. Results and Discussion

Figure 2 shows the combined cases by reported date from February 11/21 to August 29/21 for the four sewersheds and provides the relative contribution of each sewershed to the total clinical case load. The reported date is a derived variable that best estimates when the disease was acquired, and refers to the earliest available date from: symptom onset (the first day that COVID-19 symptoms occurred), laboratory specimen collection date (positive result), or reported date.

**Figure 2.**
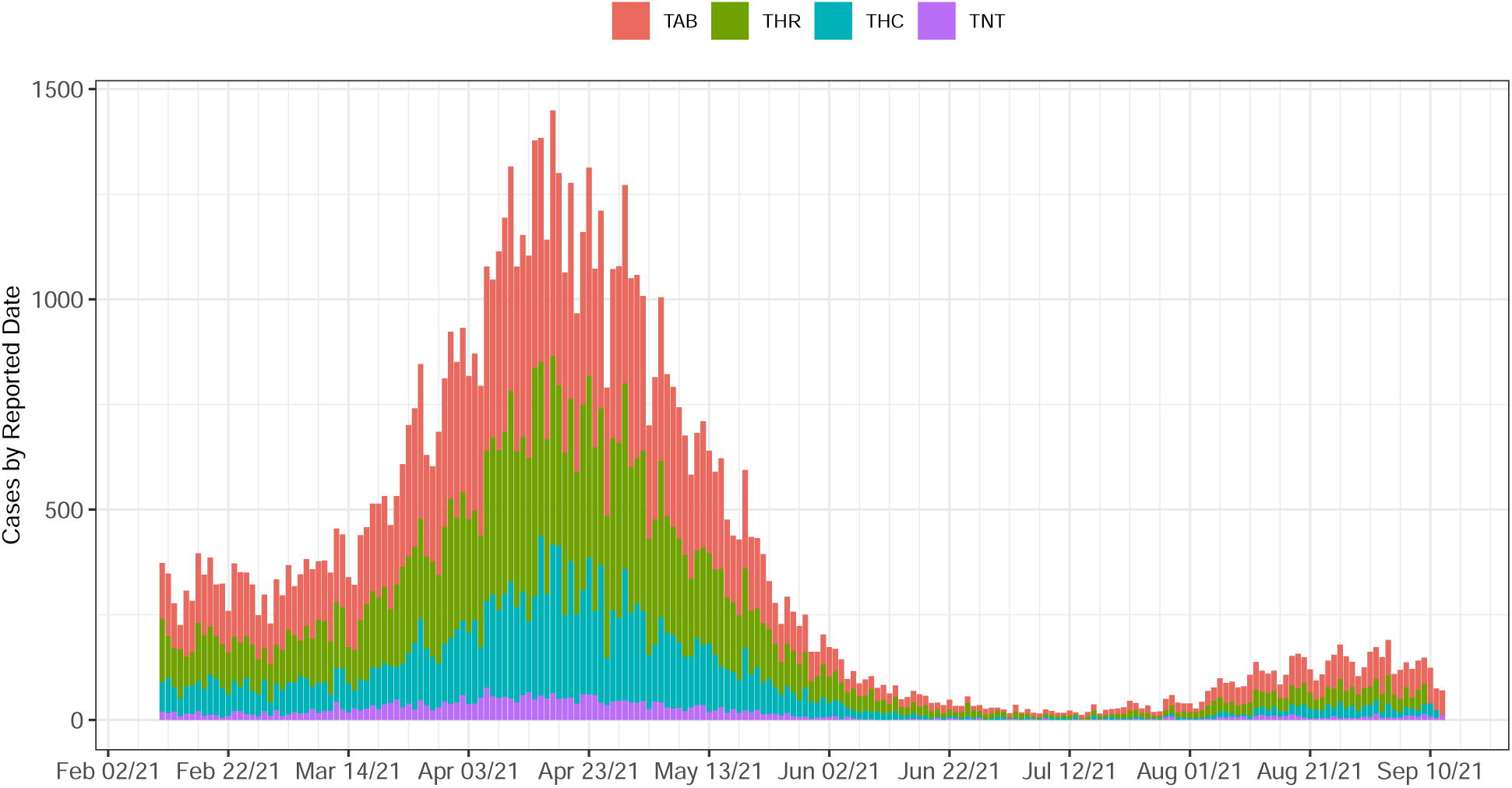
Combined cases by reported date for the TAB, THR, THC and TNT WWTPs sewersheds (February 11/21 – August 29/21)

The complete QTA process is demonstrated for different normalization approaches, parameters, metrics and selected sewersheds for both long and short-term trends. Some results are provided in the body for illustrative purposes while others are provided in the Supplement. All the results are considered in the results and discussion sections.

The QTA examples with various normalizations and case data are sewershed specific and interpretations of the trend results are based on the integration of clinical and wastewater surveillance signals [9–11] summarized in Table 2 and Table 3 which integrates the WC ratio [8]. Further interpretation of these results may require additional local expertise or tools to interpret results that appear to be unclear or contradictory. A high degree of coordination between jurisdictional experts (e.g., epidemiologists, wastewater operators and laboratory personnel) can assist with the identification of additional elements required for local interpretation (e.g., methodological problems) [8,10,11].

### 3.1. Ashbridges Bay Sewershed QTA

The TAB extended aggregated data set was downloaded from the Ontario Dashboard and used to generate the longer-term trends as part of the QTA under different normalization conditions (*C*_*vb*_, *L*_*v*_ and *L*_*vb*_). In this section we focus on the TAB sewershed which has an approximate area of 25,000 ha and a population of 1.6 million which is about 52 % of the population of the City of Toronto (Table 1). Here, we consider the different wastewater signal normalizations and how this influences the resulting trends and interpretations within the QTA process. The goal is to better understand the scope and limitations associated with the QTA process when considering a single sewershed under different normalization modes, parameters and metrics. Note that the detailed analysis of *L*_*v*_ and *L*_*vb*_ normalizations are provided in the Supplement but summarized and discussed here.

Figure 3 shows the Ashbridges Bay WWTP sewershed, overlay plot of the wastewater virus signal (WVS) for PMMoV normalized N1 viral viral concentration (*C*_*vb*_) (upper right panel) and the cases by reported date (lower right panel) as demonstrated on the Ontario Dashboard. The corresponding aggregated data sets were used to generate trends as part of the QTA under the different normalizing conditions (unnormalized, *C*_*vb*_, *L*_*v*_, *L*_*vb*_), respectively.

**Figure 3.**
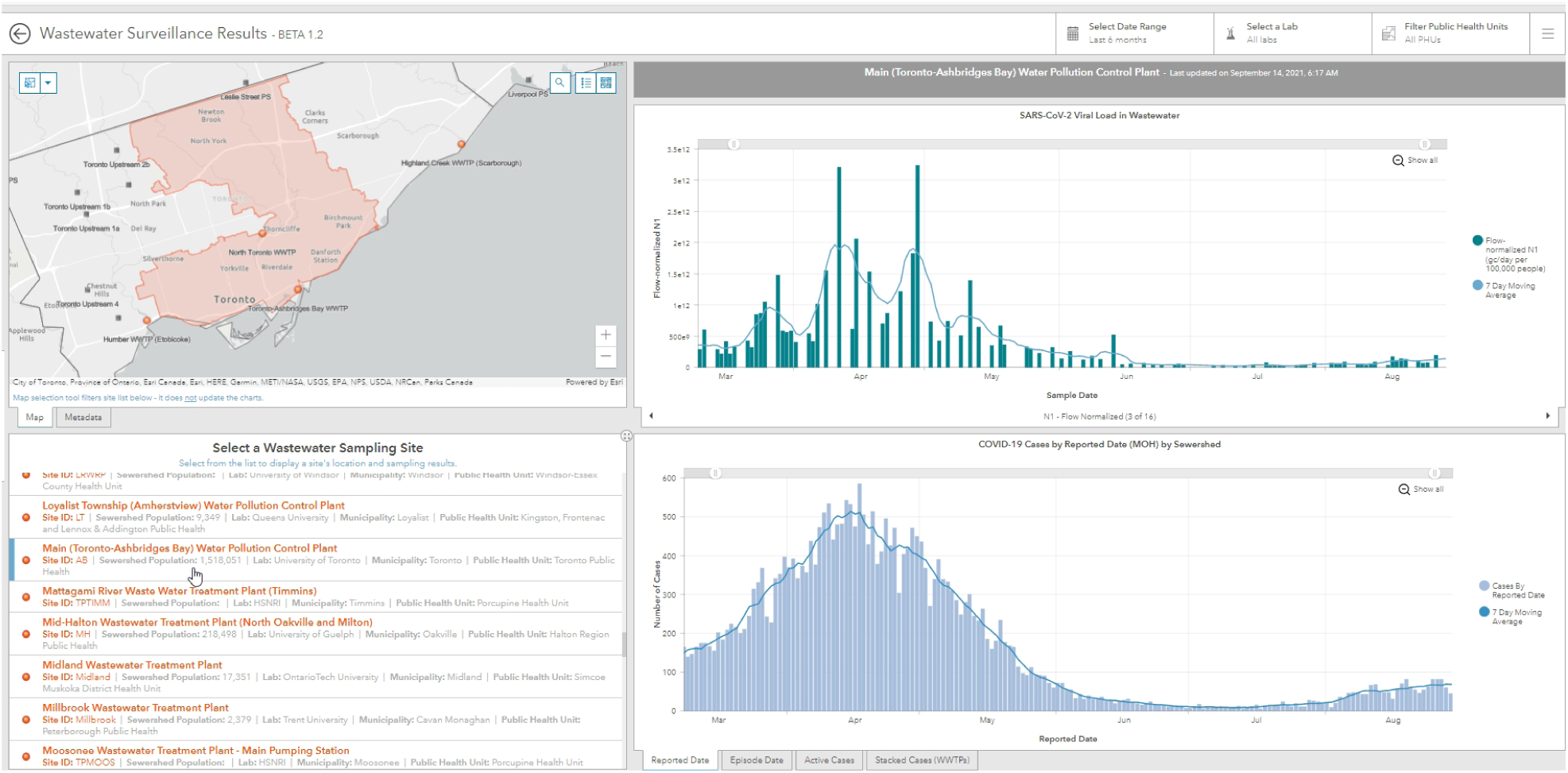
Toronto Ashbridges Bay sewershed with PMMoV-normalized N1 concentration (*C*_*vb*_) and clinical case data from the Ontario Dashboard

#### 3.1.1. PMMoV Normalization (C_vb_) and Cases by Reported Date

Figure 4 provides the longer-term QTA of clinical cases by reported date and the PMMoV-normalized wastewater viral concentration from March 21/21 to August 22/21. Table 4 summarizes the trend results with the 95% confidence interval (CI) and Table 5 provides the interpretations based on the PHAC interpretations given in Table 2. The standard errors (SE) associated with the estimated breakpoint and trend slopes for both the CCC and WVS, for each time interval, are provided in Tables 6 and 7, respectively. This is an example of the complete QTA 2-page model report for the WVS metric that may be used directly to inform public health decisions.

**Figure 4.**
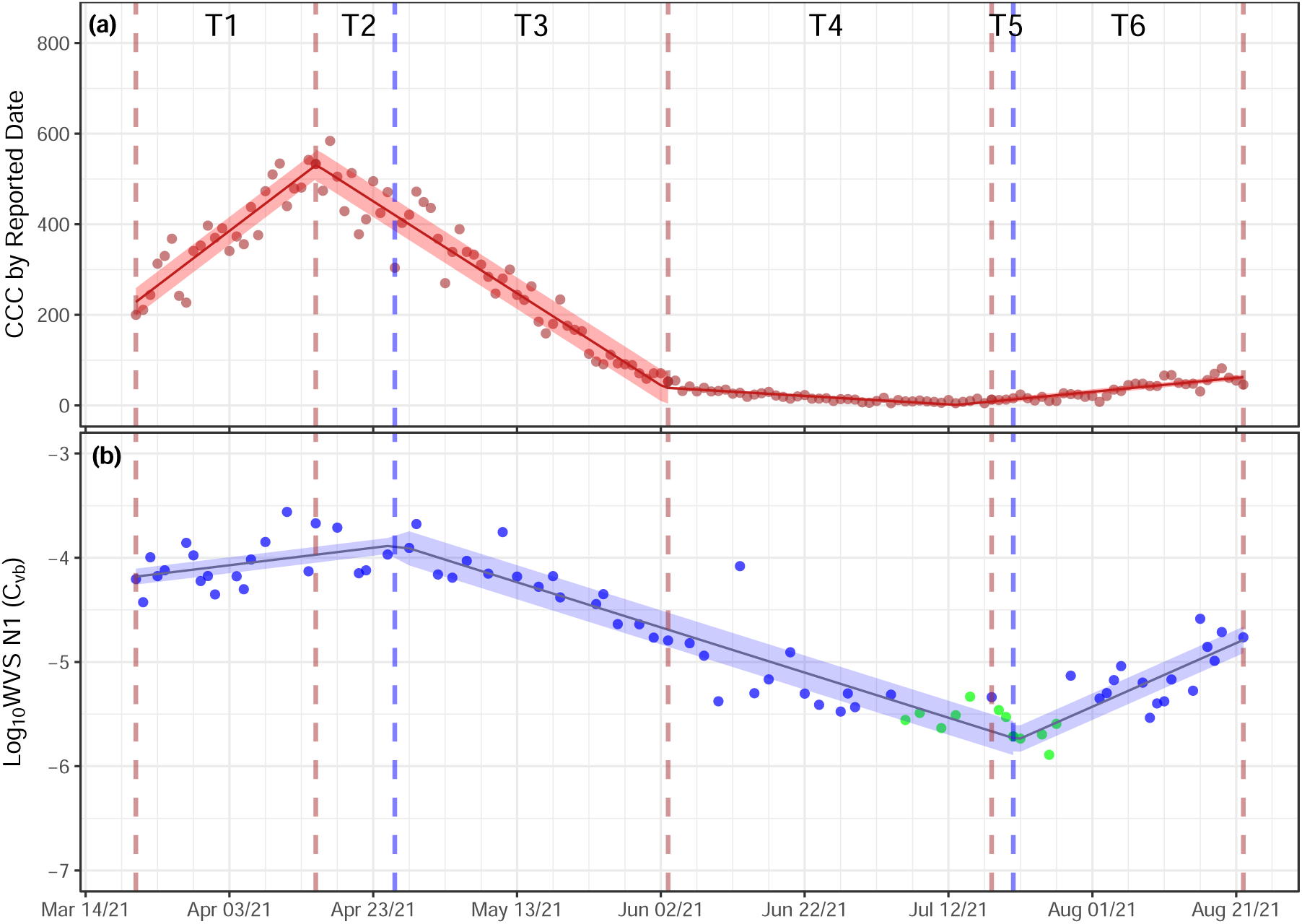
The TAB sewershed QTA of clinical cases by reported date (a) and log_10_ PMMoV-normalized N1 (*C*_*vb*_) viral concentration (b) (March 21/21 - August 22/21)

**Table 4.**
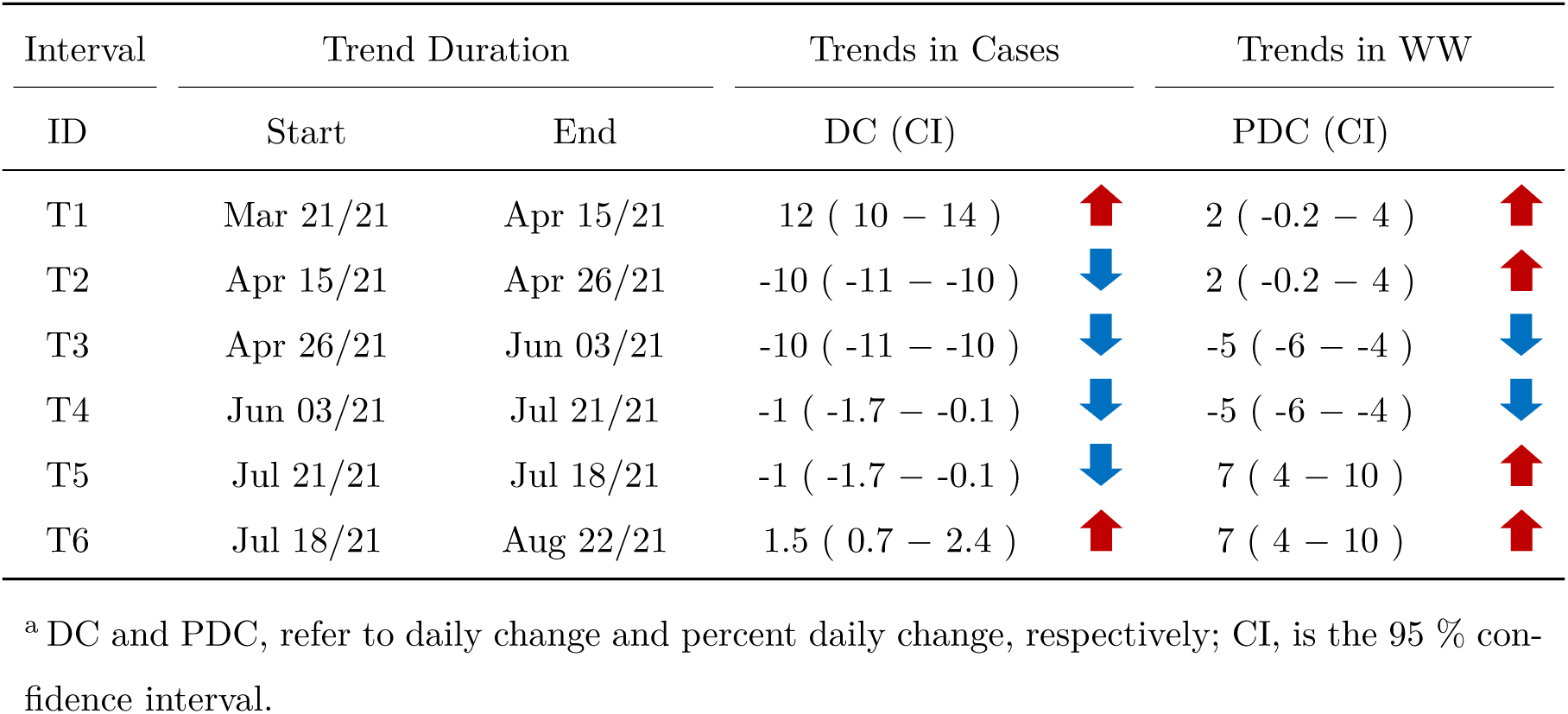
Summary aggregated trend results for the TAB WWTP sewershed^*a*^

| Interval | Trend Duration |  | Trends in Cases |  | Trends in WW |  |
| --- | --- | --- | --- | --- | --- | --- |
| ID | Start | End | DC (CI) |  | PDC (CI) |  |
| T1 | Mar 21/21 | Apr 15/21 | 12 ( 10 – 14 ) | ↑ | 2 ( -0.2 – 4 ) | ↑ |
| T2 | Apr 15/21 | Apr 26/21 | -10 ( -11 – -10 ) | ↓ | 2 ( -0.2 – 4 ) | ↑ |
| T3 | Apr 26/21 | Jun 03/21 | -10 ( -11 – -10 ) | ↓ | -5 ( -6 – -4 ) | ↓ |
| T4 | Jun 03/21 | Jul 21/21 | -1 ( -1.7 – -0.1 ) | ↓ | -5 ( -6 – -4 ) | ↓ |
| T5 | Jul 21/21 | Jul 18/21 | -1 ( -1.7 – -0.1 ) | ↓ | 7 ( 4 – 10 ) | ↑ |
| T6 | Jul 18/21 | Aug 22/21 | 1.5 ( 0.7 – 2.4 ) | ↑ | 7 ( 4 – 10 ) | ↑ |

<sup>a</sup> DC and PDC, refer to daily change and percent daily change, respectively; CI, is the 95 % confidence interval.

**Table 5.**
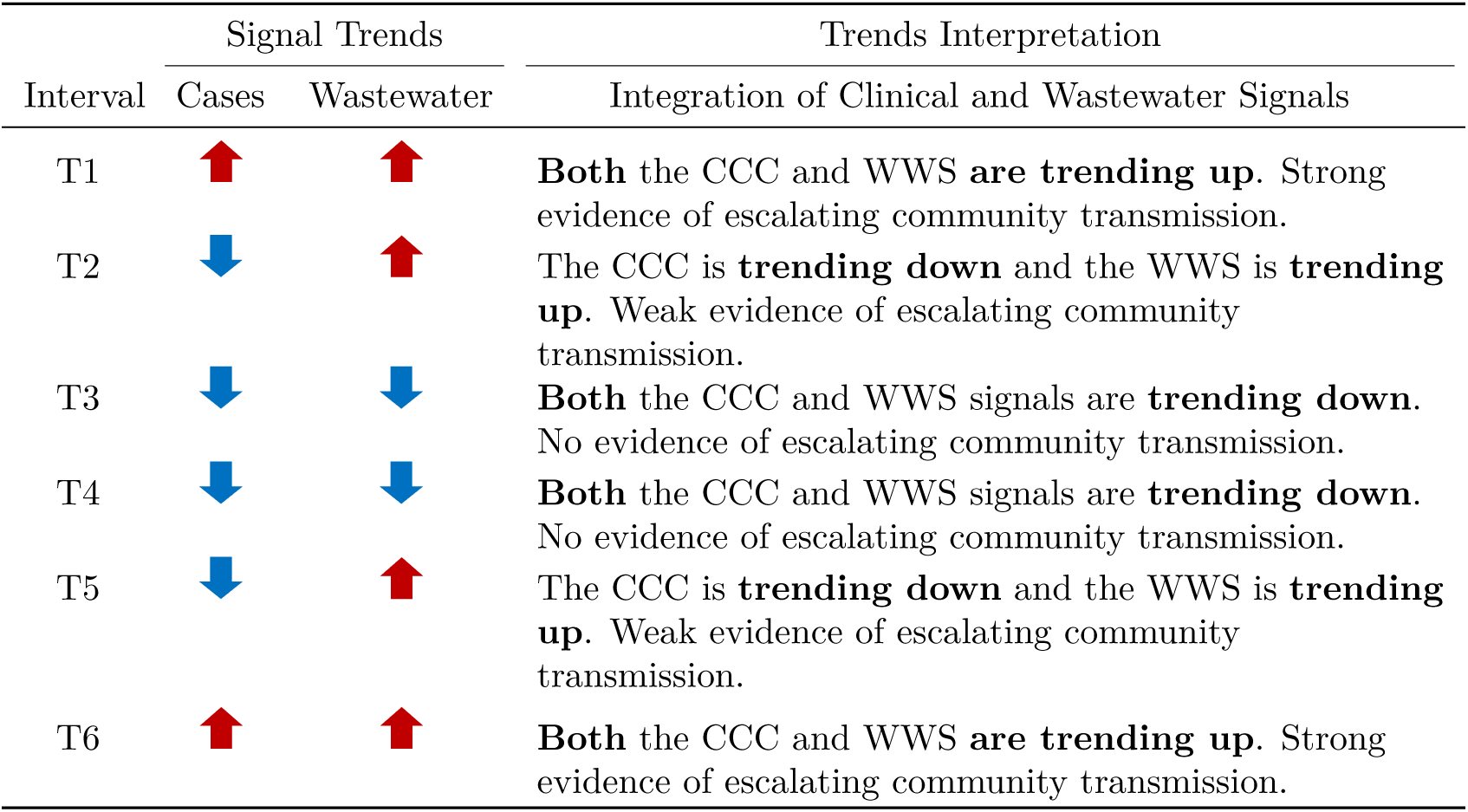
Summary interpretation of trend results for the TAB WWTP sewershed

**Table 6.**
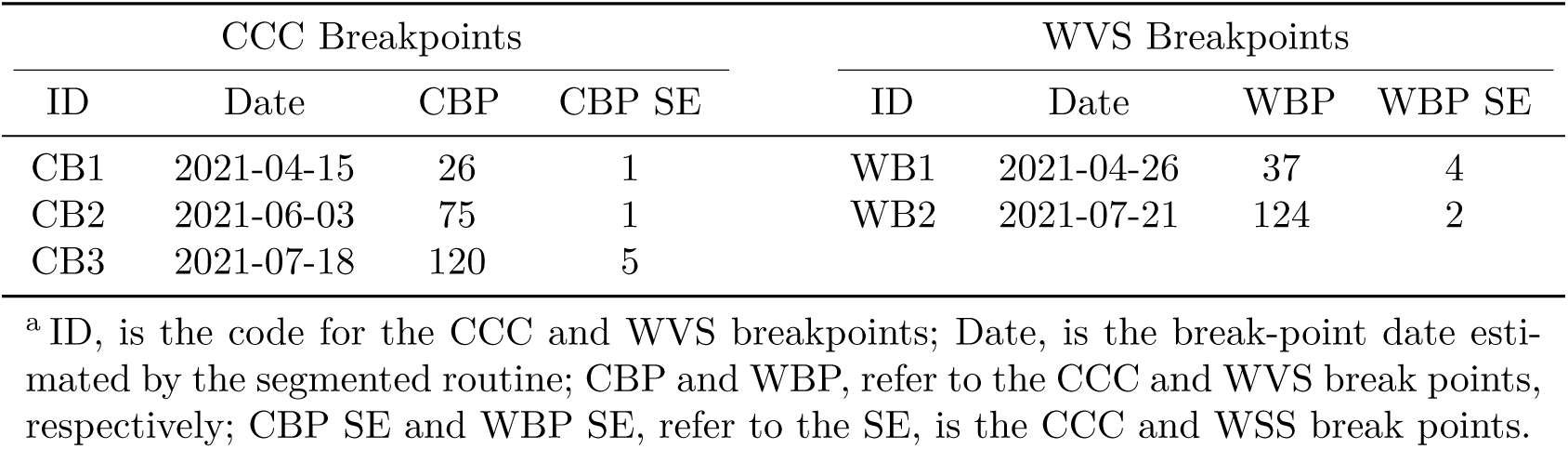
The CCC and WVS breakpoints (CBP and WBP) and associated SE^*a*^

| CCC Breakpoints |  |  |  | WVS Breakpoints |  |  |  |
| --- | --- | --- | --- | --- | --- | --- | --- |
| ID | Date | CBP | CBP SE | ID | Date | WBP | WBP SE |
| CB1 | 2021-04-15 | 26 | 1 | WB1 | 2021-04-26 | 37 | 4 |
| CB2 | 2021-06-03 | 75 | 1 | WB2 | 2021-07-21 | 124 | 2 |
| CB3 | 2021-07-18 | 120 | 5 |  |  |  |  |

<sup>a</sup> ID, is the code for the CCC and WVS breakpoints; Date, is the break-point date estimated by the segmented routine; CBP and WBP, refer to the CCC and WVS break points, respectively; CBP SE and WBP SE, refer to the SE, is the CCC and WSS break points.

**Table 7.** The CCC and WVS trends and associated standard errors (SE)^*a*^

| CCC Trend Lines |  |  |  | WVS Trend Lines |  |  |  |
| --- | --- | --- | --- | --- | --- | --- | --- |
| ID | n | DC | DC SE | ID | n | PDC | PDC SE |
| CS1 | 25 | 12.1 | 0.8 | WS1 | 21 | 2.0 | 1.1 |
| CS2 | 49 | -10.2 | 0.3 | WS2 | 36 | -4.9 | 0.3 |
| CS3 | 45 | -0.9 | 0.4 | WS3 | 24 | 7.3 | 1.3 |
| CS4 | 35 | 1.5 | 0.4 |  |  |  |  |

<sup>a</sup> ID, is the identificaiton code for the CCC and WVS trend lines; n, is the number of data points within each trend interval; DC, is the daily change or slope in cases per day for the CCC; PDC, is the percent daily change for the WVS or the slope of the trend line within each interval; DC SE and PCD SE, refer to the standard error of the DC and PDC, respectively and correlate to the error of the slope of the linear trend line within each interval.

#### 3.1.2. PMMoV Normalization (C_vb_) with WCR

This section introduces the WCR metric to demonstrate the versatility of the QTA approach and constitutes Part 2 of 2 of the model QTA report. Here we adopt the interpretations provided by Xiao et al, 2021 [8] within the QTA framework and this is provided in Figure 5 and Tables 8 − 11 for the TAB sewershed longer-term QTA.

**Figure 5.**
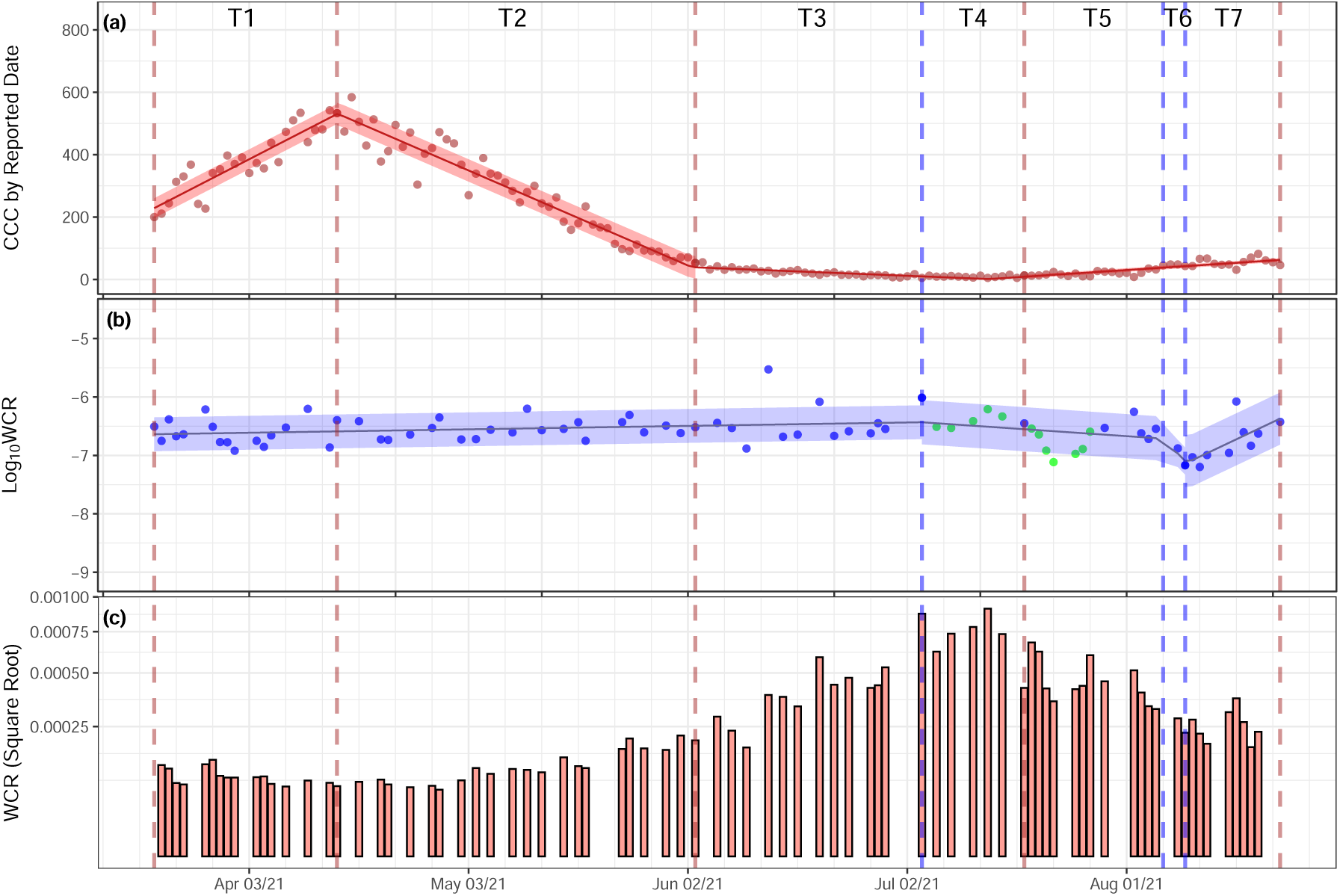
The TAB sewershed trend analysis of clinical cases by reported date (a) and log_10_ WC trend analysis (b) and WC values (c).

**Table 8.**
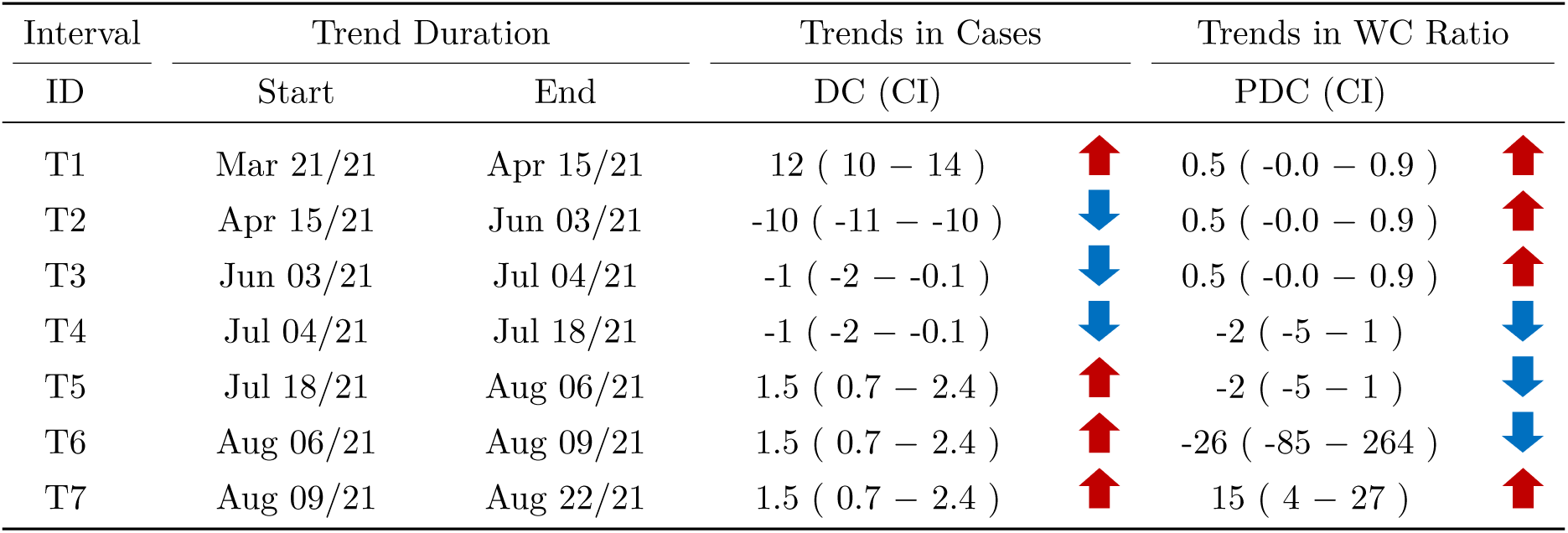
Summary aggregated trend results for the TAB WWTP sewershed

**Table 9.**
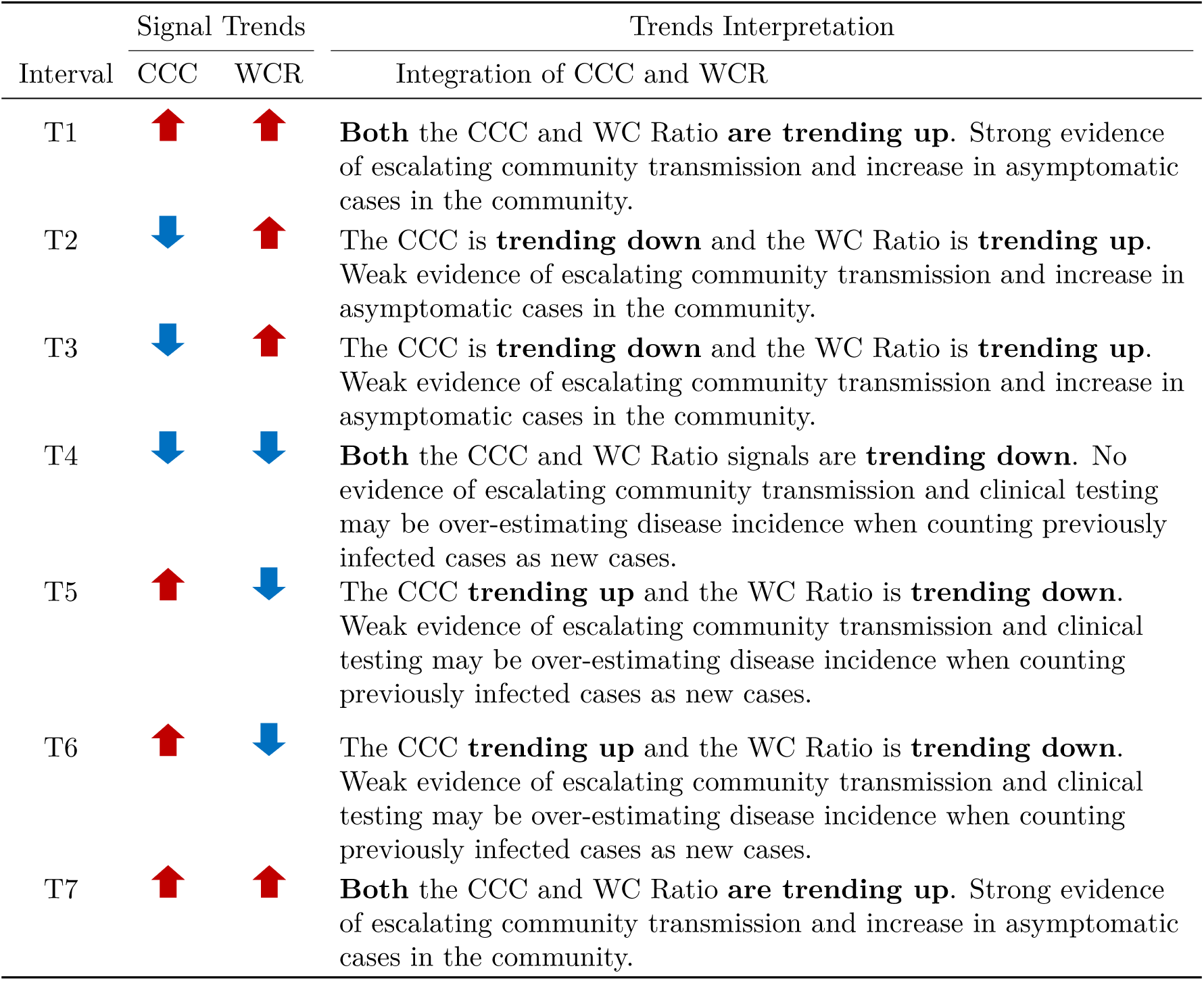
Summary interpretation of trend results for the TAB WWTP sewershed CCC and WCR

**Table 10.** The CCC and WVS estimated breakpoints and associated standard errors^*a*^

| CCC Breakpoints |  |  |  | WCR Breakpoints |  |  |  |
| --- | --- | --- | --- | --- | --- | --- | --- |
| ID | Date | CBP | CBP SE | ID | Date | WCBP | WCBP SE |
| CB1 | 2021-04-15 | 26 | 1 | WB1 | 2021-07-04 | 106 | 14 |
| CB2 | 2021-06-03 | 75 | 1 | WB2 | 2021-08-06 | 139 | 7 |
| CB3 | 2021-07-18 | 120 | 5 | WB3 | 2021-08-09 | 142 | 2 |

<sup>a</sup> ID, is the code for the CCC and WC breakpoints; Date, is the breakpoint date estimated by the segmented routine; CBP and WCBP, refer to the CSS and WC breakpoints; CBP SE and WCBP SE, refer to the SE of the CSB and WCBP, within each interval.

**Table 11.** The CCC and WC estimated trend lines slopes and associated standard errors^*a*^

| CCC Trend Lines |  |  |  | WCR Trend Lines |  |  |  |
| --- | --- | --- | --- | --- | --- | --- | --- |
| ID | n | DC | DC SE | ID | n | PDWC | PDWC SE |
| CS1 | 26 | 12.1 | 0.8 | WS1 | 53 | 0.5 | 0.2 |
| CS2 | 49 | -10.2 | 0.3 | WS2 | 17 | -1.9 | 1.5 |
| CS3 | 45 | -0.9 | 0.4 | WS3 | 0 | -25.8 | 121.9 |
| CS4 | 35 | 1.5 | 0.4 | WS4 | 11 | 14.6 | 5.1 |

<sup>a</sup> ID, is the code for the CCC and WC trend lines; n, is the number of data points within each interval; DC and DW, are the daily change in the CSS and percent daily change (PDC) of the WC of the trend line (or slope) within each interval; DC SE, and PDWC SE, refer to the standard error in the corresponding units of the DC and PDWC.

#### 3.1.3. TAB Correlation of Wastewater to Clinical Case Counts

An assessment of the linear and non-linear correlation between *C*_*vb*_, *L*_*v*_, *L*_*vb*_ and *mN* 1 (un-normalized N1) and clinical case counts by reported data were also considered (Figure 6, Table 12). In terms of the Linear correlation results and the *R*^2^)values, the PMMoV normalization (*C*_*vb*_) was slightly better than the un-normalized (*mN* 1) which was better than the flow normalization (*L*_*v*_) and the mean PMMoV-flow normalization (*L*_*vb*_) for the TAB WWTP, sequentially. However, when considering the non-linear correlation results, the AICcWt factor which refers to the proportion of the total predictive power that can attributed to the model, it was highest (0.63) for the the mean PMMoV-flow normalization (*L*_*vb*_). The adjusted coefficient of determination ranged from 0.79 – 0.85 and polynomial of order 6 – 8 provided the best model fit (based on the minimum AICc up to a polynomial of order 9). Based on the linear analysis the *C*_*vb*_ is the preferred normalization mode when considering the gene target N1 and based on the non-linear correlation the *L*_*vb*_ is preferred. Also considering that the adjusted *R*^2^ (*aR*^2^) is higher than the *R*^2^ for both *C*_*vb*_ and *L*_*vb*_ using the PMMoV-flow normalization is preferred over the PMMoV normalization. Similar screening level analyses can be conducted for N1N2 or N2 prior to settling on a specific normalization mode that can then be used consistently for a given sewershed.

**Figure 6.**
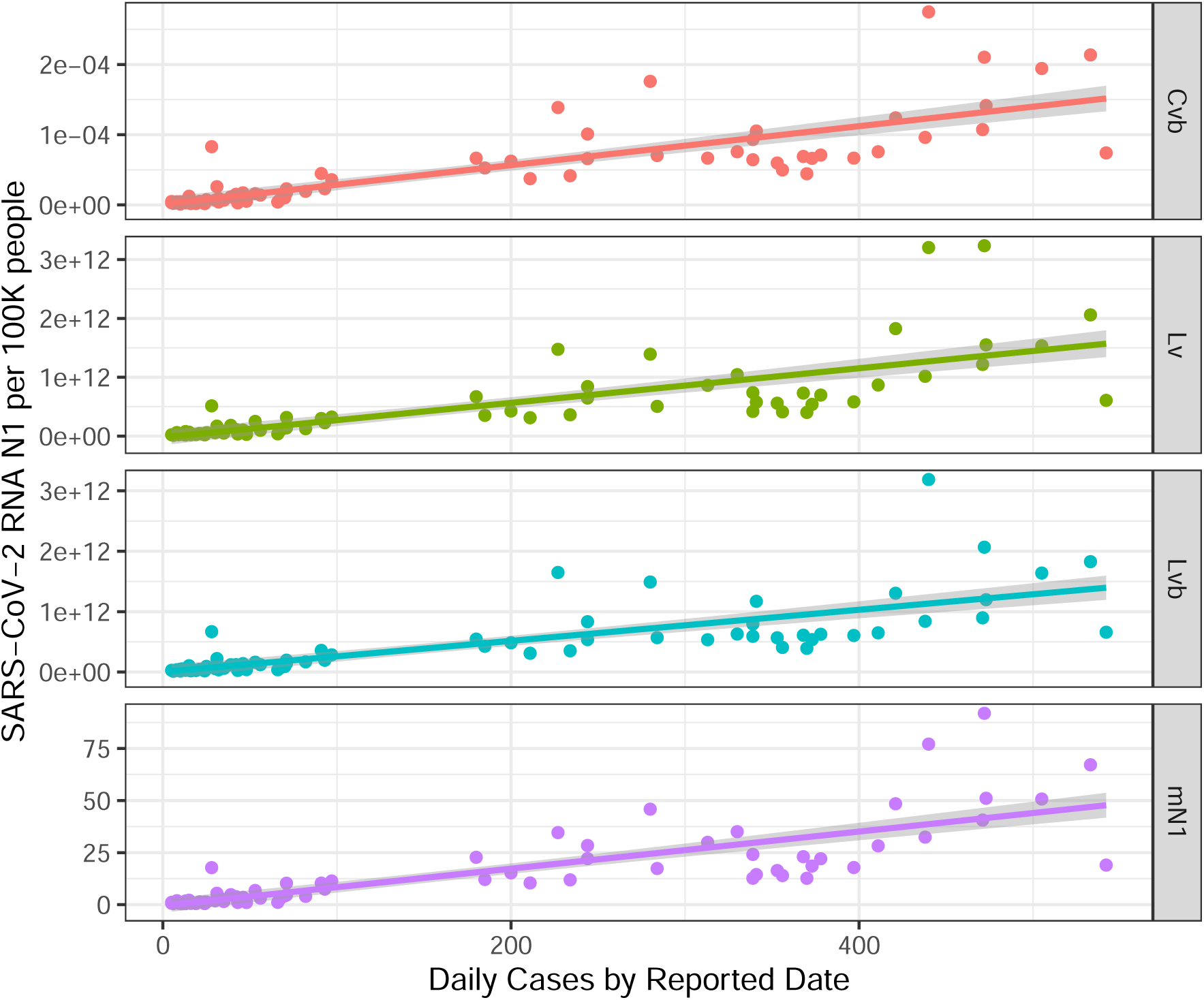
The TAB sewershed linear correlation normalizations for N1 (biomarker normalized (*C*_*vb*_), biomarker-flow noarmalized (*L*_*vb*_), flow-normalized (*L*_*v*_) and un-normalized (*mN* 1) versus clinical cases by reported date (Feb 14/21 - July 13/21) with 95% confidence band

**Table 12.** TAB N1 WVS (*C*_*vb*_, *L*_*v*_, *L*_*vb*_, mN1) versus the daily CCC per 100,000 people correlation results^*a*^

| Normalization | Linear Correlation Results |  |  | Nonlinear Correlation Results |  |  |  |
| --- | --- | --- | --- | --- | --- | --- | --- |
| | Slope ± SE | $R^2$ | P-Value | AICc | AICcWt | $aR^2$ | PO |
| $C_{vb}$ | $3e-07 \pm 2e-08$ | 0.67 | $< 0.01$ | -1489 | 0.52 | 0.84 | 6 |
| $L_{vb}$ | $3e+09 \pm 2e+08$ | 0.59 | $< 0.01$ | 4395 | 0.63 | 0.79 | 6 |
| $L_v$ | $3e+09 \pm 3e+08$ | 0.59 | $< 0.01$ | 4384 | 0.45 | 0.79 | 6 |
| $mN1$ | $9e-02 \pm 7e-03$ | 0.66 | $< 0.01$ | 538 | 0.36 | 0.85 | 8 |

<sup>a</sup> Slope ± SE, is the slope and standard error associated with the linear correlation regression; $R^2$ , is the coefficient of determination; p-value, is the probability that the predictor (WVS) is related to the response (CCC) compared to the null hypothesis of being unrelated; AICc, is the Akaike information criterion; AICcWt, is the AICc weight; $aR^2$ , is the adjusted $R^2$ and PO, is the polynomial order of the best fit curve up to order 9.

## 4. Summary Findings

The methodology of the QTA (Figure 1) provided a means to integrate the three key steps of data normalization, trend analysis and interpretation in a semi-automated approach to reduce personal bias.

The normalization methods of the WVS considered included PMMoV as a biomarker (*C*_*vb*_), the mean 24-hour daily flow rate (*L*_*v*_) and combined median PMMoV-flow (*L*_*vb*_) (Eq. 1 – 3) all generated based on the reported N1 primer results as gene counts per milliliter (gc/mL). In all cases the wastewater samples collected represented 24-hour composite samples from the influent of the corresponding wastewater treatment plants. Un-normalized mean N1 (*mN* 1) was only considered in the linear trend analysis. We recommend the decision of the SARS-CoV-2 primer to select for the QTA (N1, N2, N1N2, E, others) should be done in consultation with the laboratories conducting the RT-qPCR analysis of the wastewater samples. Generally, the type of normalization most appropriate was found to be sewershed specific and dependent on the time scale considered, but in some cases significant similarities were observed between the different normalization methods. Additional effort is required to collect and analyze the necessary metadata (flow) so this should be considered at the planning stages.

To automate the trend analysis, the minimum AIC was introduced as a means to select the best polynomial-fit and to determine the seed inflection points for the segmented break-point linear regression model. Despite the noisy data sets, this method was found effective using a 2 – 9 order polynomial (*m* = 2 − 9, step 2 (iv) Figure 1). However, depending on the resolution required and data sets considered, the upper value of *m* may be readily increased to serve the purpose and computed seed inflection points may be manually selected for the intended purpose. An additional consideration of the method included error propagation with the standard error (SE) and 95 % confidence interval (CI), associated with the breakpoints and trends. As the data set sample size (*n*) is reduced the corresponding SE and CI increase and reducing the confidence level in the final results and potentially compromising the interpretations. Sample size of *n <* 3 is not recommended to determine trends. As an example, in our QTA analysis for the current data sets we observed a trend based on *n* = 3 (Figure S5, THR shorter term *C*_*vb*_ CCC by reported date) which also included the highest SE and CI that crossed the zero value and did not prove to be significant at the 95 % confidence level.

The interpretation relied on pre-determined interpretations considering the integration of the CCC and WVS or WCR trend results provided in Table 2 and 3. These interpretation need to be considered within the local context to best inform public health decisions.

Specific summary findings for each sewershed (TAB, THR, THC and TNT) are provided below with reference to the the Supplement where additional results for each sewershed were provided.

### 4.1. TAB Sewershed

The TAB sewershed is a combined system and was the largest in land area (25,000 ha) and most populous (1.6 M) (Table 1) of the four sewersheds within the TPH jurisdiction. The TAB sewershed data set was used to assess a longer-term QTA analysis (March 14/21 – August 22/21) with different normalizations (*C*_*v*_, *L*_*v*_, *L*_*vb*_), an assessment of CCC by reported data versus episode date along with the use of the WCR compared to the more direct WVS. The Supplement includes additional QTA results using *L*_*v*_ and *L*_*vb*_ normalizations shown in Figures S1 – S3 and Tables S1 – S12.

From Figure 4 *C*_*vb*_, we can see that the WVS trends up past the peak of the CCC (end of interval T1) and continues to trend up for 14-days (end of interval T2) while cases are trending down during interval T2. This may be indicative that virus shedding continues to increase (additive loadings by active cases) within the community up to the beginning of interval T2. This was consistently observed in the *L*_*v*_, *L*_*vb*_ normalizations for CCC by reported date and also for *L*_*vb*_ with CCC by episode date (Figures S1 – S3).

A comparison between *L*_*vb*_) using CCC by episode date versus reported date (Figures S2 and S3) shows differences in the number of intervals and timing of the main transition points. However, the trend lines show a consistent direction and slope regardless of how the CCC were reported.

During interval T3 – T6 or T7 of Figure 4 and Figures S1 – S3, the trends are in concordance with some lag observed juxtaposing between the CCC and WVS. No consistent lead or lag was observed for the TAB sewershed data set.

A baseline in cases was observed during June 3/21 – July 18/21 (interval T4 and T5) with a corresponding baseline in the WVS during T5 and T6. During this period also a significant percentage of the WVS was below the laboratory reported LOD of about 1 gc/mL. The CCC during this period were between 10 – 50 within the service population of 1.6 million with rate of change from 0 – 2 cases per day. This low rate in CCC shows a baseline response in the WVS up until when the CCC counts exceeds about 40 CCC or about 1 in 40,000 reported cases. But it is roughly estimated that the reported cases are an underestimate of the shedding population by a factor in the range of 5 to 10, when considering asymptomatic cases. Taking the lower range of this factor provides a rough estimate of the resolution of the wastewater signal to be in the order of about 1 in 10,000 cases. Further work in enumerating and estimating fecal shedding rates, within well defined groups, such as university residences, has been conducted and it is expected that these results would assist in determining better estimates of community prevalence though the use of models [35].

### 4.2. THR Sewershed

The THR sewershed serves a population of 685,000 with an area of 18,000 ha having a partially combined sewer system with a calculated median day sewage flow rate of 234 ML/d, during the sampling campaign. The Supplement includes the QTA results regarding the short-term trend analyses for the THR sewershed for *C*_*vb*_, *L*_*v*_ and *L*_*vb*_ normalization modes based on CCC by reported date with linear correlation analysis (Figures S4 – S8 and Tables S13 – S25).

From Figure S5 *C*_*vb*_ and Table S13 we can see that the WVS trends up marginally but close to a baseline and generally not responsive to the CCC increases (from 5 to 10 and from 10 to 35 cases) during intervals (T1 T3) from July 6/21 to August 23/21. Only during interval T4 (August 23 – 25/21) when CCC increased above 40 do we see a response and increase in the WVS trend. However, throughout this time period, the WVS trends are not statistically significant (CI range crosses zero) and this may be related to being at the apparent limit of resolution. A closer look shows that the CCC are below 10 during T1, less than 40 during interval T2 and less than 50 during T3 – T4 and assuming a factor of 10 in community cases compared to the CCC, we have a ratio in the order of less than 1/10,000 based on a population served of 685,000. From Figure S5 its also observed that 16/18 (89 %) WVS data points are below the LOD (color coded green) of 2.5 gc/mL during T1 and suggests that we are at the limit of resolving the change in cases during these event up to when the CCC increases to above the order of 1/10,000.

Comparing the WVS trends using flow normalization (*L*_*v*_) compared to the *L*_*vb*_ and *C*_*vb*_ normalizations, shows an increase in the number of intervals or break-points (6 versus 4) suggesting the flow normalization, at low CCC, shows higher variability. This may be attributed to the fact that the THR sewershed is a partially combined system.

A review of the correlation of the CCC by reported date to the WVS by 100K population (Figure S8 and Table S25) shows a poor linear data correlation (*R*^2^ *<* 0.1 and p-value *>* 0.03, generally). The non-linear correlation analysis identifies the *L*_*v*_ and *mN* 1 linear models to provide and AICcWt of 0.63 to 0.68 but with a *aR*^2^ of 0.11 an d0.04. It appears that when a significant part of the WVS data set is below the LOD (19/36 or 53% of the data points) or close to the LOD, reflecting low changes in CCC, the changes in WVS trends may not be significantly reflective of trends low CCC changes.

### 4.3. THC Sewershed

The THC sewershed serves a population of 533,000 with an area of 10,600 ha having a separated sewer system with a calculated median day sewage flow rate of 164 ML/d, during the sampling campaign. The Supplement includes the QTA results regarding the shorter-term trend analyses for the THC sewershed for *L*_*vb*_ normalization modes based on CCC by reported date and WC ratio with linear correlation analysis (Figures S9 – S12 and Tables S26 – S34).

From Figure S10 (*L*_*vb*_) we see that the WVS and CCC are in concordance during interval T1 and T3 but discordant during T2 and in all cases the WVS trends are statistically significant with all data above the reported LOD. During this period (March 15/21 – May 15/21) the CCC by reported date range from 50 to 300 with a daily increase of about 7 cases per day and a decrease during interval T3 of about 6 cases per day. The results of this QTA suggests how the change and transition from strong evidence fo escalating community transmission to weak evidence and no evidence of escalating community transmission may be informed by the WVS trends, statistics and interpretations (Tables S26 – S29).

Figure S11 (WC ratio of $L_{vb}) shows the WC ratio and CCC within four intervals (T1 – T4). During interval T1 and T3 the WC ratio supports the up-trend of CCC suggesting that there is an increase in asymptomatic cases in the community. During interval T2 the WC ratio is trending down while the CCC is trending up and the WC ratio trend results is suggesting that clinical testing may be over-estimating when counting previously infected cases as new cases. During interval T4 the CCC cases are trending down while the WC ratio is trending up which is suggesting that asymptomatic cases in the community are increasing despite the decreasing CCC trend indicating weak evidence of escalating community transmission.

Figure S12 provides linear correlation of the CCC by reported date to the WVS by 100K population. Table S34 shows a weak but significant linear data correlation (*R*^2^ *<* 0.24 and p-value *<* 0.02, generally). The non-linear AICwt is between 0.54 – 0.63 with *C*_*vb*_ and *L*_*v*_ normalization providing the best model correlation. This data set differs from the THR data set by not having any data below the LOD. However the the weak linear correlation appears to have contributed to a statistically insignificant trends in 3 of the 4 time intervals at the 95% confidence level and this is attributed to smaller sample size (*n*) when compared to the daily CCC.

### 4.4. TNT Sewershed

The TNT sewershed serves an equivalent population of about 55,000 with an area of 5,000 ha having a combined sewer system with a calculated median day sewage flow rate of 18 ML/d, during the sampling campaign. The Supplement includes the QTA results regarding the longer-term trend analyses (March 14/21− August 29/21) for the TNT sewershed for *C*_*vb*_, *L*_*vb*_ and *L*_*v*_ normalizations compared to CCC by reported date and WC ratio with linear correlation analysis (Figures 14 – S17 and Tables S35 – S46).

Figure S14 using *C*_*vb*_ normalization, shows the CCC and WVS trends over seven time intervals (T1 – T7) and similarly to the TAB longer-term QTA results the WVS, WVS trends up past the peak of the CCC (end of interval T1) and continues to trend up for 13-days (end of interval T2) while cases are trending down during interval T2. This appears to be a common phenomena most evident in the longer-term QTA results and attributed to continued virus shedding in the community beyond the peak observed in CCC.

Further from Figure S14, during interval T3 and T4, both the WVS and CCC are trending down in concordance. During interval T5 the CCC trend is at baseline and the WVS is trending down but at a low level with much of the data below the LOD and this continues during interval T6 and T7. During T6 and T7 the WVS does not change and does not respond to the increase and decrease in CCC while the trends in cases are less than 1 case/day. For the TNT sewershed, the QTA results are similar for both the *C*_*vb*_ and *L*_*v*_ normalizations (Figure S15 and Tables 39 – S42) using the WVS metric directly.

The trends (Figure S16 and Table S43 – S45) are increasing during intervals T1 – T3 and T7 and this suggests that clinical testing may be over-estimating disease incidence when counting previously infected cases as new cases. During intervals T4 – T6 the trends are decreasing which suggests that clinical testing may be over-estimating disease incidence when counting previously infected cases as new cases.

Figure S17 provides linear correlation of the CCC by reported date to the WVS by 100K population. Table S47 shows a good and significant linear data correlation (0.63 *≤ R*^2^ *≤* 0.73 and p-value *<* 0.01) as well as non-linear correlation. Based on the AICcWt value of 1.0 and 0.98 the *C*_*vb*_ and *L*_*vb*_ normalization modes are preferred. This data set is similar to the TAB data set in correlation values (Table 12 and showing similar consistent properties in terms of significant trends.

## 5. Conclusions and Recommendations

The primary purpose of this work was to demonstrate the utility of the QTA approach to help inform public health decisions. Data sets from four urban sewersheds within the Toronto PHU were analyzed using QTA both over a longer-term (5-months) and shorter-term (4 – 8-weeks) and under different normalizations (*C*_*vb*_, *L*_*v*_, *L*_*vb*_) and with different metrics (WCR and WVS). Our QTA demonstrated clearly quantifiable trends from real data for different sewersheds with data from two different laboratories under different time periods. Further evaluations for smaller municipal sewershed has been conducted (not reported here) and has shown the robustness of the QTA process and this is currently being documented.

The longer-term (5-months) QTA for TAB and TNT sewersheds demonstrated that PMMoV-normalized (*C*_*vb*_) and median flow-PMMoV normalization (*L*_*vb*_) generally provide more conservative results, in terms of increased WVS prevalence, over flow-normalized (*L*_*v*_) SARS-CoV-2 RNA data. The shorter-term (5 – 8 weeks) QTA on THC and TNT sewersheds provided similar findings to the longer-term analysis and provided some additional insights into the potential impact of the data at or below the method LOD.

Overall the major WVS and CCC trends were captured by the three normalizations (*C*_*vb*_, *L*_*v*_ or *L*_*vb*_) however, significant differences in trends were observed particularly when the data contained a large percentage (over 50%) of data below the reported LOD. To assess the more conservative and recommended normalization approach, for any given sewershed, it is suggested that an initial comparative assessment of the QTA process using different normalizations be conducted and assessed based on a linear and non-linear assessment based on the AICcWt criteria to determine the appropriate normalization mode. Further, the extent of clinical testing and the underlying assumption that it reflects the general population cases, should be considered carefully as part of the integration of the WVS and CCC for interpretation. It is expected, that situations will arise (e.g., due to reduced severity of symptoms clinical testing becomes less prevalent) that may make this integration less relevant. In such situations, the WVS may become a determining stand alone representative metric.

Since the ratio *L*_*vb*_*/C*_*vb*_ is the median PMMoV divided by the sample PMMoV 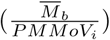 these normalization modes may yield similar QTA results particularly when this ratio is relatively constant during the sampling campaign within a given sewershed.

Further testing on the QTA method is ongoing to further automate the process to generate complete analysis and interpretations for different sewershed types and normalizations.

The work presented in this manuscript also served as a case study and pilot to assess the benefits of the QTA process for production level reporting. The analysis, based on PMMoV-normalized wastewater signals, was well received with a positive feedback from the TPH and PHAC. It is noteworthy, that in the case of the sewersheds within the TPH jurisdiction, the QTA analysis showed that the WVS was generally not predictive or did not lead the CCC. Despite this, the QTA served as a complimentary and confirmatory metric. To this end it is recommended that QTA continue to be evaluated and used to generate timely interpretative quantitative reports to support public health decisions as a complementary tool to clinical data information.

## Data Availability

All data produced in the present study are available upon reasonable request to the authors

## 8. Acknowledgments

The authors extend their appreciation to: The *City of Toronto and Toronto Water Team* who supported and contributed to the wastewater field sampling and surveillance initiative including, Eliav Eini and Eris Prod for providing the essential flow data for each WWTP site; the *Ryerson University Team* including: Hussain Aqeel, Peter Vi, Hooman Sarvi, Farnaz Farahbakhsh, Devon Hock, Neil Kennedy, Alexandra Johnston, Eden Hataley, Wynona Klemt; the *MECP Geomatic and Dashboard Data Team* including Zita Lo, Ela Lichtblau, Ante Girzelj, Menglu Wang, Ian Bowles and Ric Mucciacciaro; the *R-coding team* including Vince Pileggi for developing the concept and writing the R-code for the complete QTA analysis and reporting; Melanie Raby for updating, maintaining and extending the original R-code by Vince Pileggi for the *Hub* and for Zeinab Khansari for automating and validation of the R-code source for the *Hub*. The R-code (for data transcription not the focus of this publication) served the primary function of transcribing the data received from all the university labs onto the data *Hub* for visualization and data downloads; Dawn Benson, MECP legal counsel, for drafting the agreements between MECP and the Ontario universities; the MECP Wastewater Surveillance *Core Team* for leading the initiative including Steven Carrasco, Bahar Aminvaziri, Erin Harrigan Podgaiz, Maggie Tompkins, Carmen Ghetea and David Pipher; the *Technical Team* for providing valuable comments to the draft manuscript including Susan Weir, Cecily Anne Flemming, Ann-Marie Abbey, Albert Simhon, Vince Pileggi, Alex Ho Shing Chik and Tim Fletcher. John Minnery of Public Health Ontario for critical review and comments of the draft manuscript. Without the hard work and dedication of this multidisciplinary group of people this publication would not have been possible

