## Supplement for "Quantitative Trend Analysis of SARS-CoV-2 RNA in Municipal Wastewater Exemplified with Sewershed-Specific COVID-19 Clinical Case Counts"

The following additional QTA results (all based on N1) are included in this Supplement: Toronto Ashbridges Bay (TAB) sewershed longer term QTA: (1) WWS $L_v$ and CCC by reported date (Figure S1 and Tables S1 – S4); (2) WWS $L_{vb}$ and CCC by reported date (Figure S2 and Tables S5 – S8); (3) WWS $L_{vb}$ and CCC by episode date (Figure S3 and Tables S9 – S12); Toronto Humber River (THR) sewershed shorter term QTA: (4) WWS $C_{vb}$ and CCC by reported date (Figure S5 and Tables S13 – S16); (5) WWS $L_v$ and CCC by reported date (Figure S6 and Tables S17 – S20); (6) WWS $L_{vb}$ and CCC by reported date (Figure S7 and Tables S21 – S24); (7) Linear correlation of WWS to CCC by reported date (Figure S8 and Table S25); Toronto Highland Creek (THC) sewershed shorter term QTA: (8) WWS $L_{vb}$ and CCC by reported date (Figure S9 and S10 and Tables S26 – S29); (9) WCR $L_{vb}$ and CCC by reported date (Figure S11 and Tables S30 – S33); (10) Linear correlation of WWS to CCC by reported date (Figure S12 and Table S34); North Toronto (TNT) sewershed longer term QTA: (11) WWS $C_{vb}$ and CCC by reported date (Figure S13 and S14 and Tables S35 – S38); (12) WWS $L_v$ and CCC by reported date (Figure S15 and Tables S39 – S42); (13) WCR $L_{vb}$ and CCC by reported date (Figure S16 and Tables S43 – S46) and (14) Linear correlation of WWS to CCC by reported date (Figure S17 and Table S47).

This Supplement also includes a description of the Ontario Data Template (S15), the extended aggregated data set (EAD) (S16) and sample R-Code of key routines written to generate the QTA reports (S17).

*Corresponding Author

*Email addresses:* (Vince Pileggi), (Jayson Shurgold), (Jianxian Sun), (Minqing Ivy Yang), (Elizabeth Edwards), (Hui Peng), (Amir Tehrani), (Kimberley Gilbride), (Claire Oswald), (Shinthuja Wijayasri), (Dana Al-Bargash), (Rebecca Stuart), (Zeinab Khansari), (Melanie Raby), (Janis Thomas), (Tim Fletcher), (Albert Simhon)

### 1. Toronto Ashbridges Bay (TAB) Longer Term QTA Based on Flow-normalized N1 Viral Load ($L_v$) and Cases by Reported Date

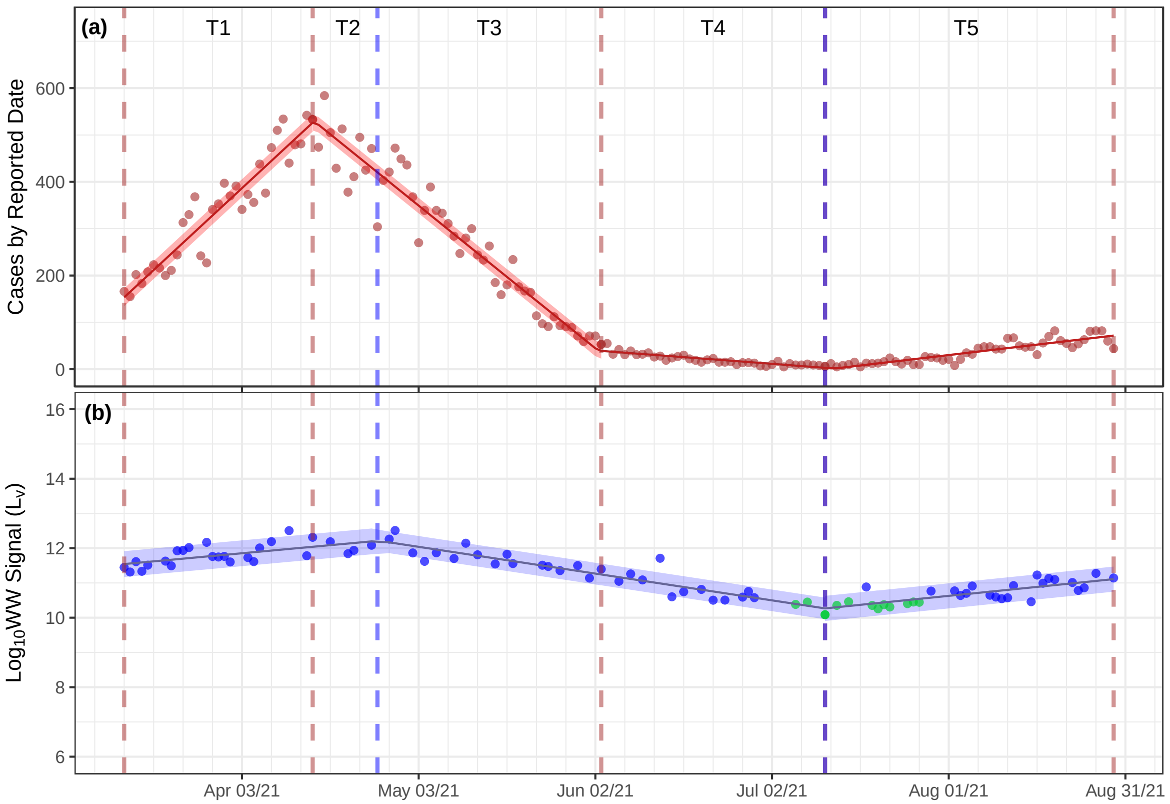


**Figure S1.** The TAB sewershed QTA of CCC by reported date (a) and $\log_{10}$ flow-normalized N1 ($L_v$) viral load.

**Table S1.** Summary aggregated trend results for the TAB WWTP sewershed[a]

| Interval | Trend Duration | | Trends in CCC | Trends in WW |
|---|---|---|---|---|
| ID | Start | End | DC (CI) | PDC (CI) |
| T1 | Mar 14/21 | Apr 15/21 | 12 ( 11 – 13 ) | 4 ( 2 – 5 ) |
| T2 | Apr 15/21 | Apr 26/21 | -10 ( -11 – -10 ) | 4 ( 2 – 5 ) |
| T3 | Apr 26/21 | Jun 03/21 | -10 ( -11 – -10 ) | -6 ( -6 – -5 ) |
| T4 | Jun 03/21 | Jul 11/21 | -1 ( -1.7 – -0.2 ) | -6 ( -6 – -5 ) |
| T5 | Jul 11/21 | Aug 29/21 | 1.5 ( 0.9 – 2.1 ) | 4 ( 3 – 5 ) |

[a] DC and PDC, refer to daily change and percent daily change, respectively; CI, is the 95 % confidence interval.

**Table S2.** Summary interpretation of trend results for the TAB WWTP sewershed

| | Signal Trends | | Trends Interpretation |
|---|---|---|---|
| Interval | CCC | WWS | Integration of CCC and WWS Signals |
| T1 | ↑ | ↑ | **Both** the CCC and WWS **are trending up**. Strong evidence of escalating community transmission. |
| T2 | ↓ | ↑ | The CCC is **trending down** and the WWS is **trending up**. Weak evidence of escalating community transmission. |
| T3 | ↓ | ↓ | **Both** the CCC and WWS signals are **trending down**. No evidence of escalating community transmission. |
| T4 | ↓ | ↓ | **Both** the CCC and WWS signals are **trending down**. No evidence of escalating community transmission. |
| T5 | ↑ | ↑ | **Both** the CCC and WWS **are trending up**. Strong evidence of escalating community transmission. |

**Table S3.** The CCC and WWS breakpoints (CBP and WBP) and associated SE[a]

| CCC Breakpoints | | | | WWS Breakpoints | | | |
|---|---|---|---|---|---|---|---|
| ID | Date | CBP | CBP SE | ID | Date | WBP | WBP SE |
| CB1 | 2021-04-15 | 33 | 1 | WB1 | 2021-04-26 | 44 | 3 |
| CB2 | 2021-06-03 | 82 | 1 | WB2 | 2021-07-11 | 120 | 3 |
| CB3 | 2021-07-11 | 120 | 5 | | | | |

[a] ID, is the code for the CCC and WWS breakpoints; Date, is the break-point date estimated by the segmented routine; CBP and WBP, refer to the CCC and WWS break points, respectively; CBP SE and WBP SE, refer to the SE of the CCC and WWS break points.

**Table S4.** The CCC and WWS trends and associated standard errors (SE)[a]

| CCC Trend Lines | | | | WWS Trend Lines | | | |
|---|---|---|---|---|---|---|---|
| ID | n | DC | DC SE | ID | n | PDC | PDC SE |
| CS1 | 32 | 11.7 | 0.5 | WS1 | 26 | 3.6 | 0.8 |
| CS2 | 49 | -10.2 | 0.3 | WS2 | 32 | -5.7 | 0.4 |
| CS3 | 38 | -1.0 | 0.4 | WS3 | 31 | 4.0 | 0.6 |
| CS4 | 49 | 1.5 | 0.3 | | | | |

[a] ID, is the identification code for the CCC and WWS trend lines; n, is the number of data points within each trend interval; DC, is the daily change or slope in cases per day for the CCC; PDC, is the percent daily change for the WWS or the slope of the trend line within each interval; DC SE and PCD SE, refer to the standard error of the DC and PDC, respectively and correlate to the error of the slope of the linear trend line within each interval.

### 2. TAB Longer Term QTA Based on the Median PMMoV-Flow Normalized N1 Viral Load ($L_{vb}$) and CCC by Reported Date

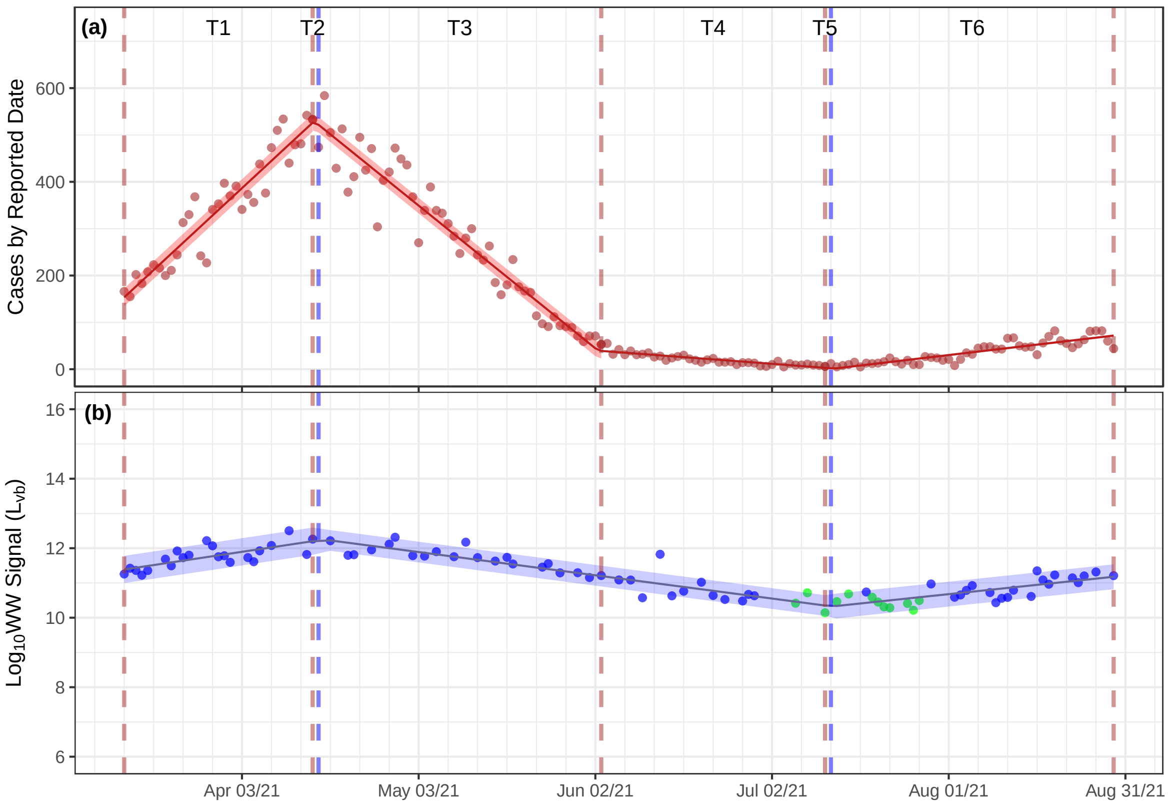


**Figure S2.** The TAB sewershed QTA of CCC by reported date (a) and $\log_{10}$ PMMoV-flow normalized N1 ($L_{vb}$) viral load (b).

**Table S5.** Summary aggregated trend results for the TAB WWTP sewershed[a]

| Interval | Trend Duration | | Trends in CCC | | Trends in WW | |
|---|---|---|---|---|---|---|
| ID | Start | End | DC (CI) | | PDC (CI) | |
| T1 | Mar 14/21 | Apr 15/21 | 12 ( 11 – 13 ) | ↑ | 6 ( 4 – 9 ) | ↑ |
| T2 | Apr 15/21 | Apr 16/21 | -10 ( -11 – -10 ) | ↓ | 6 ( 4 – 9 ) | ↑ |
| T3 | Apr 16/21 | Jun 03/21 | -10 ( -11 – -10 ) | ↓ | -5 ( -6 – -4 ) | ↓ |
| T4 | Jun 03/21 | Jul 11/21 | -1 ( -1.7 – -0.2 ) | ↓ | -5 ( -6 – -4 ) | ↓ |
| T5 | Jul 11/21 | Jul 12/21 | 1.5 ( 0.9 – 2.1 ) | ↑ | -5 ( -6 – -4 ) | ↓ |
| T6 | Jul 12/21 | Aug 29/21 | 1.5 ( 0.9 – 2.1 ) | ↑ | 4 ( 3 – 6 ) | ↑ |

[a] DC and PDC, refer to daily change and percent daily change, respectively; CI, is the 95 % confidence interval.

**Table S6.** Summary interpretation of trend results for the TAB WWTP sewershed

| | Signal Trends | | Trends Interpretation |
|---|---|---|---|
| Interval | CCC | WWS | Integration of CCC and WWS Signals |
| T1 | ↑ | ↑ | **Both** the CCC and WWS **are trending up**. Strong evidence of escalating community transmission. |
| T2 | ↓ | ↑ | The CCC is **trending down** and the WWS is **trending up**. Weak evidence of escalating community transmission. |
| T3 | ↓ | ↓ | **Both** the CCC and WWS signals are **trending down**. No evidence of escalating community transmission. |
| T4 | ↓ | ↓ | **Both** the CCC and WWS signals are **trending down**. No evidence of escalating community transmission. |
| T5 | ↑ | ↓ | The CCC **trending up** and the WWS is **trending down**. Weak evidence of escalating community transmission. |
| T6 | ↑ | ↑ | **Both** the CCC and WWS **are trending up**. Strong evidence of escalating community transmission. |

**Table S7.** The CCC and WWS breakpoints (CBP and WBP) and associated SE[a]

| CCC Breakpoints | | | | WWS Breakpoints | | | |
|---|---|---|---|---|---|---|---|
| ID | Date | CBP | CBP SE | ID | Date | WBP | WBP SE |
| CB1 | 2021-04-15 | 33 | 1 | WB1 | 2021-04-16 | 35 | 3 |
| CB2 | 2021-06-03 | 82 | 1 | WB2 | 2021-07-12 | 121 | 3 |
| CB3 | 2021-07-11 | 120 | 5 | | | | |

[a] ID, is the code for the CCC and WWS breakpoints; Date, is the break-point date estimated by the segmented routine; CBP and WBP, refer to the CCC and WWS break points, respectively; CBP SE and WBP SE, refer to the SE of the CCC and WWS break points.

**Table S8.** The CCC and WWS trends and associated standard errors (SE)[a]

| CCC Trend Lines | | | | WWS Trend Lines | | | |
|---|---|---|---|---|---|---|---|
| ID | n | DC | DC SE | ID | n | PDC | PDC SE |
| CS1 | 32 | 11.7 | 0.5 | WS1 | 22 | 6.0 | 1.2 |
| CS2 | 49 | -10.2 | 0.3 | WS2 | 36 | -5.0 | 0.4 |
| CS3 | 38 | -1.0 | 0.4 | WS3 | 31 | 4.2 | 0.7 |
| CS4 | 49 | 1.5 | 0.3 | | | | |

[a] ID, is the identification code for the CCC and WWS trend lines; n, is the number of data points within each trend interval; DC, is the daily change or slope in cases per day for the CCC; PDC, is the percent daily change for the WWS or the slope of the trend line within each interval; DC SE and PCD SE, refer to the standard error of the DC and PDC, respectively and correlate to the error of the slope of the linear trend line within each interval.

### 3. TAB Longer Term QTA Based on the Median PMMoV-Flow Normalized N1 Viral Load ($L_{vb}$) and CCC by Episode Date

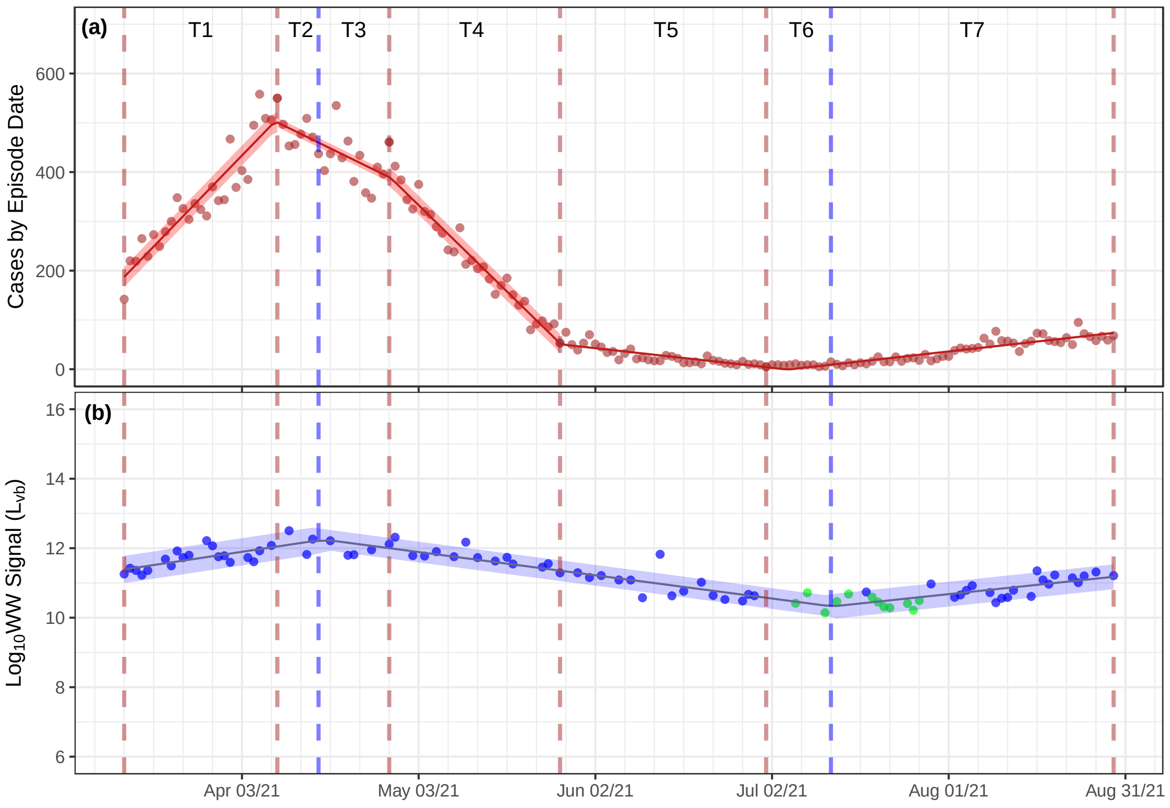


**Figure S3.** The TAB sewershed QTA of CCC by reported date (a) and $\log_{10}$ median PMMoV-flow normalized N1 ($Lvb$) viral loading (b)

**Table S9.** Summary aggregated trend results for the TAB WWTP sewershed[a]

| Interval | Trend Duration | | Trends in CCC | | Trends in WW | |
|---|---|---|---|---|---|---|
| ID | Start | End | DC (CI) | | PDC (CI) | |
| T1 | Mar 14/21 | Apr 09/21 | 12 ( 11 – 14 ) | ↑ | 6 ( 4 – 9 ) | ↑ |
| T2 | Apr 09/21 | Apr 16/21 | -6 ( -8 – -4 ) | ↓ | 6 ( 4 – 9 ) | ↑ |
| T3 | Apr 16/21 | Apr 28/21 | -6 ( -8 – -4 ) | ↓ | -5 ( -6 – -4 ) | ↓ |
| T4 | Apr 28/21 | May 27/21 | -12 ( -13 – -11 ) | ↓ | -5 ( -6 – -4 ) | ↓ |
| T5 | May 27/21 | Jul 01/21 | -1 ( -2.0 – -0.6 ) | ↓ | -5 ( -6 – -4 ) | ↓ |
| T6 | Jul 01/21 | Jul 12/21 | 1.3 ( 1.0 – 1.7 ) | ↑ | -5 ( -6 – -4 ) | ↓ |
| T7 | Jul 12/21 | Aug 29/21 | 1.3 ( 1.0 – 1.7 ) | ↑ | 4 ( 3 – 6 ) | ↑ |

[a] DC and PDC, refer to daily change and percent daily change, respectively; CI, is the 95 % confidence interval.

**Table S10.** Summary interpretation of trend results for the TAB WWTP sewershed

| | Signal Trends | | Trends Interpretation |
|---|---|---|---|
| Interval | CCC | WWS | Integration of CCC and WWS Signals |
| T1 | ↑ | ↑ | **Both** the CCC and WWS **are trending up**. Strong evidence of escalating community transmission. |
| T2 | ↓ | ↑ | The CCC is **trending down** and the WWS is **trending up**. Weak evidence of escalating community transmission. |
| T3 | ↓ | ↓ | **Both** the CCC and WWS signals are **trending down**. No evidence of escalating community transmission. |
| T4 | ↓ | ↓ | **Both** the CCC and WWS signals are **trending down**. No evidence of escalating community transmission. |
| T5 | ↓ | ↓ | **Both** the CCC and WWS signals are **trending down**. No evidence of escalating community transmission. |
| T6 | ↑ | ↓ | The CCC **trending up** and the WWS is **trending down**. Weak evidence of escalating community transmission. |
| T7 | ↑ | ↑ | **Both** the CCC and WWS **are trending up**. Strong evidence of escalating community transmission. |

**Table S11.** The CCC and WWS breakpoints (CBP and WBP) and associated SE[a]

| CCC Breakpoints | | | | WWS Breakpoints | | | |
|---|---|---|---|---|---|---|---|
| ID | Date | CBP | CBP SE | ID | Date | WBP | WBP SE |
| CB1 | 2021-04-09 | 27 | 1 | WB1 | 2021-04-16 | 35 | 3 |
| CB2 | 2021-04-28 | 46 | 2 | WB2 | 2021-07-12 | 121 | 3 |
| CB3 | 2021-05-27 | 75 | 1 | | | | |
| CB4 | 2021-07-01 | 110 | 4 | | | | |

[a] ID, is the code for the CCC and WWS breakpoints; Date, is the break-point date estimated by the segmented routine; CBP and WBP, refer to the CCC and WWS break points, respectively; CBP SE and WBP SE, refer to the SE of the CCC and WWS break points.

**Table S12.** The CCC and WWS trends and associated standard errors (SE)[a]

| CCC Trend Lines | | | | WWS Trend Lines | | | |
|---|---|---|---|---|---|---|---|
| ID | n | DC | DC SE | ID | n | PDC | PDC SE |
| CS1 | 26 | 12.3 | 0.6 | WS1 | 22 | 6.0 | 1.2 |
| CS2 | 19 | -5.8 | 0.9 | WS2 | 36 | -5.0 | 0.4 |
| CS3 | 29 | -11.7 | 0.5 | WS3 | 31 | 4.2 | 0.7 |
| CS4 | 35 | -1.3 | 0.4 | | | | |
| CS5 | 59 | 1.3 | 0.2 | | | | |

[a] ID, is the identification code for the CCC and WWS trend lines; n, is the number of data points within each trend interval; DC, is the daily change or slope in cases per day for the CCC; PDC, is the percent daily change for the WWS or the slope of the trend line within each interval; DC SE and PCD SE, refer to the standard error of the DC and PDC, respectively and correlate to the error of the slope of the linear trend line within each interval.

### 4. Toronto Humber River (THR) Shorter Term QTA Based on PMMoV-normalized N1 Concentration ($C_{vb}$) and CCC by Reported Date

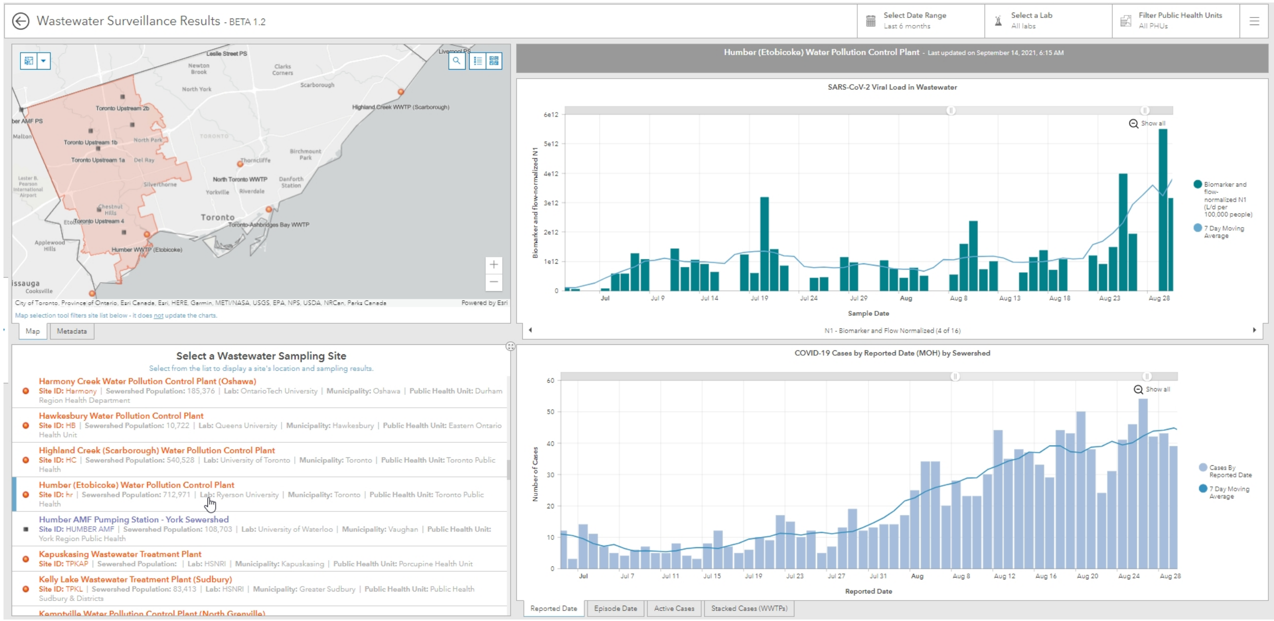


**Figure S4.** Toronto Humber River sewershed (THR) with PMMoV-normalized N1 concentration ($C_{vb}$) and clinical case data from the Ontario Dashboard

**Table S13.** Summary aggregated trend results for the TAB WWTP sewershed[a]

| Interval | Trend Duration | | Trends in CCC | | Trends in WW | |
|---|---|---|---|---|---|---|
| ID | Start | End | DC (CI) | | PDC (CI) | |
| T1 | Jul 28/21 | Aug 20/21 | 1 ( 1.0 – 1.8 ) | ⬆ | 2 ( -1.0 – 4 ) | ⬆ |
| T2 | Aug 20/21 | Aug 23/21 | -4 ( -10.1 – 1.5 ) | ⬇ | 2 ( -1.0 – 4 ) | ⬆ |
| T3 | Aug 23/21 | Aug 25/21 | 11 ( -7.1 – 29.5 ) | ⬆ | 168 ( -20 – 801 ) | ⬆ |

[a] DC and PDC, refer to daily change and percent daily change, respectively; CI, is the 95 % confidence interval.

**Table S14.** Summary interpretation of trend results for the TAB WWTP sewershed

| | Signal Trends | | Trends Interpretation |
|---|---|---|---|
| Interval | CCC | WWS | Integration of CCC and WWS |
| T1 | ⬆ | ⬆ | **Both** the CCC and WWS **are trending up**. Strong evidence of escalating community transmission. |
| T2 | ⬇ | ⬆ | The CCC is **trending down** and the WWS is **trending up**. Weak evidence of escalating community transmission. |
| T3 | ⬆ | ⬆ | **Both** the CCC and WWS **are trending up**. Strong evidence of escalating community transmission. |

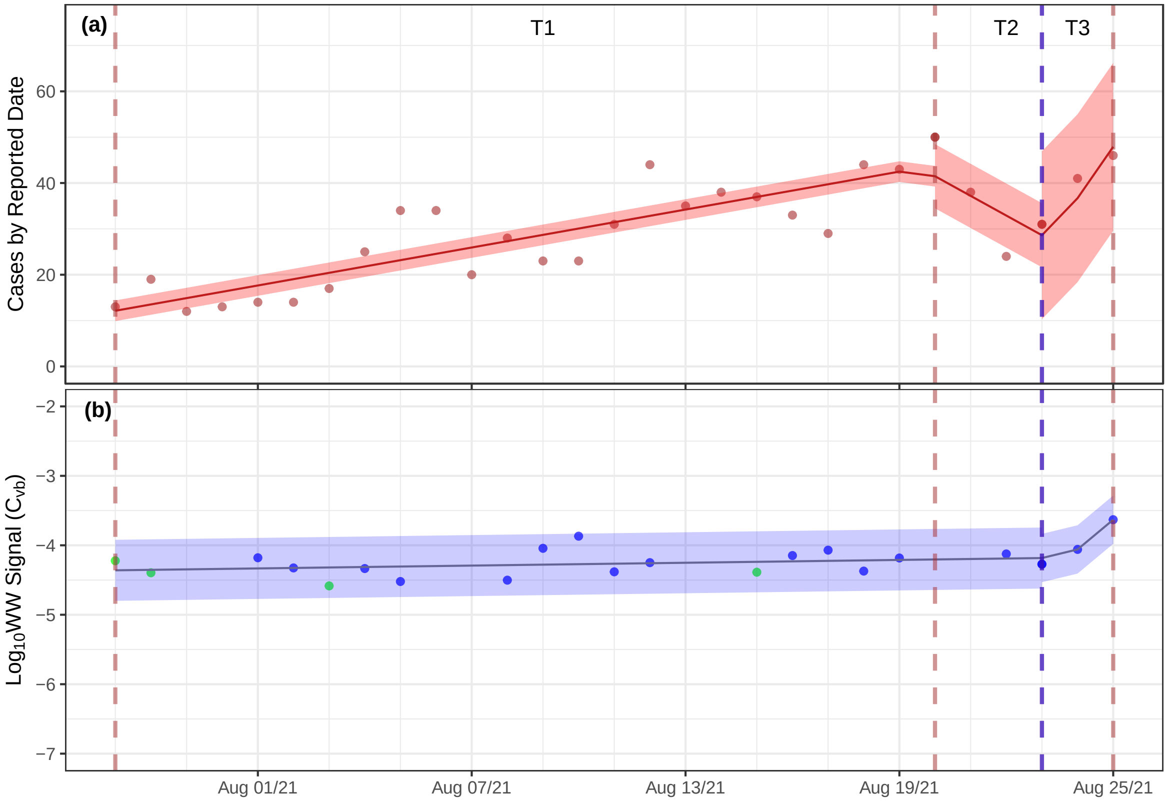


**Figure S5.** The THR sewershed QTA of CCC by reported date (a) and $\log_{10}$ PMMoV-normalized N1 ($C_{vb}$) viral concentration (b).

**Table S15.** The CCC and WWS breakpoints (CBP and WBP) and associated SE[a]

| CCC Breakpoints | | | | WWS Breakpoints | | | |
|---|---|---|---|---|---|---|---|
| ID | Date | CBP | CBP SE | ID | Date | WBP | WBP SE |
| CB1 | 2021-08-20 | 24 | 1 | WB1 | 2021-08-23 | 28 | 1 |
| CB2 | 2021-08-23 | 27 | 1 | | | | |

[a] ID, is the code for the CCC and WWS breakpoints; Date, is the break-point date estimated by the segmented routine; CBP and WBP, refer to the CCC and WWS break points, respectively; CBP SE and WBP SE, refer to the SE of the CCC and WWS break points.

**Table S16.** The CCC and WWS trends and associated standard errors (SE)[a]

| CCC Trend Lines | | | | WWS Trend Lines | | | |
|---|---|---|---|---|---|---|---|
| ID | n | DC | DC SE | ID | n | PDC | PDC SE |
| CS1 | 23 | 1.4 | 0.2 | WS1 | 20 | 1.6 | 1.2 |
| CS2 | 3 | -4.3 | 2.8 | WS2 | 2 | 168.0 | 77.7 |
| CS3 | 2 | 11.2 | 8.8 | | | | |

[a] ID, is the identification code for the CCC and WWS trend lines; n, is the number of data points within each trend interval; DC, is the daily change or slope in cases per day for the CCC; PDC, is the percent daily change for the WWS or the slope of the trend line within each interval; DC SE and PCD SE, refer to the standard error of the DC and PDC, respectively and correlate to the error of the slope of the linear trend line within each interval.

### 5. THR Shorter Term Flow-Normalized N1 viral loading ($L_v$) and CCC by Reported Date

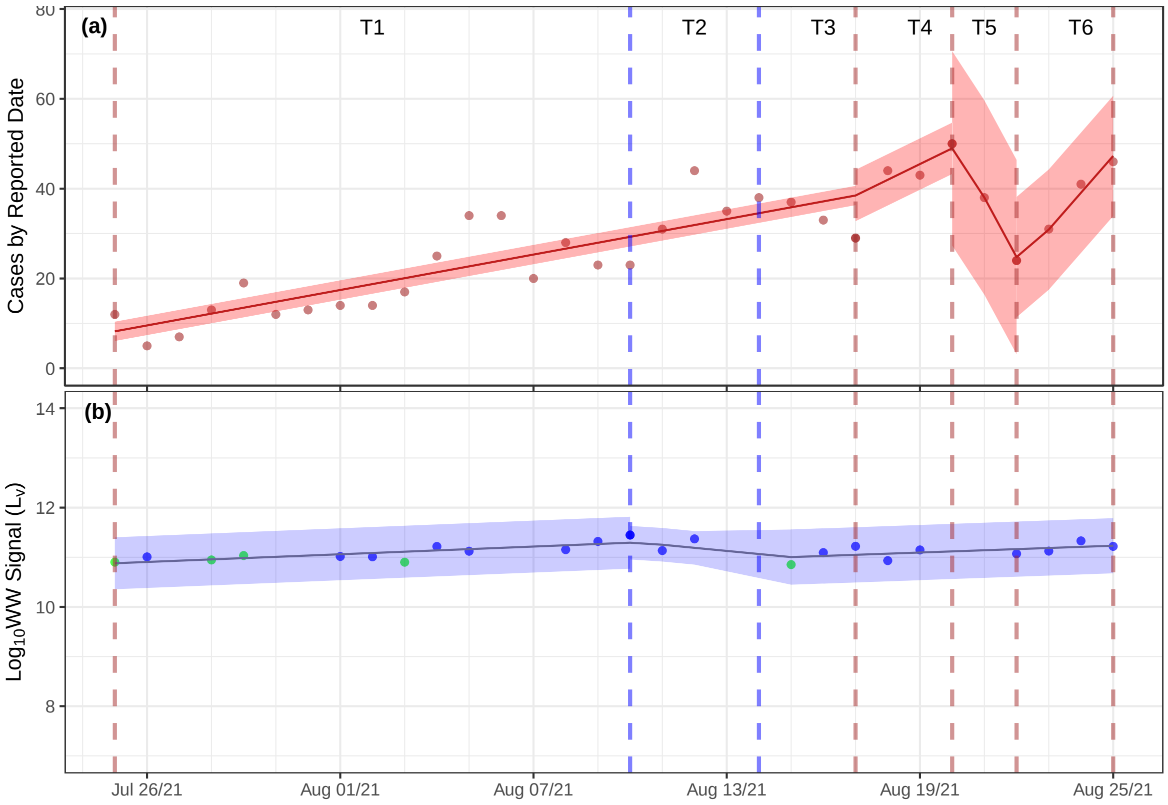


**Figure S6.** The THR sewershed QTA of CCC by reported date (a) and $\log_{10}$ Flow-normalized N1 ($L_v$) viral loading (b)

**Table S17.** Summary aggregated trend results for the THR WWTP sewershed[a]

| Interval | Trend Duration | | Trends in CCC | | Trends in WW | |
|---|---|---|---|---|---|---|
| ID | Start | End | DC (CI) | | PDC (CI) | |
| T1 | Jul 25/21 | Aug 10/21 | 1 ( 1.0 – 1.7 ) | ↑ | 6 ( 3 – 10 ) | ↑ |
| T2 | Aug 10/21 | Aug 14/21 | 1 ( 1.0 – 1.7 ) | ↑ | -13 ( -63 – 103 ) | ↓ |
| T3 | Aug 14/21 | Aug 17/21 | 1 ( 1.0 – 1.7 ) | ↑ | 5 ( -0.5 – 12 ) | ↑ |
| T4 | Aug 17/21 | Aug 20/21 | 4 ( -4.7 – 11.7 ) | ↑ | 5 ( -0.5 – 12 ) | ↑ |
| T5 | Aug 20/21 | Aug 22/21 | -13 ( -29.7 – 3.2 ) | ↓ | 5 ( -0.5 – 12 ) | ↑ |
| T6 | Aug 22/21 | Aug 25/21 | 8 ( -0.0 – 16.4 ) | ↑ | 5 ( -0.5 – 12 ) | ↑ |

[a] DC and PDC, refer to daily change and percent daily change, respectively; CI, is the 95 % confidence interval.

**Table S18.** Summary interpretation of trend results for the THR WWTP sewershed

| | Signal Trends | | Trends Interpretation |
|---|---|---|---|
| Interval | CCC | WWS | Integration of CCC and WWS Signals |
| T1 | ⬆ | ⬆ | **Both** the CCC and WWS **are trending up**. Strong evidence of escalating community transmission. |
| T2 | ⬆ | ⬇ | The CCC **trending up** and the WWS is **trending down**. Weak evidence of escalating community transmission. |
| T3 | ⬆ | ⬆ | **Both** the CCC and WWS **are trending up**. Strong evidence of escalating community transmission. |
| T4 | ⬆ | ⬆ | **Both** the CCC and WWS **are trending up**. Strong evidence of escalating community transmission. |
| T5 | ⬇ | ⬆ | The CCC is **trending down** and the WWS is **trending up**. Weak evidence of escalating community transmission. |
| T6 | ⬆ | ⬆ | **Both** the CCC and WWS **are trending up**. Strong evidence of escalating community transmission. |

**Table S19.** The CCC and WWS breakpoints (CBP and WBP) and associated SE[a]

| CCC Breakpoints | | | | WWS Breakpoints | | | |
|---|---|---|---|---|---|---|---|
| ID | Date | CBP | CBP SE | ID | Date | WBP | WBP SE |
| CB1 | 2021-08-17 | 24 | 4 | WB1 | 2021-08-10 | 17 | 3 |
| CB2 | 2021-08-20 | 27 | 1 | WB2 | 2021-08-14 | 22 | 7 |
| CB3 | 2021-08-22 | 29 | 0 | | | | |

[a] ID, is the code for the CCC and WWS breakpoints; Date, is the break-point date estimated by the segmented routine; CBP and WBP, refer to the CCC and WWS break points, respectively; CBP SE and WBP SE, refer to the SE of the CCC and WWS break points.

**Table S20.** The CCC and WWS trends and associated standard errors (SE)[a]

| CCC Trend Lines | | | | WWS Trend Lines | | | |
|---|---|---|---|---|---|---|---|
| ID | n | DC | DC SE | ID | n | PDC | PDC SE |
| CS1 | 23 | 1.3 | 0.2 | WS1 | 12 | 6.1 | 1.6 |
| CS2 | 3 | 3.5 | 4.0 | WS2 | 4 | -13.3 | 49.8 |
| CS3 | 2 | -13.2 | 8.0 | WS3 | 9 | 5.4 | 2.8 |
| CS4 | 3 | 8.2 | 4.0 | | | | |

[a] ID, is the identification for the CCC and WWS trend; n, is the data points within each trend interval; DC, is the daily change or slope in cases per day; PDC, is the percent daily change for the WWS within each interval; DC SE and PCD SE, refer to the standard error of the DC and PDC and correlate to the error of the slope of the linear trend lines.

### 6. THR Shorter Term Median PMMoV-Flow Normalized N1 Viral Load ($L_{vb}$) and CCC by Reported Date

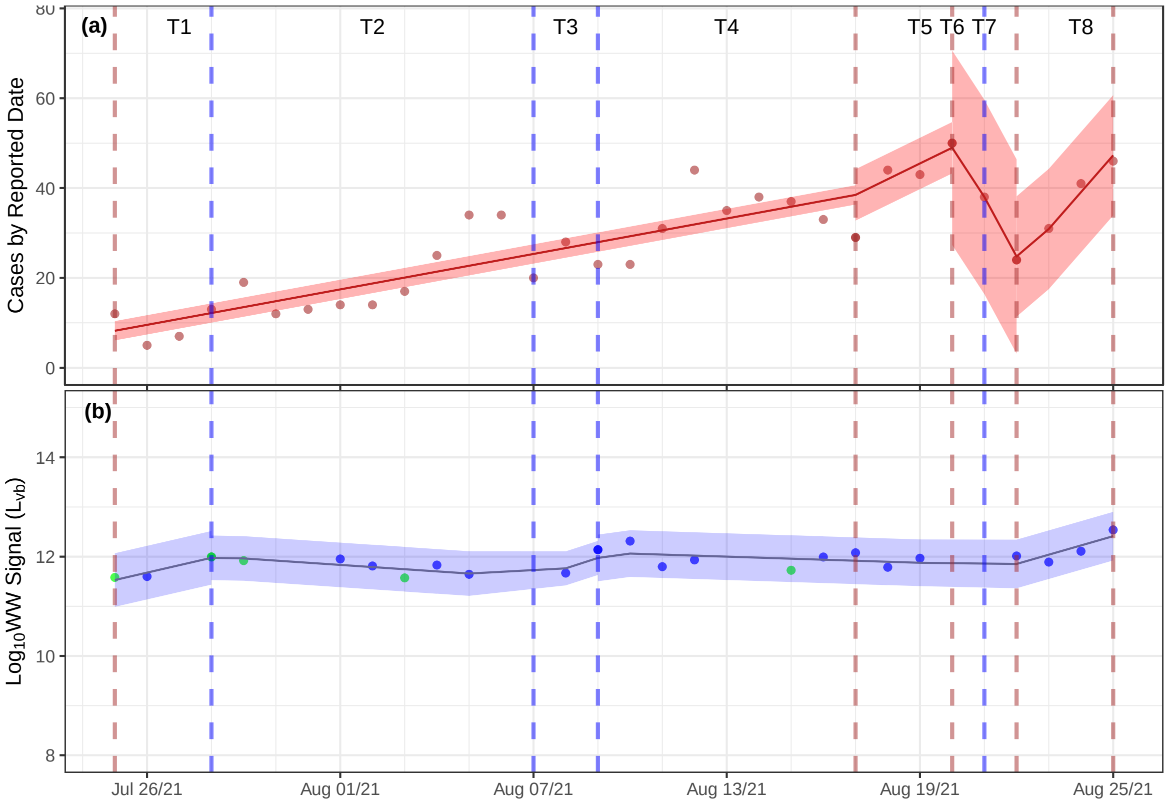


**Figure S7.** The TAB sewershed QTA of CCC by reported date (a) and $\log_{10}$ median PMMoV-flow normalized N1 ($C_{vb}$) viral load (b).

**Table S21.** Summary aggregated trend results for the TAB WWTP sewershed[a]

| Interval | Trend Duration | | Trends in CCC | | Trends in WW | |
|---|---|---|---|---|---|---|
| ID | Start | End | DC (CI) | | PDC (CI) | |
| T1 | Jul 25/21 | Jul 28/21 | 1 ( 1.0 – 1.7 ) | ⬆ | 41 ( -7 – 113 ) | ⬆ |
| T2 | Jul 28/21 | Aug 07/21 | 1 ( 1.0 – 1.7 ) | ⬆ | -10 ( -23 – 6 ) | ⬇ |
| T3 | Aug 07/21 | Aug 09/21 | 1 ( 1.0 – 1.7 ) | ⬆ | 62 ( -54 – 475 ) | ⬆ |
| T4 | Aug 09/21 | Aug 17/21 | 1 ( 1.0 – 1.7 ) | ⬆ | -5 ( -14 – 5 ) | ⬇ |
| T5 | Aug 17/21 | Aug 20/21 | 4 ( -4.7 – 11.7 ) | ⬆ | -5 ( -14 – 5 ) | ⬇ |
| T6 | Aug 20/21 | Aug 21/21 | -13 ( -29.7 – 3.2 ) | ⬇ | -5 ( -14 – 5 ) | ⬇ |
| T7 | Aug 21/21 | Aug 22/21 | -13 ( -29.7 – 3.2 ) | ⬇ | 54 ( 3 – 129 ) | ⬆ |
| T8 | Aug 22/21 | Aug 25/21 | 8 ( -0.0 – 16.4 ) | ⬆ | 54 ( 3 – 129 ) | ⬆ |

[a] DC and PDC, refer to daily change and percent daily change, respectively; CI, is the 95 % confidence interval.

**Table S22.** Summary interpretation of trend results for the TAB WWTP sewershed

| | Signal Trends | | Trends Interpretation |
|---|---|---|---|
| Interval | CCC | WWS | Integration of CCC and WWS Signals |
| T1 | ⬆ | ⬆ | **Both** the CCC and WWS **are trending up**. Strong evidence of escalating community transmission. |
| T2 | ⬆ | ⬇ | The CCC **trending up** and the WWS is **trending down**. Weak evidence of escalating community transmission. |
| T3 | ⬆ | ⬆ | **Both** the CCC and WWS **are trending up**. Strong evidence of escalating community transmission. |
| T4 | ⬆ | ⬇ | The CCC **trending up** and the WWS is **trending down**. Weak evidence of escalating community transmission. |
| T5 | ⬆ | ⬇ | The CCC **trending up** and the WWS is **trending down**. Weak evidence of escalating community transmission. |
| T6 | ⬇ | ⬇ | **Both** the CCC and WWS signals are **trending down**. No evidence of escalating community transmission. |
| T7 | ⬇ | ⬆ | The CCC is **trending down** and the WWS is **trending up**. Weak evidence of escalating community transmission. |
| T8 | ⬆ | ⬆ | **Both** the CCC and WWS **are trending up**. Strong evidence of escalating community transmission. |

**Table S23.** The CCC and WWS breakpoints (CBP and WBP) and associated SE[a]

| CCC Breakpoints | | | | WWS Breakpoints | | | |
|---|---|---|---|---|---|---|---|
| ID | Date | CBP | CBP SE | ID | Date | WBP | WBP SE |
| CB1 | 2021-08-17 | 24 | 4 | WB1 | 2021-07-28 | 4 | 1 |
| CB2 | 2021-08-20 | 27 | 1 | WB2 | 2021-08-07 | 14 | 2 |
| CB3 | 2021-08-22 | 29 | 0 | WB3 | 2021-08-09 | 16 | 1 |
| | | | | WB4 | 2021-08-21 | 29 | 1 |

[a] ID, is the code for the CCC and WWS breakpoints; Date, is the break-point date estimated by the segmented routine; CBP and WBP, refer to the CCC and WWS break points, respectively; CBP SE and WBP SE, refer to the SE of the CCC and WWS break points.

**Table S24.** The CCC and WWS trends and associated standard errors (SE)[a]

| CCC Trend Lines | | | | WWS Trend Lines | | | |
|---|---|---|---|---|---|---|---|
| ID | n | DC | DC SE | ID | n | PDC | PDC SE |
| CS1 | 23 | 1.3 | 0.2 | WS1 | 3 | 41.0 | 21.1 |
| CS2 | 3 | 3.5 | 4.0 | WS2 | 7 | -9.5 | 7.7 |
| CS3 | 2 | -13.2 | 8.0 | WS3 | 2 | 62.2 | 79.6 |
| CS4 | 3 | 8.2 | 4.0 | WS4 | 10 | -4.6 | 4.8 |
| | | | | WS5 | 4 | 53.7 | 20.3 |

[a] ID, is the identification code for the CCC and WWS trend lines; n, is the number of data points within each trend interval; DC, is the daily change or slope in cases per day for the CCC; PDC, is the percent daily change for the WWS or the slope of the trend line within each interval; DC SE and PCD SE, refer to the standard error of the DC and PDC, respectively and correlate to the error of the slope of the linear trend line within each interval.

### 7. THR Linear Correlation of WWS to CCC by Reported Date

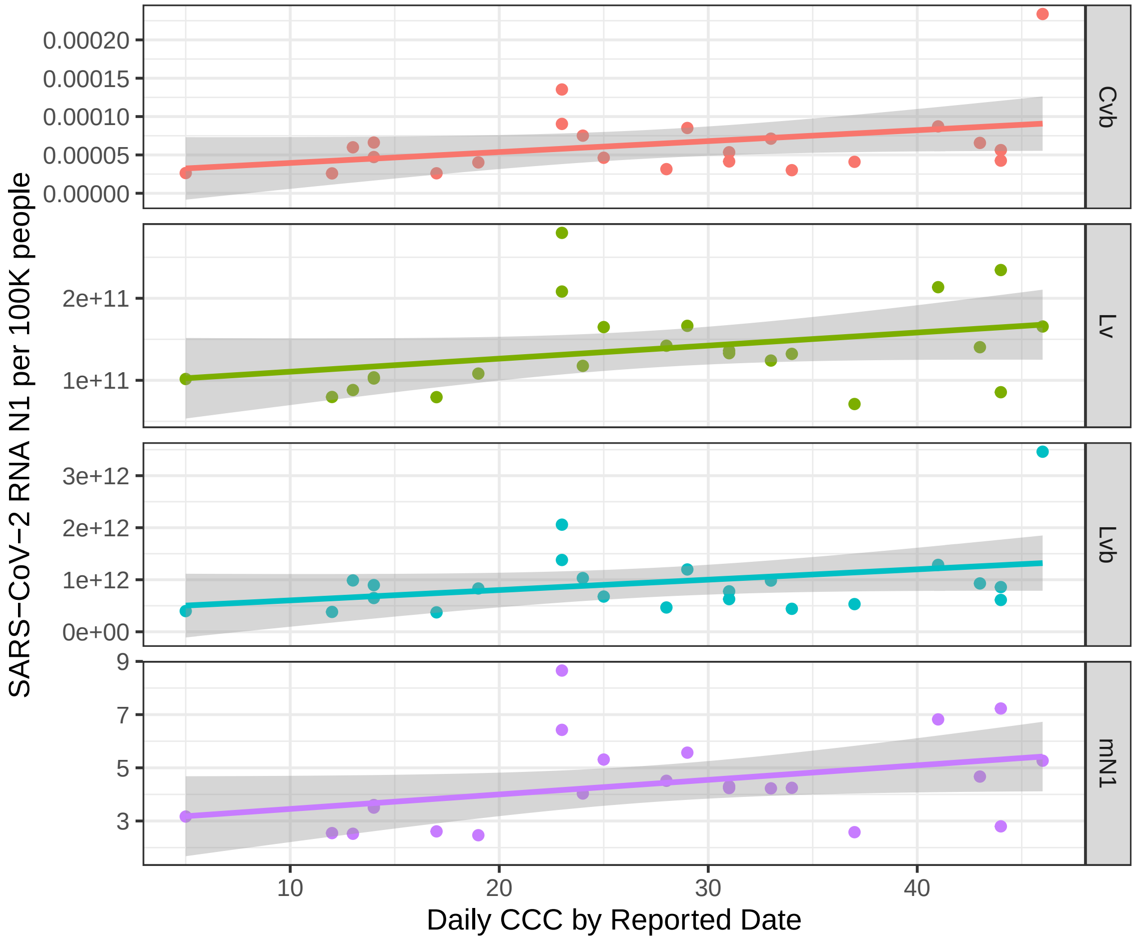


**Figure S8.** The THR sewershed linear correlation normalizations for N1 ($C_{vb}$, $L_{vb}$, $L_v$ and $mN1$) versus CCC by reported date with 95 % confidence bands.

**Table S25.** THR N1 WWS ($C_{vb}$, $L_v$, $L_{vb}$, mN1) versus CCC per 100,000 people correlation results

| | Linear Correlation Results | | | Nonlinear Correlation Results | | | |
|---|---|---|---|---|---|---|---|
| Normalization | Slope ± SE | $R^2$ | P-Value | AICc | AICcWt | $aR^2$ | PO |
| $C_{vb}$ | 1e-06 ± 8e-07 | 0.10 | 0.08 | -625 | 0.36 | 0.42 | 8 |
| $L_{vb}$ | 2e+10 ± 1e+10 | 0.08 | 0.10 | 2060 | 0.29 | 0.38 | 8 |
| $L_v$ | 2e+09 ± 9e+08 | 0.08 | 0.10 | 1899 | 0.68 | 0.04 | 1 |
| $mN1$ | 5e-02 ± 3e-02 | 0.11 | 0.07 | 154 | 0.63 | 0.11 | 1 |

[a] Slope ± SE, is the slope and standard error associated with the linear correlation regression; $R^2$, is the coefficient of determination; p-value, is the probability that the predictor (WVS) is related to the response (CCC) compared to the null hypothesis of being unrelated; AICc, is the Akaike information criterion; AICcWt, is the AICc weight; $aR^2$, is the adjusted $R^2$ and PO, is the polynomial order of the best fit curve up to order 9.

### 8. Toronto Highland Creek (THC) Shorter Term Median PMMoV-Flow N1 Viral Loading ($L_{vb}$) and CCC by Reported Date

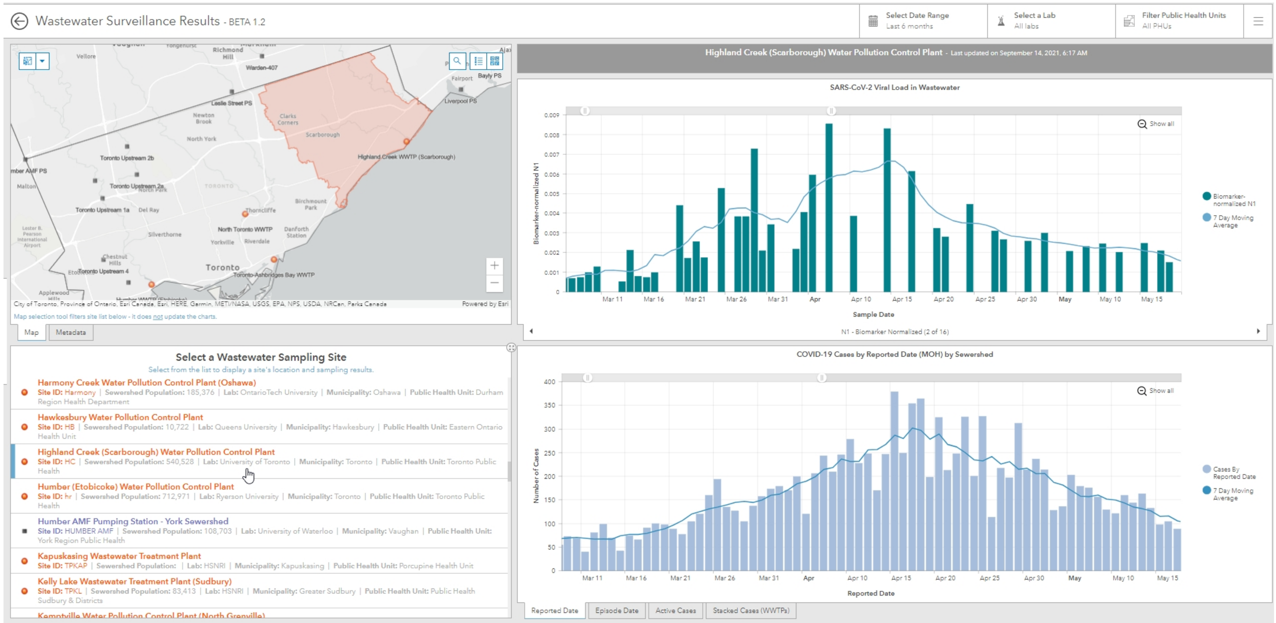


**Figure S9.** Toronto Highland Creek (THC) sewershed with viral load and clincal case data from the Ontario Dashboard

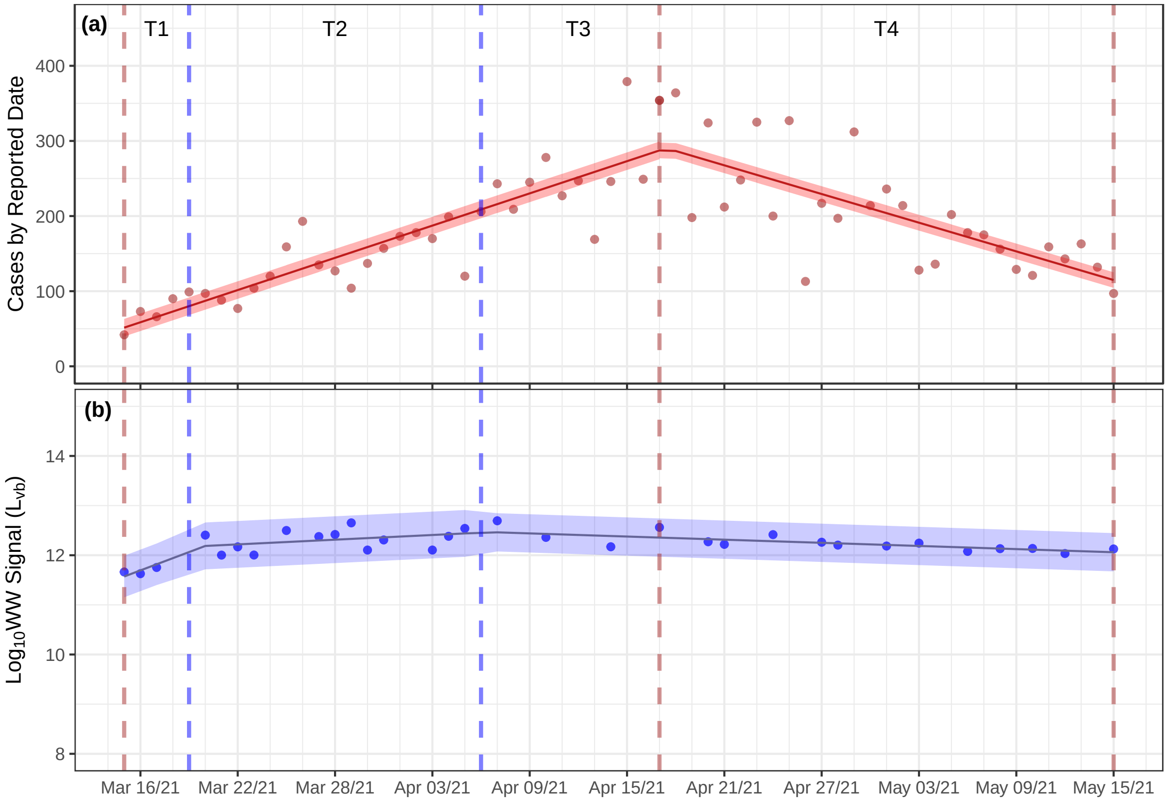


**Figure S10.** The THC sewershed QTA of CCC by reported date (a) and $L_{vb}$ viral loading (b)

**Table S26.** Summary aggregated trend results for the THC WWTP sewershed[a]

| Interval | Trend Duration | | Trends in CCC | | Trends in WW | |
|---|---|---|---|---|---|---|
| ID | Start | End | DC (CI) | | PDC (CI) | |
| T1 | Mar 15/21 | Mar 19/21 | 7 ( 6 – 9 ) | ↑ | 33 ( -23 – 130 ) | ↑ |
| T2 | Mar 19/21 | Apr 06/21 | 7 ( 6 – 9 ) | ↑ | 4 ( -0.5 – 8 ) | ↑ |
| T3 | Apr 06/21 | Apr 17/21 | 7 ( 6 – 9 ) | ↑ | -2 ( -4 – -1 ) | ↓ |
| T4 | Apr 17/21 | May 15/21 | -6 ( -8 – -4 ) | ↓ | -2 ( -4 – -1 ) | ↓ |

[a] DC and PDC, refer to daily change and percent daily change, respectively; CI, is the 95 % confidence interval.

**Table S27.** Summary interpretation of trend results for the TAB WWTP sewershed

| | Signal Trends | | Trends Interpretation |
|---|---|---|---|
| Interval | CCC | WWS | Integration of CCC and WWS Signals |
| T1 | ↑ | ↑ | **Both** the CCC and WWS **are trending up**. Strong evidence of escalating community transmission. |
| T2 | ↑ | ↑ | **Both** the CCC and WWS **are trending up**. Strong evidence of escalating community transmission. |
| T3 | ↑ | ↓ | The CCC **trending up** and the WWS is **trending down**. Weak evidence of escalating community transmission. |
| T4 | ↓ | ↓ | **Both** the CCC and WWS signals are **trending down**. No evidence of escalating community transmission. |

**Table S28.** The CCC and WWS breakpoints (CBP and WBP) and associated SE[a]

| CCC Breakpoints | | | | WWS Breakpoints | | | |
|---|---|---|---|---|---|---|---|
| ID | Date | CBP | CBP SE | ID | Date | WBP | WBP SE |
| CB1 | 2021-04-17 | 34 | 2 | WB1 | 2021-03-19 | 6 | 4 |
| | | | | WB2 | 2021-04-06 | 24 | 5 |

[a] ID, is the code for the CCC and WWS breakpoints; Date, is the break-point date estimated by the segmented routine; CBP and WBP, refer to the CCC and WWS break points, respectively; CBP SE and WBP SE, refer to the SE of the CCC and WWS break points.

**Table S29.** The CCC and WWS trends and associated standard errors (SE)[a]

| CCC Trend Lines | | | | WWS Trend Lines | | | |
|---|---|---|---|---|---|---|---|
| ID | n | DC | DC SE | ID | n | PDC | PDC SE |
| CS1 | 33 | 7.1 | 0.8 | WS1 | 4 | 32.8 | 30.6 |
| CS2 | 28 | -6.4 | 1.0 | WS2 | 14 | 3.7 | 2.0 |
| | | | | WS3 | 16 | -2.4 | 0.8 |

[a] ID, is the identification code for the CCC and WWS trend lines; n, is the number of data points within each trend interval; DC, is the daily change or slope in cases per day for the CCC; PDC, is the percent daily change for the WWS or the slope of the trend line within each interval; DC SE and PCD SE, refer to the standard error of the DC and PDC, respectively and correlate to the error of the slope of the linear trend line within each interval.

### 9. THC Shorter Term $L_{vb}$ and WCR with CCC by Reported Date

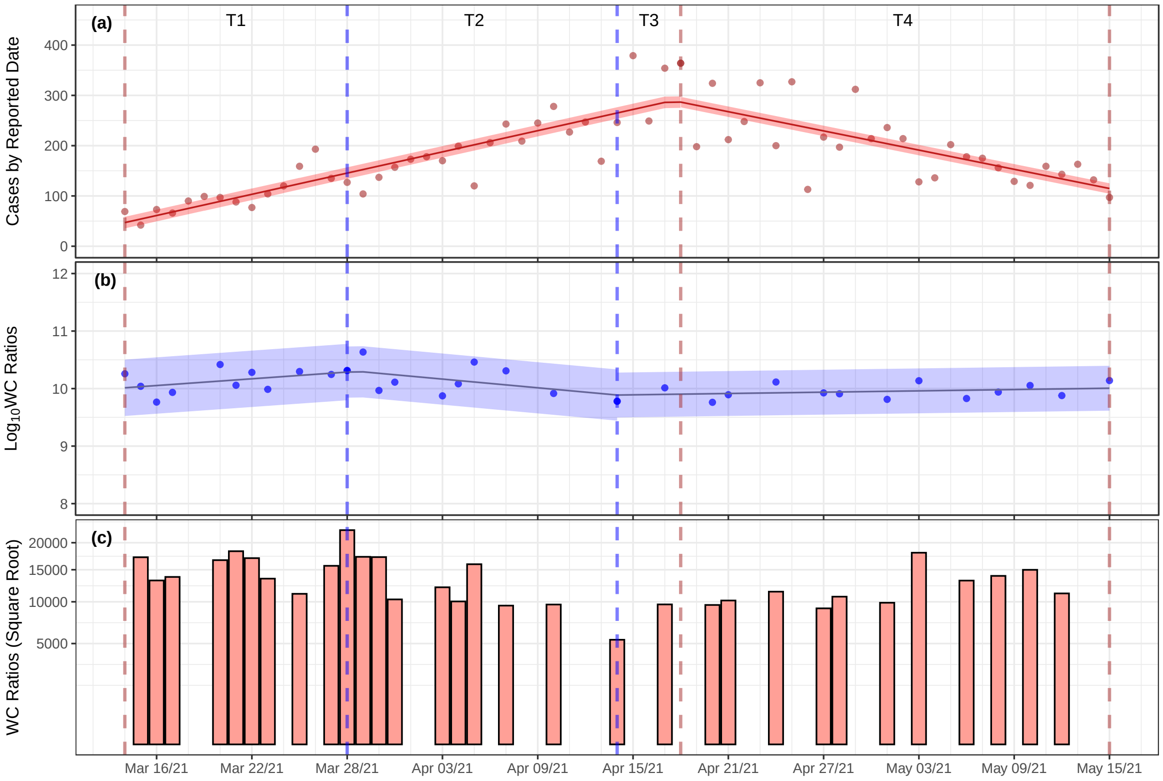


**Figure S11.** The THC sewershed trend analysis of CCC by reported date (a) and $\log_{10}$ $L_{vb}$ WC trend analysis (b) and WC values (c).

**Table S30.** Summary aggregated trend results for the TAB WWTP sewershed

| Interval | Trend Duration | | Trends in CCC | | Trends in WCR | |
|---|---|---|---|---|---|---|
| ID | Start | End | DC (CI) | | PDC (CI) | |
| T1 | Mar 14/21 | Mar 28/21 | 7 ( 6 – 9 ) | ⬆ | 5 ( -1 – 11 ) | ⬆ |
| T2 | Mar 28/21 | Apr 14/21 | 7 ( 6 – 9 ) | ⬆ | -6 ( -11 – 0 ) | ⬇ |
| T3 | Apr 14/21 | Apr 18/21 | 7 ( 6 – 9 ) | ⬆ | 1 ( -2 – 4 ) | ⬆ |
| T4 | Apr 18/21 | May 15/21 | -6 ( -8 – -4 ) | ⬇ | 1 ( -2 – 4 ) | ⬆ |

**Table S31.** Summary interpretation of trend results for the TAB WWTP sewershed CCC and WCR

| | Signal Trends | | Trends Interpretation |
|---|---|---|---|
| Interval | CCC | WCR | Integration of CCC and WCR |
| T1 | ↑ | ↑ | **Both** the CCC and WCR **are trending up**. Strong evidence of escalating community transmission and increase in asymptomatic cases in the community. |
| T2 | ↑ | ↓ | The CCC **trending up** and the WCR is **trending down**. Weak evidence of escalating community transmission and clinical testing may be over-estimating disease incidence when counting previously infected cases as new cases. |
| T3 | ↑ | ↑ | **Both** the CCC and WCR **are trending up**. Strong evidence of escalating community transmission and increase in asymptomatic cases in the community. |
| T4 | ↓ | ↑ | The CCC is **trending down** and the WCR is **trending up**. Weak evidence of escalating community transmission and increase in asymptomatic cases in the community. |

**Table S32.** The CCC and WWS estimated breakpoints and associated standard errors[a]

| CCC Breakpoints | | | | WCR Breakpoints | | | |
|---|---|---|---|---|---|---|---|
| ID | Date | CBP | CBP SE | ID | Date | WCBP | WCBP SE |
| CB1 | 2021-04-18 | 36 | 2 | WB1 | 2021-03-28 | 16 | 4 |
| | | | | WB2 | 2021-04-14 | 32 | 6 |

[a] ID, is the code for the CCC and WC breakpoints; Date, is the breakpoint date estimated by the segmented routine; CBP and WCBP, refer to the CSS and WC breakpoints; CBP SE and WCBP SE, refer to the SE of the CSB and WCBP, within each interval.

**Table S33.** The CCC and WCR estimated trend lines slopes and associated standard errors[a]

| CCC Trend Lines | | | | WCR Trend Lines | | | |
|---|---|---|---|---|---|---|---|
| ID | n | DC | DC SE | ID | n | PDWC | PDWC SE |
| CS1 | 36 | 7.0 | 0.7 | WS1 | 12 | 4.6 | 3.0 |
| CS2 | 27 | -6.4 | 1.0 | WS2 | 9 | -5.6 | 3.0 |
| | | | | WS3 | 14 | 0.9 | 1.4 |

[a] ID, is the code for the CCC and WCR trend lines; n, is the number of data points within each interval; DC and DWC, are the daily change in the CSS and percent daily change (PDC) of the WCR of the trend line (or slope) within each interval; DC SE, and PWDC SE, refer to the standard error in the corresponding units of the DC and PDWC.

### 10. THC Linear Correlation of WWS to CCC by Reported Date

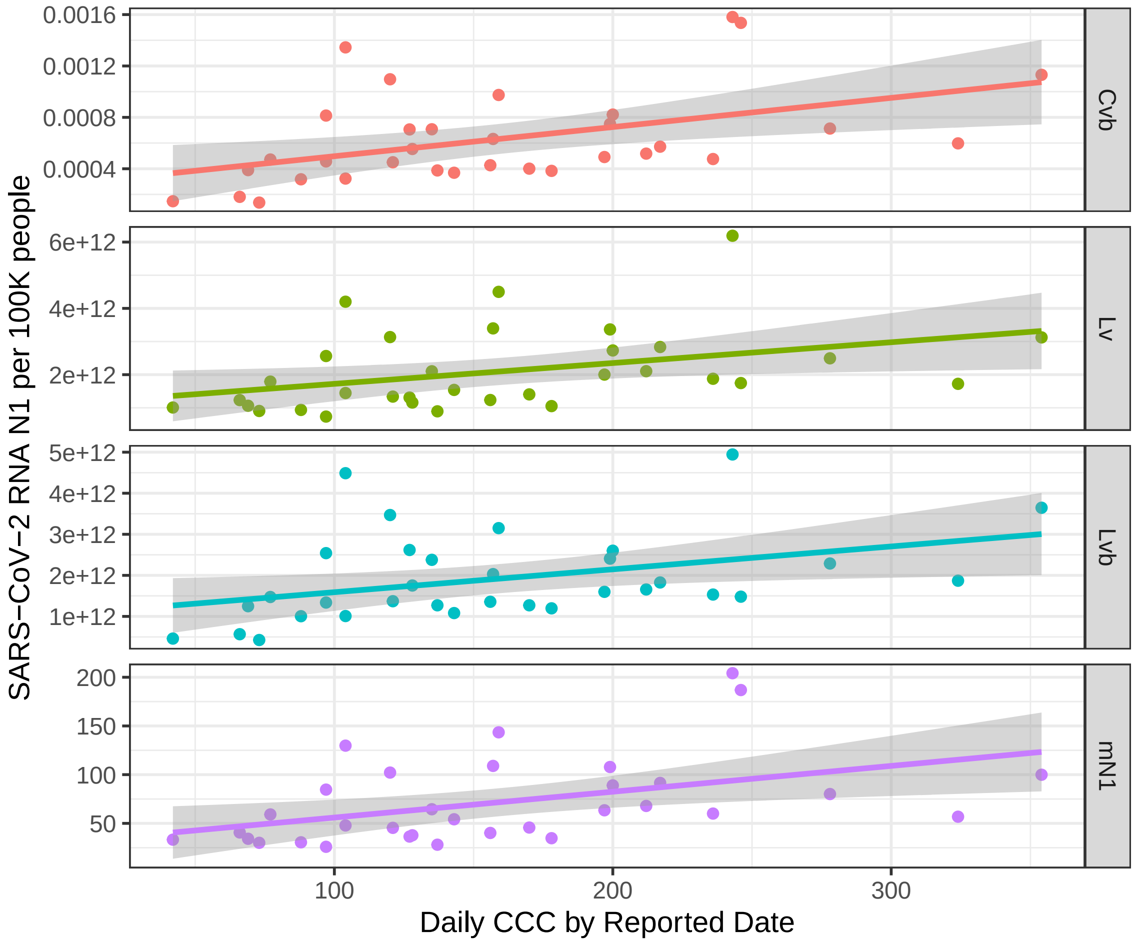


**Figure S12.** The THC sewershed linear correlation normalizations for N1 ($C_{vb}$, $L_{vb}$, $L_v$ and $mN1$) versus CCC by reported date with 95% confidence bands.

**Table S34.** THC N1 WWS ($C_{vb}$, $L_v$, $L_{vb}$, mN1) versus the daily CCC per 100,000 people correlation results

| | Linear Correlation Results | | | Nonlinear Correlation Results | | | |
|---|---|---|---|---|---|---|---|
| Normalization | Slope ± SE | $R^2$ | P-Value | AICc | AICcWt | $aR^2$ | PO |
| $C_{vb}$ | 2e-06 ± 8e-07 | 0.19 | 0.01 | -417 | 0.63 | 0.19 | 1 |
| $L_{vb}$ | 6e+09 ± 2e+09 | 0.13 | 0.03 | 1865 | 0.61 | 0.11 | 1 |
| $L_v$ | 6e+09 ± 3e+09 | 0.12 | 0.02 | 1874 | 0.63 | 0.10 | 1 |
| $mN1$ | 3e-01 ± 1e-01 | 0.17 | 0.01 | 333 | 0.54 | 0.16 | 1 |

[a] Slope ± SE, is the slope and standard error associated with the linear correlation regression; $R^2$, is the coefficient of determination; p-value, is the probability that the predictor (WVS) is related to the response (CCC) compared to the null hypothesis of being unrelated; AICc, is the Akaike information criterion; AICcWt, is the AICc weight; $aR^2$, is the adjusted $R^2$ and PO, is the polynomial order of the best fit curve up to order 9.

### 11. North Toronto (TNT) Longer Term PMMoV-normalized N1 ($C_{vb}$) viral concentration and CCC by Reported Date

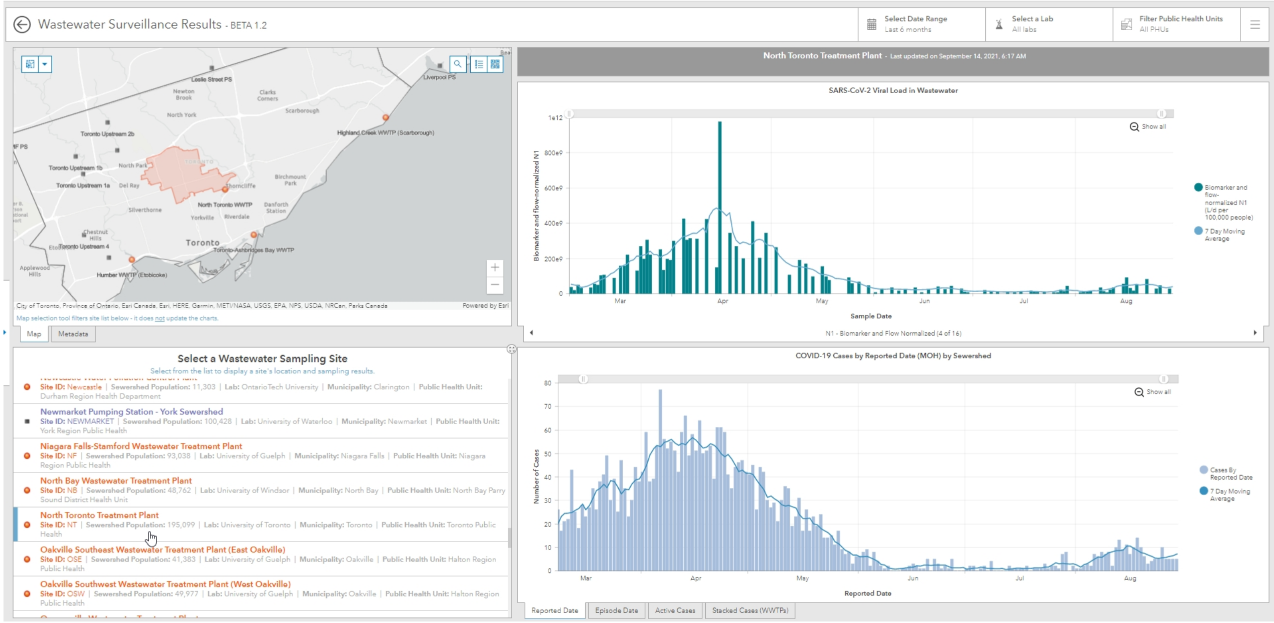


**Figure S13.** North Toronto sewershed with viral load and clincal case data from the Ontario Dashboard

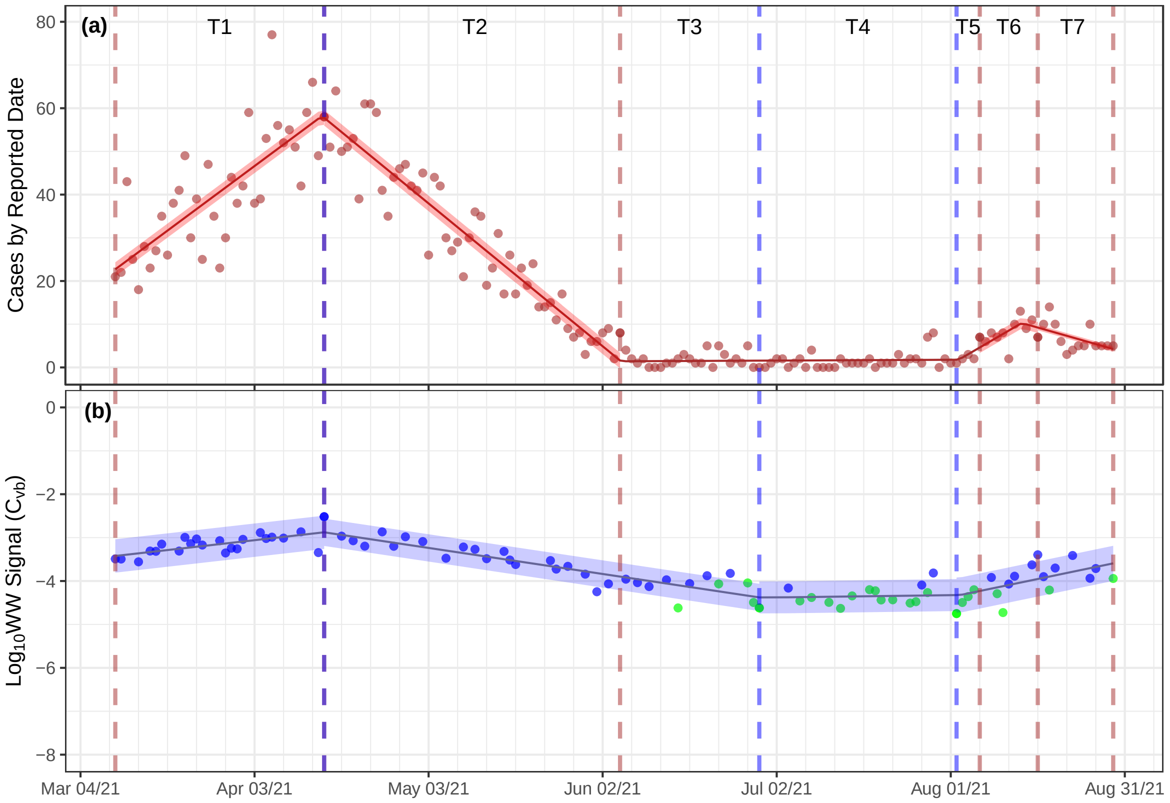


**Figure S14.** The TNT sewershed QTA of CCC by reported date (a) and $\log_{10}$ PMMoV-normalized N1 ($C_{vb}$) viral concentration (b)

**Table S35.** Summary aggregated trend results for the TAB WWTP sewershed[a]

| Interval | Trend Duration | | Trends in CCC | | Trends in WW | |
|---|---|---|---|---|---|---|
| ID | Start | End | DC (CI) | | PDC (CI) | |
| T1 | Mar 10/21 | Apr 15/21 | 1.0 ( 0.8 – 1.2 ) | ↑ | 4 ( 1 – 6 ) | ↑ |
| T2 | Apr 15/21 | Apr 15/21 | -1 ( -1.2 – -1.0 ) | ↓ | 4 ( 1 – 6 ) | ↑ |
| T3 | Apr 15/21 | Jun 05/21 | -1 ( -1.2 – -1.0 ) | ↓ | -4 ( -5 – -4 ) | ↓ |
| T4 | Jun 05/21 | Jun 29/21 | 0.0 ( -0.08 – 0.09 ) | — | -4 ( -5 – -4 ) | ↓ |
| T5 | Jun 29/21 | Aug 06/21 | 0.0 ( -0.08 – 0.09 ) | — | 0.4 ( -2.6 – 3.5 ) | ↑ |
| T6 | Aug 06/21 | Aug 16/21 | 0.8 ( -0.3 – 1.8 ) | ↑ | 0.4 ( -2.6 – 3.5 ) | ↑ |
| T7 | Aug 16/21 | Aug 29/21 | -0.4 ( -1.0 – 0.2 ) | ↓ | 0.4 ( -2.6 – 3.5 ) | ↑ |

[a] DC and PDC, refer to daily change and percent daily change, respectively; CI, is the 95 % confidence interval.

**Table S36.** Summary interpretation of trend results for the TAB WWTP sewershed

| | Signal Trends | | Trends Interpretation |
|---|---|---|---|
| Interval | CCC | WWS | Integration of CCC and WWS Signals |
| T1 | ↑ | ↑ | **Both** the CCC and WWS **are trending up**. Strong evidence of escalating community transmission. |
| T2 | ↓ | ↑ | The CCC is **trending down** and the WWS is **trending up**. Weak evidence of escalating community transmission. |
| T3 | ↓ | ↓ | **Both** the CCC and WWS signals are **trending down**. No evidence of escalating community transmission. |
| T4 | — | ↓ | The CCC is **baseline** and the WWS is **trending down**. No evidence of escalating community transmission. |
| T5 | — | ↑ | The CCC is **baseline** and the WWS is **trending up**. Some evidence of escalating community transmission. |
| T6 | ↑ | ↑ | **Both** the CCC and WWS **are trending up**. Strong evidence of escalating community transmission. |
| T7 | ↓ | ↑ | The CCC is **trending down** and the WWS is **trending up**. Weak evidence of escalating community transmission. |

**Table S37.** The CCC and WWS breakpoints (CBP and WBP) and associated SE[a]

| CCC Breakpoints | | | | WWS Breakpoints | | | |
|---|---|---|---|---|---|---|---|
| ID | Date | CBP | CBP SE | ID | Date | WBP | WBP SE |
| CB1 | 2021-04-15 | 37 | 1 | WB1 | 2021-04-15 | 37 | 4 |
| CB2 | 2021-06-05 | 88 | 2 | WB2 | 2021-06-29 | 112 | 7 |
| CB3 | 2021-08-06 | 150 | 5 | | | | |
| CB4 | 2021-08-16 | 160 | 4 | | | | |

[a] ID, is the code for the CCC and WWS breakpoints; Date, is the break-point date estimated by the segmented routine; CBP and WBP, refer to the CCC and WWS break points, respectively; CBP SE and WBP SE, refer to the SE of the CCC and WWS break points.

**Table S38.** The CCC and WWS trends and associated standard errors (SE)[a]

| CCC Trend Lines | | | | WWS Trend Lines | | | |
|---|---|---|---|---|---|---|---|
| ID | n | DC | DC SE | ID | n | PDC | PDC SE |
| CS1 | 36 | 1.0 | 0.1 | WS1 | 23 | 3.5 | 1.1 |
| CS2 | 51 | -1.1 | 0.1 | WS2 | 31 | -4.5 | 0.4 |
| CS3 | 62 | 0.0 | 0.0 | WS3 | 22 | 0.4 | 1.5 |
| CS4 | 10 | 0.8 | 0.5 | | | | |
| CS5 | 13 | -0.4 | 0.3 | | | | |

[a] ID, is the identification code for the CCC and WWS trend lines; n, is the number of data points within each trend interval; DC, is the daily change or slope in cases per day for the CCC; PDC, is the percent daily change for the WWS or the slope of the trend line within each interval; DC SE and PCD SE, refer to the standard error of the DC and PDC, respectively and correlate to the error of the slope of the linear trend line within each interval.

### 12. TNT Longer Term N1 $L_v$ and CCC by Reported Date

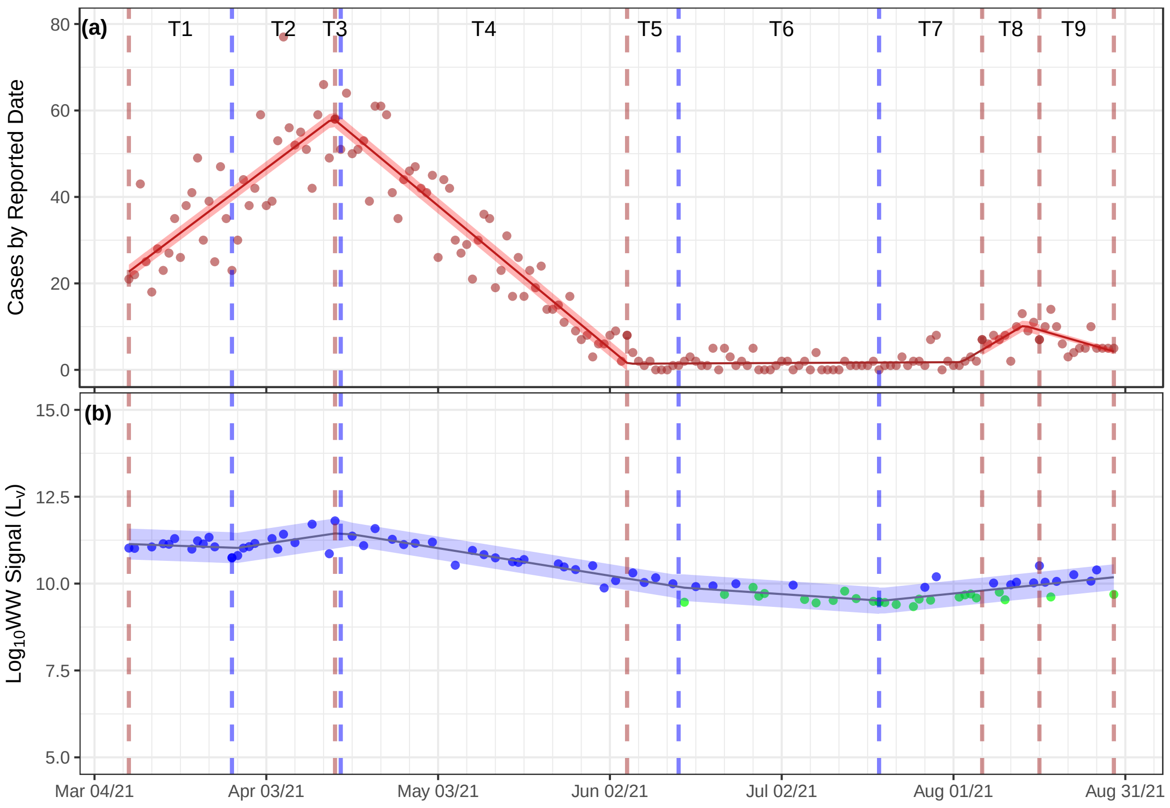


**Figure S15.** The TNT sewershed QTA of CCC by reported date (a) and $\log_{10}$ N1 $L_v$ viral loading (b)

**Table S39.** Summary aggregated trend results for the TAB WWTP sewershed[a]

| Interval | Trend Duration | | Trends in CCC | | Trends in WW | |
|---|---|---|---|---|---|---|
| ID | Start | End | DC (CI) | | PDC (CI) | |
| T1 | Mar 10/21 | Mar 28/21 | 1.0 ( 0.8 – 1.2 ) | ↑ | -1 ( -6 – 4 ) | ↓ |
| T2 | Mar 28/21 | Apr 16/21 | 1.0 ( 0.8 – 1.2 ) | ↑ | 6 ( 0.3 – 12 ) | ↑ |
| T3 | Apr 16/21 | Apr 15/21 | 1.0 ( 0.8 – 1.2 ) | ↑ | -6 ( -7 – -5 ) | ↓ |
| T4 | Apr 15/21 | Jun 05/21 | -1 ( -1.2 – -1.0 ) | ↓ | -6 ( -7 – -5 ) | ↓ |
| T5 | Jun 05/21 | Jun 14/21 | 0.0 ( -0.08 – 0.09 ) | — | -6 ( -7 – -5 ) | ↓ |
| T6 | Jun 14/21 | Jul 19/21 | 0.0 ( -0.08 – 0.09 ) | — | -2 ( -5 – -0 ) | ↓ |
| T7 | Jul 19/21 | Aug 06/21 | 0.0 ( -0.08 – 0.09 ) | — | 4 ( 2 – 6 ) | ↑ |
| T8 | Aug 06/21 | Aug 16/21 | 0.8 ( -0.3 – 1.8 ) | ↑ | 4 ( 2 – 6 ) | ↑ |
| T9 | Aug 16/21 | Aug 29/21 | -0.4 ( -1.0 – 0.2 ) | ↓ | 4 ( 2 – 6 ) | ↑ |

[a] DC and PDC, refer to daily change and percent daily change, respectively; CI, is the 95 % confidence interval.

**Table S40.** Summary interpretation of trend results for the TAB WWTP sewershed

| | Signal Trends | | Trends Interpretation |
|---|---|---|---|
| Interval | CCC | WWS | Integration of CCC and WWS Signals |
| T1 | ⬆ | ⬇ | The CCC **trending up** and the WWS is **trending down**. Weak evidence of escalating community transmission. |
| T2 | ⬆ | ⬆ | **Both** the CCC and WWS **are trending up**. Strong evidence of escalating community transmission. |
| T3 | ⬆ | ⬇ | The CCC **trending up** and the WWS is **trending down**. Weak evidence of escalating community transmission. |
| T4 | ⬇ | ⬇ | **Both** the CCC and WWS signals are **trending down**. No evidence of escalating community transmission. |
| T5 | — | ⬇ | The CCC is **baseline** and the WWS is **trending down**. No evidence of escalating community transmission. |
| T6 | — | ⬇ | The CCC is **baseline** and the WWS is **trending down**. No evidence of escalating community transmission. |
| T7 | — | ⬆ | The CCC is **baseline** and the WWS is **trending up**. Some evidence of escalating community transmission. |
| T8 | ⬆ | ⬆ | **Both** the CCC and WWS **are trending up**. Strong evidence of escalating community transmission. |
| T9 | ⬇ | ⬆ | The CCC is **trending down** and the WWS is **trending up**. Weak evidence of escalating community transmission. |

**Table S41.** The CCC and WWS breakpoints (CBP and WBP) and associated SE[a]

| CCC Breakpoints | | | | WWS Breakpoints | | | |
|---|---|---|---|---|---|---|---|
| ID | Date | CBP | CBP SE | ID | Date | WBP | WBP SE |
| CB1 | 2021-04-15 | 37 | 1 | WB1 | 2021-03-28 | 20 | 6 |
| CB2 | 2021-06-05 | 88 | 2 | WB2 | 2021-04-16 | 38 | 3 |
| CB3 | 2021-08-06 | 150 | 5 | WB3 | 2021-06-14 | 98 | 9 |
| CB4 | 2021-08-16 | 160 | 4 | WB4 | 2021-07-19 | 133 | 5 |

[a] ID, is the code for the CCC and WWS breakpoints; Date, is the break-point date estimated by the segmented routine; CBP and WBP, refer to the CCC and WWS break points, respectively; CBP SE and WBP SE, refer to the SE of the CCC and WWS break points.

**Table S42.** The CCC and WWS trends and associated standard errors (SE)[a]

| CCC Trend Lines | | | | WWS Trend Lines | | | |
|---|---|---|---|---|---|---|---|
| ID | n | DC | DC SE | ID | n | PDC | PDC SE |
| CS1 | 36 | 1.0 | 0.1 | WS1 | 13 | -1.3 | 2.7 |
| CS2 | 51 | -1.1 | 0.1 | WS2 | 11 | 5.8 | 2.7 |
| CS3 | 62 | 0.0 | 0.0 | WS3 | 25 | -5.9 | 0.6 |
| CS4 | 10 | 0.8 | 0.5 | WS4 | 14 | -2.5 | 1.2 |
| CS5 | 13 | -0.4 | 0.3 | WS5 | 27 | 3.9 | 3.9 |

[a] ID, is the identification code for the CCC and WWS trend lines; n, is the number of data points within each trend interval; DC, is the daily change or slope in cases per day for the CCC; PDC, is the percent daily change for the WWS or the slope of the trend line within each interval; DC SE and PCD SE, refer to the standard error of the DC and PDC, respectively and correlate to the error of the slope of the linear trend line within each interval.

### 13. TNT Longer Term $L_{vb}$ with WCR Ratio and CCC by Reported Date

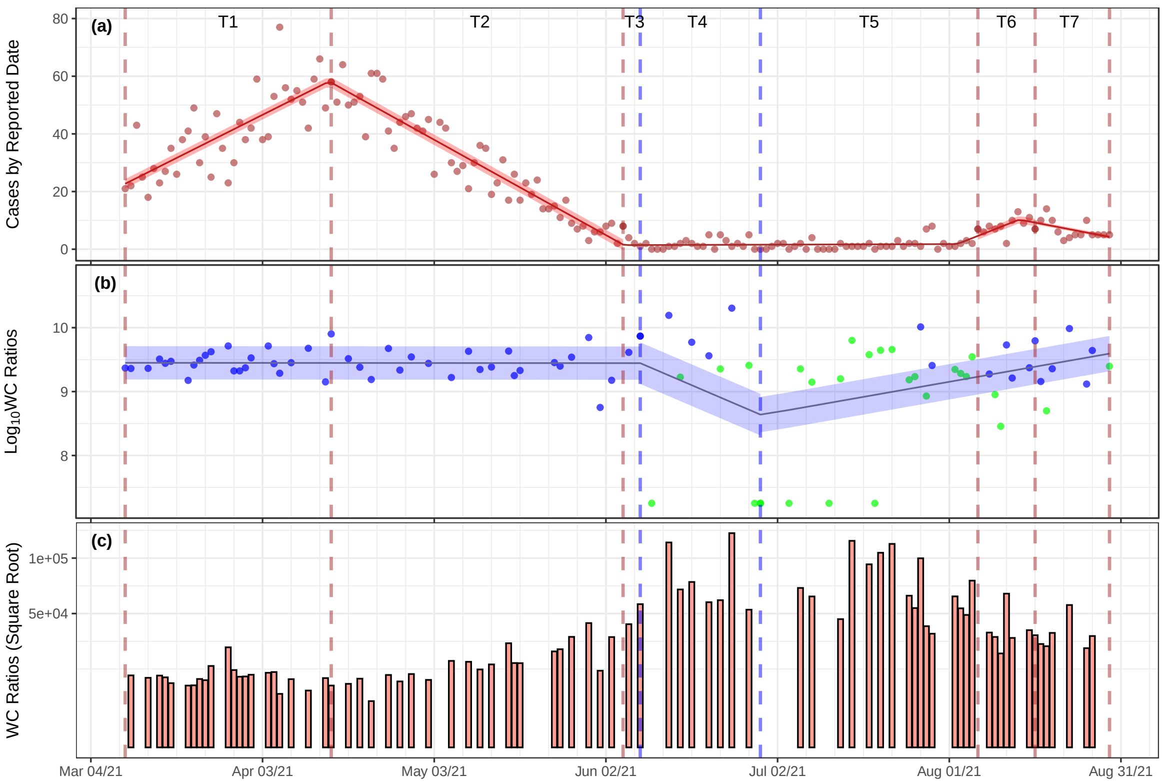


**Figure S16.** The TAB sewershed trend analysis of CCC by reported date (a) and $\log_{10}$ WCR trend analysis (b) and WCR values (c).

**Table S43.** Summary aggregated trend results for the TAB WWTP sewershed

| Interval | Trend Duration | | Trends in CCC | | Trends in WCR | |
|---|---|---|---|---|---|---|
| ID | Start | End | DC (CI) | | PDC (CI) | |
| T1 | Mar 10/21 | Apr 15/21 | 1.0 ( 0.8 – 1.2 ) | ↑ | 0.0 ( -1.5 – 1.5 ) | — |
| T2 | Apr 15/21 | Jun 05/21 | -1 ( -1.2 – -1.0 ) | ↓ | 0.0 ( -1.5 – 1.5 ) | — |
| T3 | Jun 05/21 | Jun 08/21 | 0.0 ( -0.08 – 0.09 ) | — | 0.0 ( -1.5 – 1.5 ) | — |
| T4 | Jun 08/21 | Jun 29/21 | 0.0 ( -0.08 – 0.09 ) | — | -8 ( -20 – 5 ) | ↓ |
| T5 | Jun 29/21 | Aug 06/21 | 0.0 ( -0.08 – 0.09 ) | — | 4 ( 0.6 – 7 ) | ↑ |
| T6 | Aug 06/21 | Aug 16/21 | 0.8 ( -0.3 – 1.8 ) | ↑ | 4 ( 0.6 – 7 ) | ↑ |
| T7 | Aug 16/21 | Aug 29/21 | -0.4 ( -1.0 – 0.2 ) | ↓ | 4 ( 0.6 – 7 ) | ↑ |

**Table S44.** Summary interpretation of trend results for the TAB WWTP sewershed CCC and WCR

| | Signal Trends | | Trends Interpretation |
|---|---|---|---|
| Interval | CCC | WCR | Integration of CCC and WCR |
| T1 | ↑ | — | The CCC is **trending up** and the WCR is **baseline**. Some evidence of escalating community transmission and there is sufficient public health capacity. |
| T2 | ↓ | — | The CCC is **trending down** and the WC Ratio signal is **baseline**. No evidence of escalating community transmission and there is sufficient public health capacity. |
| T3 | — | — | **Both** the CCC and WCR signals are **baseline**. No evidence of escalating community transmission and there is sufficient public health capacity. |
| T4 | — | ↓ | The CCC is **baseline** and the WCR is **trending down**. No evidence of escalating community transmission and clinical testing may be over-estimating disease incidence when counting previously infected cases as new cases. |
| T5 | — | ↑ | The CCC is **baseline** and the WCR is **trending up**. Some evidence of escalating community transmission and increase in asymptomatic cases in the community. |
| T6 | ↑ | ↑ | **Both** the CCC and WCR **are trending up**. Strong evidence of escalating community transmission and increase in asymptomatic cases in the community. |
| T7 | ↓ | ↑ | The CCC is **trending down** and the WCR is **trending up**. Weak evidence of escalating community transmission and increase in asymptomatic cases in the community. |

**Table S45.** The CCC and WWS estimated breakpoints and associated standard errors[a]

| CCC Breakpoints | | | | WCR Breakpoints | | | |
|---|---|---|---|---|---|---|---|
| ID | Date | CBP | CBP SE | ID | Date | WCBP | WCBP SE |
| CB1 | 2021-04-15 | 37 | 1 | WB1 | 2021-06-08 | 91 | 12 |
| CB2 | 2021-06-05 | 88 | 2 | WB2 | 2021-06-29 | 112 | 7 |
| CB3 | 2021-08-06 | 150 | 5 | | | | |
| CB4 | 2021-08-16 | 160 | 4 | | | | |

[a] ID, is the code for the CCC and WC breakpoints; Date, is the breakpoint date estimated by the segmented routine; CBP and WCBP, refer to the CSS and WC breakpoints; CBP SE and WCBP SE, refer to the SE of the CSB and WCBP, within each interval.

**Table S46.** The CCC and WCR estimated trend lines slopes and associated standard errors[a]

| CCC Trend Lines | | | | WCR Trend Lines | | | |
|---|---|---|---|---|---|---|---|
| ID | n | DC | DC SE | ID | n | PDWC | PDWC SE |
| CS1 | 37 | 1.0 | 0.1 | WS1 | 45 | 0.0 | 0.8 |
| CS2 | 51 | -1.1 | 0.1 | WS2 | 9 | -8.4 | 7.1 |
| CS3 | 37 | -1.1 | 0.1 | WS3 | 36 | 3.7 | 1.5 |
| CS4 | 51 | -1.1 | 0.1 | | | | |
| CS5 | 37 | -1.1 | 0.1 | | | | |

[a] ID, is the code for the CCC and WCR trend lines; n, is the number of data points within each interval; DC and DWC, are the daily change in the CSS and percent daily change (PDC) of the WCR of the trend line (or slope) within each interval; DC SE, and PWDC SE, refer to the standard error in the corresponding units of the DC and PDWC.

### 14. TNT Linear Correlation of WWS to CCC by Reported Date

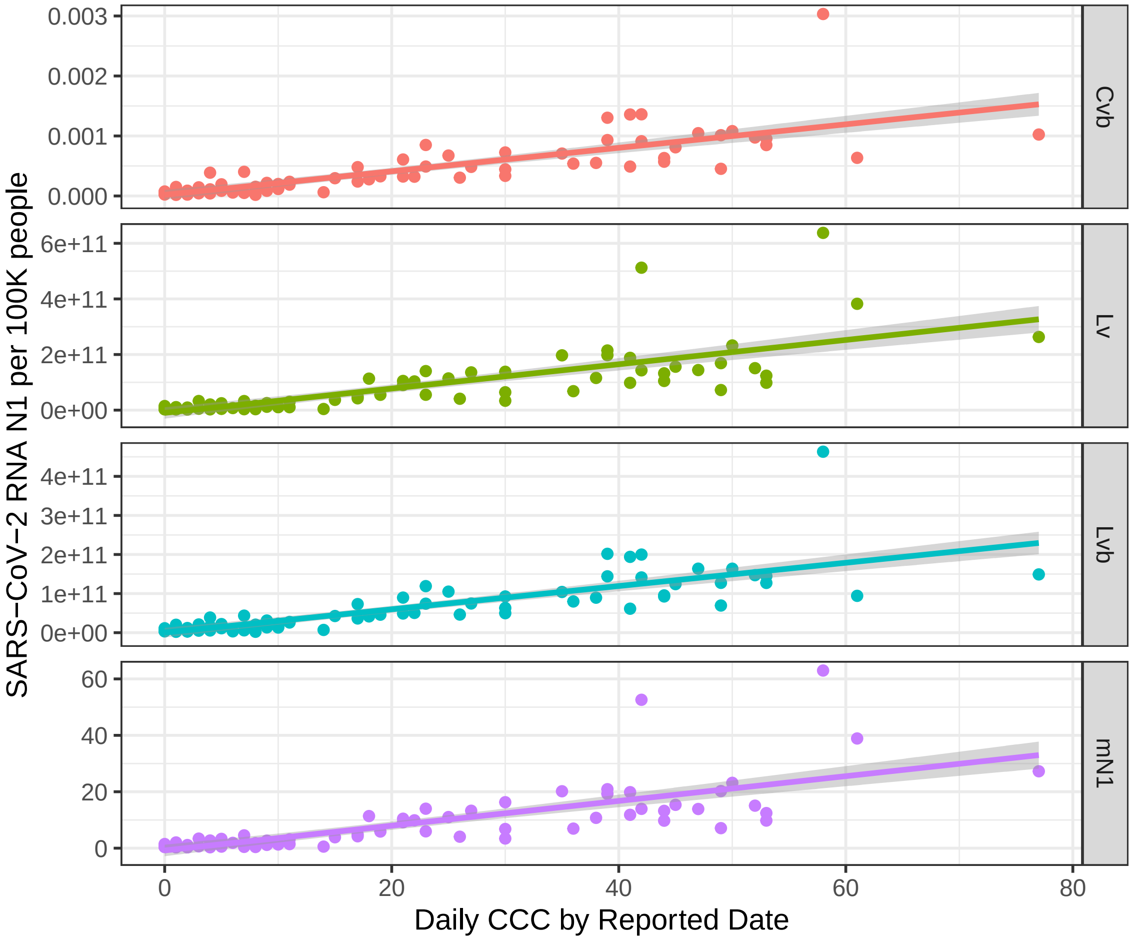


**Figure S17.** The TNT sewershed linear correlation normalizations for N1 ($C_{vb}$, $L_{vb}$), $L_v$ and un-normalized $mN1$) versus clinical case counts (CCC) by reported date with 95% confidence bands

**Table S47.** TNT N1 WWS ($C_{vb}$, $L_v$, $L_{vb}$, mN1) versus the daily CCC Correlation Results

| | Linear Correlation Results | | | Nonlinear Correlation Results | | | |
|---|---|---|---|---|---|---|---|
| Normalization | Slope ± SE | $R^2$ | P-Value | AICc | AICcWt | $aR^2$ | PO |
| $C_{vb}$ | 2e-05 ± 2e-06 | 0.65 | < 0.01 | -1171 | 0.98 | 0.72 | 9 |
| $L_{vb}$ | 3e+09 ± 2e+08 | 0.65 | < 0.01 | 1865 | 1.00 | 0.73 | 9 |
| $L_v$ | 4e+09 ± 4e+08 | 0.59 | < 0.01 | 4528 | 0.79 | 0.69 | 9 |
| $mN1$ | 4e-01 ± 4e-02 | 0.59 | < 0.01 | 333 | 0.65 | 0.68 | 9 |

[a] Slope ± SE, is the slope and standard error associated with the linear correlation regression; $R^2$, is the coefficient of determination; p-value, is the probability that the predictor (WVS) is related to the response (CCC) compared to the null hypothesis of being unrelated; AICc, is the Akaike information criterion; AICcWt, is the AICc weight; $aR^2$, is the adjusted $R^2$ and PO, is the polynomial order of the best fit curve up to order 9.

### 15. Ontario Data Template (ODT)

The Ontario Data Template (ODT) is an MS Excel$^{TM}$ Workbook with eight worksheets designed to capture the essential data and metadata associated with field campaigns for wastewater surveillance for SARS-CoV-2. It was generated from the Open Data Model (ODM) version 1.1 [1] and is used by all participating Laboratories in the Ontario Wastewater Surveillance Initiative (WSI). The three main worksheets with key data fields are described and shown in screen prints below:

1. Part of Worksheet 6, "Sample" is shown in Table S48 along with the tabs of the other seven worksheets that comprise the collected and submitted primary laboratory data and associate metadata. The Sample worksheet is used to report on the: sample identification ('sampleID'); the sampling site typically at the influent to the WWTP ('siteID'); stream of wastewater sample (type); type of sample collected ('collection') (e.g., 24-hour composite, time proportional); start ('dateTimeStart') and end ('dateTimeEnd') of the sample collection; the volume, in liters, of the sample collected ('sizeL'); field sample collection storage during transport ('filedSmapleTempC') typically shipped on ice and temperature given as 4 $^{o}$C; and indication if shipped on ice (typically yes) ('shippedOnIce').
2. Part of Worksheet 7, "WWMeasure" is shown in Table S49. The WWMeasure worksheet is used to report on the: sample identification ('sampleII'); the analysis date ('analysisDate'); the fraction of the sample analyzed ('fractionAnalyzed'); the RNA gene type sampled ('type') representing N1, N2 and PMMoV in this particular case; the unit of the reporte value ('unit'); index of the technical or biological replicates ('index') and the concentration value of the qPCR results ('value').
3. Part of Worksheet 8, "SiteMeasure" is shown in Table S50. The SiteMeasure worksheet is used to report on the: a unique site id associated with the type of field measure conducted ('uSiteMeasureID'); the sampling site typically at the influent to the WWTP ('siteID'); sample identification ('sampleID'); the date-time of the site measurement ('dateTime'); stream of wastewater sample (type); the description of the field measurement taken ('typeDescription'); the type of aggregation associated with the field measurement (e.g., point sample or mean) ('aggregation'); the value of the measurement taken ('value') and the units of measurement ('unit').

Note that some of the paramter fields (e.g., site ID, sampleID) are used as primary or foreign keys and duplicated to facilitate aggregation and data validation prior to data transcription to the Extended Aggregated WSI Data set and making this data available for download.

**Table S48.** Sample screen print of part of Worksheet 6 (Sample) of the Ontario Data Template

| | A | B | C | D | F | G | H | I | J |
|---|---|---|---|---|---|---|---|---|---|
| 1 | sampleID | siteID | type | collection | dateTimeStart | dateTimeEnd | sizeL | fieldSampleTempC | shippedOnIce |
| 521 | hr_2021Jun13_c24 | hr | rawWW | cpTP24h | 2021-06-13 0:05 | 2021-06-14 0:00 | 1.0 | 4.0 | Yes |
| 534 | hr_2021Jun14_c24 | hr | rawWW | cpTP24h | 2021-06-14 0:05 | 2021-06-15 0:00 | 1.0 | 4.0 | Yes |
| 535 | hr_2021Jun15_c24 | hr | rawWW | cpTP24h | 2021-06-15 0:05 | 2021-06-16 0:00 | 1.0 | 4.0 | Yes |
| 536 | hr_2021Jun16_c24 | hr | rawWW | cpTP24h | 2021-06-16 0:05 | 2021-06-17 0:00 | 1.0 | 4.0 | Yes |
| 537 | hr_2021Jun17_c24 | hr | rawWW | cpTP24h | 2021-06-17 0:05 | 2021-06-18 0:00 | 1.0 | 4.0 | Yes |
| 538 | hr_2021Jun20_c24 | hr | rawWW | cpTP24h | 2021-06-20 0:05 | 2021-06-21 0:00 | 1.0 | 4.0 | Yes |
| 551 | hr_2021Jun21_c24 | hr | rawWW | cpTP24h | 2021-06-21 0:05 | 2021-06-22 0:00 | 1.0 | 4.0 | Yes |
| 552 | hr_2021Jun22_c24 | hr | rawWW | cpTP24h | 2021-06-22 0:05 | 2021-06-23 0:00 | 1.0 | 4.0 | Yes |
| 564 | hr_2021Jun23_c24 | hr | rawWW | cpTP24h | 2021-06-23 0:05 | 2021-06-24 0:00 | 1.0 | 4.0 | Yes |
| 565 | hr_2021Jun24_c24 | hr | rawWW | cpTP24h | 2021-06-24 0:05 | 2021-06-25 0:00 | 1.0 | 4.0 | Yes |
| 566 | hr_2021Jun27_c24 | hr | rawWW | cpTP24h | 2021-06-27 0:05 | 2021-06-28 0:00 | 1.0 | 4.0 | Yes |
| 568 | hr_2021Jun28_c24 | hr | rawWW | cpTP24h | 2021-06-28 0:05 | 2021-06-29 0:00 | 1.0 | 4.0 | Yes |
| 569 | hr_2021Jun29_c24 | hr | rawWW | cpTP24h | 2021-06-29 0:05 | 2021-06-30 0:00 | 1.0 | 4.0 | Yes |
| 570 | hr_2021Jun30_c24 | hr | rawWW | cpTP24h | 2021-06-30 0:05 | 2021-07-01 0:00 | 1.0 | 4.0 | Yes |
| 571 | hr_2021Jul01_c24 | hr | rawWW | cpTP24h | 2021-07-01 0:05 | 2021-07-02 0:00 | 1.0 | 4.0 | Yes |
| 572 | hr_2021Jul04_c24 | hr | rawWW | cpTP24h | 2021-07-04 0:05 | 2021-07-05 0:00 | 1.0 | 4.0 | Yes |
| 585 | hr_2021Jul05_c24 | hr | rawWW | cpTP24h | 2021-07-05 0:05 | 2021-07-06 0:00 | 1.0 | 4.0 | Yes |
| 586 | hr_2021Jul06_c24 | hr | rawWW | cpTP24h | 2021-07-06 0:05 | 2021-07-07 0:00 | 1.0 | 4.0 | Yes |
| 592 | hr_2021Jul07_c24 | hr | rawWW | cpTP24h | 2021-07-07 0:05 | 2021-07-08 0:00 | 1.0 | 4.0 | Yes |
| 593 | hr_2021Jul08_c24 | hr | rawWW | cpTP24h | 2021-07-08 0:05 | 2021-07-09 0:00 | 1.0 | 4.0 | Yes |

... | 2 - Reporter | 3 - Lab | 4 - Instrument | 5 - AssayMethod | 6 - Sample | 7 - WWMeasure | 8 - SiteMeasure | Definitio ...

**Table S49.** Sample screen print of part of Worksheet 7 (WWMeasure) of the Ontario Data Template

| | B | G | I | J | L | N | O | R | S |
|---|---|---|---|---|---|---|---|---|---|
| 1 | sampleID | analysisDate | fractionAnalyzed | type | unit | index | value | | |
| 5956 | hr_2021Jul21_c24 | 2021-07-26 | solid | covN1 | gcMl | 1_3 | 3.57 | | |
| 5957 | hr_2021Jul21_c24 | 2021-07-26 | solid | covN2 | gcMl | 1_1 | 2.71 | | |
| 5958 | hr_2021Jul21_c24 | 2021-07-26 | solid | covN2 | gcMl | 1_2 | 4.05 | | |
| 5959 | hr_2021Jul21_c24 | 2021-07-26 | solid | covN2 | gcMl | 1_3 | 2.71 | | |
| 5960 | hr_2021Jul21_c24 | 2021-07-26 | solid | nPMMoV | gcMl | 1_1 | 4261.75 | | |
| 5961 | hr_2021Jul21_c24 | 2021-07-26 | solid | nPMMoV | gcMl | 1_2 | 5644.32 | | |
| 5962 | hr_2021Jul21_c24 | 2021-07-26 | solid | nPMMoV | gcMl | 1_3 | 6807.08 | | |
| 5963 | hr_2021Jul21_c24 | 2021-07-26 | solid | covN1 | gcMl | 2_1 | 2.43 | | |
| 5964 | hr_2021Jul21_c24 | 2021-07-26 | solid | covN1 | gcMl | 2_2 | 2.43 | | |
| 5965 | hr_2021Jul21_c24 | 2021-07-26 | solid | covN1 | gcMl | 2_3 | 2.43 | | |
| 5966 | hr_2021Jul21_c24 | 2021-07-26 | solid | covN2 | gcMl | 2_1 | 2.68 | | |
| 5967 | hr_2021Jul21_c24 | 2021-07-26 | solid | covN2 | gcMl | 2_2 | 2.68 | | |
| 5968 | hr_2021Jul21_c24 | 2021-07-26 | solid | covN2 | gcMl | 2_3 | 2.68 | | |
| 5969 | hr_2021Jul21_c24 | 2021-07-26 | solid | nPMMoV | gcMl | 2_1 | 5508.33 | | |
| 5970 | hr_2021Jul21_c24 | 2021-07-26 | solid | nPMMoV | gcMl | 2_2 | 5398.89 | | |
| 5971 | hr_2021Jul21_c24 | 2021-07-26 | solid | nPMMoV | gcMl | 2_3 | 7008.29 | | |
| 5972 | hr_2021Jul22_c24 | 2021-07-26 | solid | covN1 | gcMl | 1_2 | 3.15 | | |
| 5973 | hr_2021Jul22_c24 | 2021-07-26 | solid | covN1 | gcMl | 1_3 | 2.85 | | |
| 5974 | hr_2021Jul22_c24 | 2021-07-26 | solid | covN2 | gcMl | 1_1 | 2.71 | | |
| 5975 | hr_2021Jul22_c24 | 2021-07-26 | solid | covN2 | gcMl | 1_2 | 2.71 | | |

... | 2 - Reporter | 3 - Lab | 4 - Instrument | 5 - AssayMethod | 6 - Sample | 7 - WWMeasure | 8 - SiteMeasure

**Table S50.** Sample screen print of part of Worksheet 8 of the Ontario Data Template

| 1 | uSiteMeasureID | siteID | sampleID | dateTime | type | typeDescription | aggregation | value | unit |
|---|---|---|---|---|---|---|---|---|---|
| 566 | hr_flow_133 | hr | hr_2021Jun27_c24 | 2021-06-27 0:00 | wwFlow | Flow data from WWTP | mean | 258.8 | ML/D |
| 568 | hr_flow_134 | hr | hr_2021Jun28_c24 | 2021-06-28 0:00 | wwFlow | Flow data from WWTP | mean | 255.7 | ML/D |
| 569 | hr_flow_135 | hr | hr_2021Jun29_c24 | 2021-06-29 0:00 | wwFlow | Flow data from WWTP | mean | 364.9 | ML/D |
| 570 | hr_flow_136 | hr | hr_2021Jun30_c24 | 2021-06-30 0:00 | wwFlow | Flow data from WWTP | mean | 278.2 | ML/D |
| 571 | hr_flow_137 | hr | hr_2021Jul01_c24 | 2021-07-01 0:00 | wwFlow | Flow data from WWTP | mean | 235.0 | ML/D |
| 572 | hr_flow_138 | hr | hr_2021Jul04_c24 | 2021-07-04 0:00 | wwFlow | Flow data from WWTP | mean | 230.2 | ML/D |
| 585 | hr_flow_139 | hr | hr_2021Jul05_c24 | 2021-07-05 0:00 | wwFlow | Flow data from WWTP | mean | 243.7 | ML/D |
| 586 | hr_flow_140 | hr | hr_2021Jul06_c24 | 2021-07-06 0:00 | wwFlow | Flow data from WWTP | mean | 248.2 | ML/D |
| 592 | hr_flow_141 | hr | hr_2021Jul07_c24 | 2021-07-07 0:00 | wwFlow | Flow data from WWTP | mean | 272.5 | ML/D |
| 593 | hr_flow_142 | hr | hr_2021Jul08_c24 | 2021-07-08 0:00 | wwFlow | Flow data from WWTP | mean | 375.6 | ML/D |
| 594 | hr_flow_143 | hr | hr_2021Jul11_c24 | 2021-07-11 0:00 | wwFlow | Flow data from WWTP | mean | 239.6 | ML/D |
| 602 | hr_flow_144 | hr | hr_2021Jul12_c24 | 2021-07-12 0:00 | wwFlow | Flow data from WWTP | mean | 244.4 | ML/D |
| 614 | hr_flow_145 | hr | hr_2021Jul13_c24 | 2021-07-13 0:00 | wwFlow | Flow data from WWTP | mean | 272.2 | ML/D |
| 615 | hr_flow_146 | hr | hr_2021Jul14_c24 | 2021-07-14 0:00 | wwFlow | Flow data from WWTP | mean | 252.9 | ML/D |
| 616 | hr_flow_147 | hr | hr_2021Jul15_c24 | 2021-07-15 0:00 | wwFlow | Flow data from WWTP | mean | 287.3 | ML/D |
| 617 | hr_flow_148 | hr | hr_2021Jul18_c24 | 2021-07-18 0:00 | wwFlow | Flow data from WWTP | mean | 243.1 | ML/D |
| 619 | hr_flow_149 | hr | hr_2021Jul19_c24 | 2021-07-19 0:00 | wwFlow | Flow data from WWTP | mean | 245.1 | ML/D |
| 620 | hr_flow_150 | hr | hr_2021Jul20_c24 | 2021-07-20 0:00 | wwFlow | Flow data from WWTP | mean | 247.2 | ML/D |
| 627 | hr_flow_151 | hr | hr_2021Jul21_c24 | 2021-07-21 0:00 | wwFlow | Flow data from WWTP | mean | 235.6 | ML/D |
| 628 | hr_flow_152 | hr | hr_2021Jul22_c24 | 2021-07-22 0:00 | wwFlow | Flow data from WWTP | mean | 225.5 | ML/D |

... 2 - Reporter | 3 - Lab | 4 - Instrument | 5 - AssayMethod | 6 - Sample | 7 - WWMeasure | 8 - SiteMeasure | Definitio ...

### 16. Sample Extended Aggregated Data Set

An R-script (not described) was used to transcribe the ODT into the Extended Aggregated WSI Data set (EAD) which is made available for download from the Ontario Dashboard (landing page shown in Figure S18). A subset of the EAD is shown in Table S51 which identifies the key fields and sample data structure accessed for the QTA conducted on each sewershed. The EAD is an MS Excel$^{TM}$ csv file and is an aggregation of sewershed properties, the clinical case data from Ontario Ministry of Health and Long Term Care (MOHLTC) and wastewater RNA data along with normalized values ($C_{vb}$, $L_v$, $L_{vb}$, now shown). Some of the key fields included: the sample date when the wastewater field sample was taken ('sampleDate'); the public health unit where the sampled sewershed is located ('sys_PHU'); the site of the WWTP influent location where the sample is taken and which serves the sewershed ('siteName'); the sample identification code ('sampleID'); the type of sample collected (e.g., grab, composite, time or flow proportional 24-hour composite) ('sampleCollection'); cases by episode date, sewershed specific data collected through a direct link from MOH ('moh_cbed'); field flow measurement ('mQ'); units of the flow measurement ('flowUnit'); the geometric mean concentration of the gene RNA primers N1 ('mN1), N2 ('mN2), geometric mean of N1 and N2 (N1N2) ('mN1N2'); the biomarker PMMoV concentration value ('mBiomarker'); biomarker type ('biomarkerType') and consistent concentration units for both the gene primers and biomarker ('geneUnit').

The EAD is available for download from the Ontario Dashboard by selecting the 'DATA Extended Aggregated WSI' image shown in Figure S18 which is the landing page and launch pad to access other related products.

**Table S51.** Sample screen print of part of the EAD downloadable from the Ontario Dashboard

| | A | B | I | R | S | T | X | Y | Z | AA | AB | AD | AE | AF |
|---|---|---|---|---|---|---|---|---|---|---|---|---|---|---|
| 1 | sampleDate | sys_PHU | siteName | sampleID | sample_collection | moh_cbed | mQ | flowUnit | mN1 | mN2 | mN1N2 | mBiomarker | biomarkerType | geneUnit |
| 87 | 05/07/2021 | Toronto Public Health | Ashbridge Treatment Plant | | | 275 | | | | | | | | |
| 88 | 05/08/2021 | Toronto Public Health | Ashbridge Treatment Plant | | | 242 | | | | | | | | |
| 89 | 05/09/2021 | Toronto Public Health | Ashbridge Treatment Plant | TAB-May09 | cpTP24h | 238 | 441 | ML/d | 17 | 14 | 15 | 16004 | PMMoV | gc/mL |
| 90 | 05/10/2021 | Toronto Public Health | Ashbridge Treatment Plant | | | 283 | | | | | | | | |
| 91 | 05/11/2021 | Toronto Public Health | Ashbridge Treatment Plant | TAB-May11 | cpTP24h | 212 | 461 | ML/d | 43 | 28 | 35 | 16943 | PMMoV | gc/mL |
| 92 | 05/12/2021 | Toronto Public Health | Ashbridge Treatment Plant | | | 221 | | | | | | | | |
| 93 | 05/13/2021 | Toronto Public Health | Ashbridge Treatment Plant | TAB-May13 | cpTP24h | 204 | 443 | ML/d | 22 | 18 | 20 | 21213 | PMMoV | gc/mL |
| 94 | 05/14/2021 | Toronto Public Health | Ashbridge Treatment Plant | | | 208 | | | | | | | | |
| 95 | 05/15/2021 | Toronto Public Health | Ashbridge Treatment Plant | | | 184 | | | | | | | | |
| 96 | 05/16/2021 | Toronto Public Health | Ashbridge Treatment Plant | TAB-May16 | cpTP24h | 152 | 441 | ML/d | 12 | 5 | 8 | 15021 | PMMoV | gc/mL |
| 97 | 05/17/2021 | Toronto Public Health | Ashbridge Treatment Plant | | | 169 | | | | | | | | |
| 98 | 05/18/2021 | Toronto Public Health | Ashbridge Treatment Plant | TAB-May18 | cpTP24h | 185 | 446 | ML/d | 18 | 21 | 19 | 22227 | PMMoV | gc/mL |
| 99 | 05/19/2021 | Toronto Public Health | Ashbridge Treatment Plant | TAB-May19 | cpTP24h | 151 | 460 | ML/d | 12 | 10 | 11 | 18834 | PMMoV | gc/mL |
| 100 | 05/20/2021 | Toronto Public Health | Ashbridge Treatment Plant | | | 130 | | | | | | | | |
| 101 | 05/21/2021 | Toronto Public Health | Ashbridge Treatment Plant | | | 138 | | | | | | | | |
| 102 | 05/22/2021 | Toronto Public Health | Ashbridge Treatment Plant | | | 80 | | | | | | | | |
| 103 | 05/23/2021 | Toronto Public Health | Ashbridge Treatment Plant | | | 91 | | | | | | | | |
| 104 | 05/24/2021 | Toronto Public Health | Ashbridge Treatment Plant | TAB-May24 | cpTP24h | 98 | 432 | ML/d | 11 | 7 | 9 | 20851 | PMMoV | gc/mL |
| 105 | 05/25/2021 | Toronto Public Health | Ashbridge Treatment Plant | TAB-May25 | cpTP24h | 86 | 436 | ML/d | 10 | 8 | 9 | 15253 | PMMoV | gc/mL |
| 106 | 05/26/2021 | Toronto Public Health | Ashbridge Treatment Plant | | | 92 | | | | | | | | |
| 107 | 05/27/2021 | Toronto Public Health | Ashbridge Treatment Plant | TAB-May27 | cpTP24h | 53 | 461 | ML/d | 7 | 6 | 7 | 21296 | PMMoV | gc/mL |
| 108 | 05/28/2021 | Toronto Public Health | Ashbridge Treatment Plant | | | 75 | | | | | | | | |
| 109 | 05/29/2021 | Toronto Public Health | Ashbridge Treatment Plant | | | 50 | | | | | | | | |
| 110 | 05/30/2021 | Toronto Public Health | Ashbridge Treatment Plant | TAB-May30 | cpTP24h | 39 | 466 | ML/d | 10 | 8 | 9 | 28931 | PMMoV | gc/mL |
| 111 | 05/31/2021 | Toronto Public Health | Ashbridge Treatment Plant | | | 53 | | | | | | | | |
| 112 | 06/01/2021 | Toronto Public Health | Ashbridge Treatment Plant | TAB-June01 | cpTP24h | 70 | 455 | ML/d | 5 | 4 | 4 | 17762 | PMMoV | gc/mL |
| 113 | 06/02/2021 | Toronto Public Health | Ashbridge Treatment Plant | | | 51 | | | | | | | | |
| 114 | 06/03/2021 | Toronto Public Health | Ashbridge Treatment Plant | TAB-Jun3 | cpTP24h | 45 | 553 | ML/d | 7 | 5 | 6 | 27966 | PMMoV | gc/mL |
| 115 | 06/04/2021 | Toronto Public Health | Ashbridge Treatment Plant | | | 34 | | | | | | | | |
| 116 | 06/05/2021 | Toronto Public Health | Ashbridge Treatment Plant | | | 36 | | | | | | | | |
| 117 | 06/06/2021 | Toronto Public Health | Ashbridge Treatment Plant | TAB-Jun6 | cpTP24h | 19 | 439 | ML/d | 4 | 1 | 2 | 16643 | PMMoV | gc/mL |
| 118 | 06/07/2021 | Toronto Public Health | Ashbridge Treatment Plant | | | 32 | | | | | | | | |
| 119 | 06/08/2021 | Toronto Public Health | Ashbridge Treatment Plant | TAB-Jun8 | cpTP24h | 41 | 575 | ML/d | 5 | 3 | 4 | 27081 | PMMoV | gc/mL |
| 120 | 06/09/2021 | Toronto Public Health | Ashbridge Treatment Plant | | | 21 | | | | | | | | |
| 121 | 06/10/2021 | Toronto Public Health | Ashbridge Treatment Plant | TAB-Jun10 | cpTP24h | 23 | 486 | ML/d | 4 | 1 | 2 | 48171 | PMMoV | gc/mL |

Toronto_WWTP_Extended_Aggregate

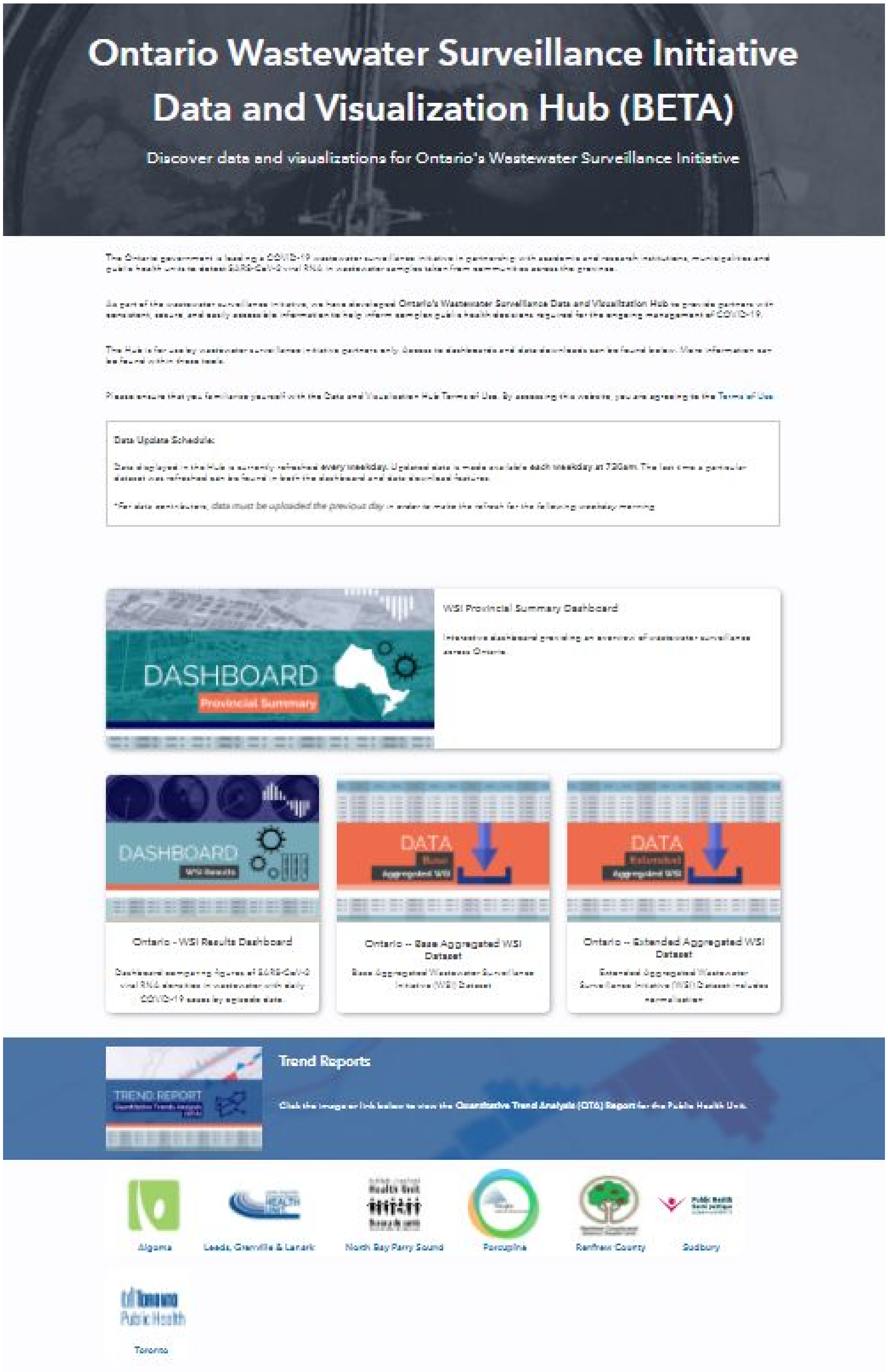


**Figure S18.** Sample screen print of the Ontario Dashboard landing page

### 17. Annotated Sample R-Code

The sample R-code includes the four key functions called by other generic R functions to display the trend lines and extract the essential statistics tabulated and compared in the summary trend results and interpretation of trend results tables:

1. Function 1 determines the seed inflection points in the log-transformed wastewater data ($C_{vb}$. See QTA method flowchart steps (iii) – (v) on how the end user may select the polynomial order to improve the trend fit. A corresponding similar function (not shown) is used to determine the inflection points of cases on a linear scale.
2. Function 2 extracts the pertinent information related to the wastewater signal trend analysis and the output is used to generate the wastewater trend figures as part of the input to Function 3. A corresponding similar function (not shown) is used to extract the pertinent information related to cases on a linear scale.
3. Function 3 generates figure of the *wastewater signal trend analysis* using a log vertical scale versus the trend period. The time intervals signifying a constant trend are clearly delineated and identified by vertical dashed blue lines.
4. Function 4 generates figure of *cases signal trend analysis* using linear vertical scale versus the same trend period as with the wastewater signal. The time intervals signifying a constant trend are clearly delineated and identified by vertical dashed red lines. The figures generated by Function 3 and 4 are aligned for ease of comparison.

```
################################################################################
# Function 1: Used to determine seed log-transformed inflection points with AIC
# ##############################################################################
# Input:
# i. df (default df_to data file) includes normalized WWS or WCR of interest
# ii. NN (default "Cvb") PMMoV wastewater normalized WWS or WCR
# iii. lod (default "loc_Cvb") PMMoV wastewater normalized LOD for WWS or WCR
# Output:
# i. inf_x_pts is the vector of seed inflection points
################################################################################
find_log_infl_points_AIC = function(df = df_to, NN = "Cvb", lod="lod_Cvb")
{

  LODv = as.numeric(unlist(df[,lod]))      # LOD values based on reported LOD
  yv = as.numeric(unlist(df[,NN]))         # Cvb, Lv or Lvb values

  minLODv = min(LODv/2, na.rm = TRUE)      # find minLOD
  minY = min(yv/2, na.rm = TRUE)           #      minY
  minUJ = min(minLODv, minY)               # min of minLOD and minY ~ minUJ

  LOD = log10(df[,lod])
  y = log10(as.numeric(unlist(df[,NN])))    # direct values
  y = ifelse(is.infinite(y),minUJ,          # check for infinite and NAN values
             ifelse(is.nan(y), minUJ, y))
  x = seq(1, max(length(y)), 1)
  lin.mod <- lm(y ~ x)

  op = 9; infl_x_pts = list(); smf = list(); sum_smf = list(); pm=list()
  for(i in 2:op)
  {
    pm[[i]] <- lm(y ~ poly(x, i), data=df2)
    xl <- seq(min(x),max(x), (max(x) - min(x))/1000)
    out = predict(pm[[i]], newdata = data.frame(x=xl))
    infl <- c(FALSE, diff(diff(out)>0)!=0)
    infl_x_pts[[i]] = xl[infl]
    smf[[i]] <- invisible(tryCatch(segmented(lin.mod, seg.Z = ~x, psi=infl_x_pts[[i]]),
                                   error = function(e) e, warning = function(w) w))
    sum_smf[[i]] <- summary(smf[[i]])[6]
  }

  rse = c();  ind = c(); AICf = c()
  for(i in 1:length(sum_smf))
  {
    rse[i] = as.numeric(unlist(sum_smf[[i]]))
    AICf[i] = invisible(tryCatch( AIC(pm[[i]]),
                                  error = function(e) e, warning = function(w) w) )
  }

  aic_min = which.min(AICf)          #(opr = which.min(rse))
  infl_pts = infl_x_pts[[aic_min]]   #(infl_pts = infl_x_pts[[opr]])
  return(infl_pts)
}
```

```r
################################################################################
# Function 2. Used to determine break points and trend lines in log WVS
################################################################################
# Input:
# i. df (default df2_tab data file) that includes normalized load of
#    interest (Cbv, Lb, Lbv) or related WCR
# ii. gene (default "Lb") PMMoV wastewater normalized loading
# iii. infl_pts (default ip_to) seed inflection points
# iv. lod (default "lod_Cvb") corresponding WWS or WCR LOD values
# Output:
# i. wws is a list of WWS or WCR related data to Lb used for trend
#   plots and subsequent analysis: a. wwDS (dft) is the WW data file input
#   data and fitted data b. wwSS (S) is a vector of slopes of trend lines
#   c. ssSD (sdS) is a vector of standard deviations related to slopes
#   d. wwDD (allDates) is a vector of break-point dates of major transitions
################################################################################
ww_log_data_lt =  function(df = df2_tab, gene = "Cvb", infl_pts = ip_to, lod ="lod_Cvb")
{
  LODv = as.numeric(unlist(df[,lod])); yv = as.numeric(unlist(df[,gene]))
  minLODv = min(LODv/2, na.rm = TRUE); minY = min(yv/2, na.rm = TRUE)
  minUJ = log10(min(minLODv, minY));   LOD = log10(df[,lod])
  y = log10(as.numeric(unlist(df[,gene])))
  y = ifelse(is.infinite(y),minUJ, ifelse(is.nan(y), minUJ, y))
  x = 1:max(length(y)); lin.mod <- lm(y ~x)
  sm1 <- segmented(lin.mod, seg.Z = ~x, psi=infl_pts)
  id = attr(na.omit(y), "na.action"); fitted = rep(NA, length(y))
  if(is.null(id)) { fitted = sm1$fitted.values}else{ fitted[-id] = sm1$fitted.values }

  # summary of model results
  t2_summ = summary(sm1) ;  wbp = confint(sm1, digits = 2)
  lpsi = nrow(confint(sm1, digits = 2))  # 95% CI of breakpoints
  wbp = c(0, wbp[1:lpsi], max(x));   nwbp = diff(wbp)
  slopes = segmented::slope(sm1); lni = nrow(slopes$x);   Sl = slopes$x[1:lni,1]
  PDC = (10^Sl-1)*100;   eS = slopes$x[1:lni, 2]
  PDE = (10^eS-1)*100;   lciS = slopes$x[1:lni, 4]; uciS = slopes$x[1:lni, 5]
  Br = confint(sm1)[1:lni-1, 1];   eB = t2_summ$psi[1:lni-1, 3]
  lciB = confint(sm1)[1:lni-1, 2];  uciB = confint(sm1)[1:lni-1, 3]

  startDate = df$sampleDate[1];  br_date = df$sampleDate[Br]
  endDate = df$sampleDate[length(df$sampleDate)];   end_day = length(df$sampleDate)
  allDates = c(startDate, br_date, endDate);  nallDates = length(allDates)
  no_dp = c();   (no_dp[1] = sum(!is.na(y[wbp[1]:wbp[2]])))
  nbpts = length(wbp)

  for(i in 2:(nbpts-1)){ no_dp[i] = sum(!is.na(y[wbp[i]:wbp[i+1]]))  }

  NDP = no_dp;  dft = tibble(sDate = df$sampleDate, x, y, my = fitted, LOD)
  dft <- dft %>% mutate(cy = "blue", cmy ="blue", clod ="green")
  wws= list(wwDS=dft, wwSS=Sl, wwSE = eS, wwSLCI = lciS, wwSUCI = uciS,
          wwDD=allDates,wwBP = Br, wwBE = eB, wwBLCI = lciB, wwBUCI = uciB,
          wwN = NDP, wwSPDC = PDC, wwSPDE = PDE)
  return(wws)
}
```

```r
################################################################################
# Function 3. Used to generate figure of wastewater trend analysis
################################################################################
# Input:
# i. dft (default dwt_to data file) that includes wastewater data
# ii. dcf (default dcf_to data file) that includes case data
# iii. gene (default "Cvb") PMMoV wastewater normalized concentration.
# iv. lod (default "lod_Cvb) LODs to color-code LOD values on figures
# v. mV (dfault mV=c(0,1,0,1)) figure margins adjusted to aggregate figures
# Output:
# i. wws_lnp is a ggplot of trends in (gene load units) versus time
#    (length of data) of trend period which can be selected by the user.
################################################################################
ww_log_plot_lt= function(dwt = dwt_tab, dcf= dcf_tab,
                          gene="Cvb", lod="lod_Cvb", mV=c(0,1,0,1))
{
  yr= c(floor(range(dwt$wwDS$my, na.rm = T)[1] ),
         ceiling(range(dwt$wwDS$my, na.rm = T)[2] ) )
  dww = dwt$wwDS; wwSlopes = dwt$wwSS; wwseSlopes = dwt$wwSE
  wwDates = dwt$wwDD; lwwd = length(wwDates); ltint = length(dwt$wwSS)
  SCI = log10(wwseSlopes)

  if(yr[1] <=0){
    (yr=c(round( floor(yr[1] - abs(min(SCI) + 0.5)),0),
          round(ceiling(yr[2]+abs(max(SCI) + 0.5)),0) ) ) }else{
    (yr=c(round(floor(yr[1]- abs(min(SCI) - 0.5)),0),
          round(ceiling(yr[2]+abs(max(SCI) - 0.5)),0) ) )  }

  ccDates = dcf$ccDD; ndww = list(); bicbs = list()
  ndww[[1]] <- dww %>% filter(sDate >= wwDates[1] & sDate <= wwDates[2])
  for(i in 2:(lwwd-1)){ ndww[[i]] <- dww %>%
      filter(sDate >= wwDates[i] & sDate <= wwDates[i+1]) }

  tdww = list()
  for(i in 1:ltint){
    tdww[[i]] <- ndww[[i]] %>%
      mutate(uci = my + SCI[i]/sqrt(nrow(ndww[[i]])),
             lci = my - SCI[i]/sqrt(nrow(ndww[[i]])), group = i) }

  ttdww = data.table::rbindlist(tdww)
  if(gene == "Cvb"){ labely = expression(paste("", Log[10],
                      "WW Signal"," (",C[vb], ")")) lod_sym = names(dww[,lod])
  }else if(gene == "Lv"){ labely = expression(paste("", Log[10],
                      "WW Signal", " (",L[v], ")")) lod_sym = names(dww[,lod])
  }else if(gene == "Lvb"){labely = expression(paste("", Log[10],
                      "WW Signal", " (",L[vb], ")")) lod_sym = names(dww[,lod])  }

   wws_lnp =  ggplot(ttdww[!is.na(ttdww$my),], aes(x=sDate, y=y, color = cy ) )+
    geom_point(aes(x=sDate, y = y, colour = y <= !!sym(lod_sym)), alpha=0.7)   +
    scale_colour_manual(values = setNames(c("green", "blue"), c(T,F))) +
    geom_line(aes(x=sDate, y=my)) +
    geom_ribbon(aes(x=sDate, ymin=lci, ymax=uci), linetype=0, fill="blue", alpha=0.2) +
    scale_x_date(date_labels="%b %d/%y", breaks="30 days", minor_breaks = "5 days",
                 limits = as.Date(c(dww$sDate[1], dww$sDate[length(dww$sDate)]))) +
```

```
    theme_bw()  + theme(legend.position = "none") +
    ylim(yr[1],yr[2]) +  xlab("") + ylab(labely) +
    theme(plot.title = element_text(hjust = 0, size = 11)) +
    theme(plot.margin=margin(t=mV[1],r=mV[2], b=mV[3], l=mV[4], "cm")) +
    geom_vline(xintercept=ccDates, color ="brown", size =1, alpha=0.5,
               linetype="dashed") + geom_vline(xintercept=wwDates[2:(lwwd-1)],
               color ="blue", size =1, alpha=0.5, linetype="dashed")
  return(wws_lnp)
}


###############################################################################
# Function 4. Used to generate figure of cases trend analysis (linear scale)
###############################################################################
# Input:
# i.   dfc (default dcf_to data file) that includes case data
# ii.  dft (default dwt_to data file) that includes wastewater data
# iii.  mV (default c(0,1,0,1) margins that allow figure alignment
# iv. chow (default "cbrdp" ) used to distinguish cases by reported data and
#          episode date (cbedp)
# Output:
# i. agplot, is a ggplot of combined WWS and CCC trends in
#    (gene units)versus time (data len) with intervals (T1 up to T8), clear
#    lines showing change in trend.
###############################################################################
cases_linear_plot_lt= function(dcf = dcf_tab, dwt = dwt_tab,
                                  mV=mV,  chow = "cbrdp")
{
  yr =c(0, 1.4*floor(range(dcf$ccDS$my, na.rm=T))[2])
  ylt = signif(floor(0.99*yr[2]),2)
  dcf2 = dcf$ccDS ;  ccSlopes = dcf$ccSS; ccDate = dcf$ccDD
  DCI = dcf$ccBPSE; wwDate = dwt$wwDD; lwwd = length(wwDate) -2
  allDates = sort(unique(c(ccDate, wwDate)))); lad = length(allDates)
  intLen = difftime(allDates[2:lad], allDates[1:(lad-1)], units="days")
  tx = ymd(allDates[1:(lad-1)]) + round(intLen/2, 0)); ltx = length(tx)
  lccd = length(ccDate)

  laba = switch(ltx-1,  c("T1", "T2"), c("T1", "T2", "T3"),
    c("T1", "T2", "T3", "T4"),  c("T1", "T2", "T3", "T4", "T5"),
    c("T1", "T2", "T3", "T4", "T5", "T6"),
    c("T1", "T2", "T3", "T4", "T5", "T6", "T7"),
    c("T1", "T2", "T3", "T4", "T5", "T6", "T7", "T8"),
    c("T1", "T2", "T3", "T4", "T5", "T6", "T7", "T8", "T9"),
    c("T1", "T2", "T3", "T4", "T5", "T6", "T7", "T8", "T9", "T10"),
    c("T1", "T2", "T3", "T4", "T5", "T6", "T7", "T8", "T9", "T10", "T11"),
    c("T1", "T2", "T3", "T4", "T5", "T6", "T7", "T8", "T9", "T10", "T11", "T12")
  )

  if(chow == "cbrdp"){yclab = "Cases by Reported Date"}
  else{yclab = "Cases by Episode Date"}
  ndcc = list()
  ndcc[[1]] <- dcf2 %>% filter(sDate >= ccDate[1] & sDate <= ccDate[2])
  for(i in 2:(lccd-1)){ ndcc[[i]] <- dcf2%>%
      filter(sDate >= ccDate[i] & sDate <= ccDate[i+1])  }
```

```
  tint = diff.Date(ccDate)); ltint = length(tint)
  CC_LCI = dcf$ccSLCI; CC_UCI = dcf$ccSUCI
  tdcc = list()
  for(i in 1:ltint){
    tdcc[[i]] <- ndcc[[i]] %>%
      mutate(uci = my + 4*CC_UCI[i], lci = my - 4*CC_LCI[i], group = i)
  }
  ttdcc = data.table::rbindlist(tdcc)

  agplot = ggplot(ttdcc,aes(x=sDate, y=y, color = cy) )+
    geom_point(aes(x=sDate, y=y, color = cy), alpha=0.6) +
    geom_line(aes(sDate,my)) +
    geom_ribbon(aes(x=sDate, ymin=lci, ymax=uci), linetype=0, fill="red", alpha=0.3)  +
    scale_x_date(date_labels="%b %d/%y", breaks="30 days", minor_breaks = "5 days",
                 limits = as.Date(c(dcf2$sDate[1], dcf2$sDate[length(dcf2$sDate)]))) +
    theme_bw() + scale_color_identity() +
    xlab("") + ylab(yclab)+
    theme(plot.title = element_text(hjust = 0, size = 11)) +
    theme(plot.margin=margin(t=mV[1],r=mV[2], b=mV[3], l=mV[4], "cm")) +
    geom_vline(xintercept=ccDate,
               color ="brown", size =1, alpha=0.5, linetype="dashed") +
    geom_vline(xintercept=wwDate[2:(lwwd+1)],
               color ="blue", size =1, alpha=0.5, linetype="dashed") +
    theme(axis.title.x=element_blank(), axis.text.x=element_blank()) +
    annotate("text", x = tx, y = ylt, label = laba) +
    coord_cartesian(ylim= c(yr[1],yr[2]), clip ="off")

  return(agplot)
}
```

### 18. References


[1] National Contributors on GitHub Website. The public health environmental surveillance open data model (PHES-ODM or ODM), v. 2.0. ODM V2.0; 2021.

[2] Water Research Foundation. Wastewater Surveillance of the COVID-19 Genetic Signal in Sewersheds Recommendations from Global Experts. WRT Summit Handout V3b; 2020.

[3] Naughton CC, Roman FA, Alvarado AGF, Tariqi AQ, Deeming MA, Bibby K, et al. Show us the Data: Global COVID-19 Wastewater Monitoring Efforts, Equity, and Gaps. medRxiv 2021. doi:10.1101/2021.03.14.21253564.

[4] Arabzadeh G R., Rauch W. Data filtering methods for SARS-CoV-2 wastewater surveillance. medRxiv 2021;e-print on arXiv, Version v1, arXiv2104.11566v1.

[11] US Centers for Disease Control and Prevention. National Wastewater Surveillance System (NWSS). US CDC; 2021.

[12] Shurgold, J., Pang, L., Wijayasri, S., Flemming, C. A., Simhon, A., Fletcher, T., Pileggi, V., Thomas, J., Stuart, R., Glier, M., Brooks, J., Mounchili, A. Outline of Wastewater Surveillance Data Interpretation (Draft Guide). Canada: Public Health Agency of Canada (Lead), University of Alberta, Ontario Ministry of the Environment, Conservation; Parks, Toronto Public Health, British Columbia CDC; 2021.

[13] Public Health Agency of Canada. Draft – SARS-CoV-2 wastewater-based surveillance: A guide for public health action. Ottawa, Canada: PHAC; 2021.

[14] Muggeo VMR. Interval estimation for the breakpoint in segmented regression: A smoothed score-based approach. Australian & New Zealand Journal of Statistics 2017;59:311–22. doi:doi:10.1111/anzs.12200.

[15] Ontario MECP. Transfer Payment Agreements (Internal MECP Agreements). 2021.

[16] Ontario MECP. MECP Wastewater Surveillances Best Practices Technical Guide (MECP Draft Internal Guide. 2021.

[17] Nicolai N., Therrien J.-D., Maere. T, Pileggi V., Swerdfeger H., Vanrolleghem P.A., Manuel D. Open Data Model for collecting, quality-ensuring and sharing of SARS-CoV-2 data and metadata, EU4S Sewage Sentinel System for SARS CoV-2 - 5th Town Hall Meeting, e-poster. ODM Reference; 2021.

[18] Ontario MECP, Ontario Clean Water Agency. MECP Wastewater Surveillance Initiative, Protocol for Evaluations of RT-qPCR Performance Characteristics (ISBN 978-1-4868-5481-3 Internal Laboratory Protocol). 2021.

[19] Toronto Water. Ashbridges Bay Treatment Plant 2020 Annual Report. TAB Report; 2021.

[20] Toronto Water. Humber River Treatment Plant 2020 Annual Report. THR Report; 2021.

[21] Toronto Water. Highland Creek Treatment Plant 2020 Annual Report. THC Report; 2021.

[22] Toronto Water. North Toronto Treatment Plant 2020 Annual Report. TNT Report; 2021.

[23] Wickham H. ggplot2: Elegant graphics for data analysis. ggplot2; Springer-Verlag New York; 2016.

[24] Wickham H, Averick M, Bryan J, Chang W, McGowan LD, François R, et al. Welcome to the tidyverse. Journal of Open Source Software 2019;4:1686. doi:10.21105/joss.01686.

[25] Grolemund G, Wickham H. Dates and times made easy with lubridate. Journal of Statistical Software 2011;40:1–25.

[26] Wickham H. Reshaping data with the reshape package. Journal of Statistical Software 2007;21:1–20.

[27] Wickham H, Seidel D. Scales: Scale functions for visualization. scales; 2020.

[28] Auguie B. gridExtra: Miscellaneous functions for "grid" graphics. 2017.

[29] Wilke CO. Cowplot: Streamlined plot theme and plot annotations for 'ggplot2'. 2020.

[30] Harris K. Kowr: Utility functions. 2021.

[31] R Core Team. R: A language and environment for statistical computing. Vienna, Austria: R Foundation for Statistical Computing; R Core Team; 2020.

[32] Zhu H. kableExtra: Construct complex table with 'kable' and pipe syntax. kableExtra; 2021.

[33] Xie Y. Bookdown: Authoring books and technical documents with R markdown. Boca Raton, Florida: bookdown; Chapman; Hall/CRC; 2016.

[34] RStudio Team. RStudio: Integrated development environment for r. Boston, MA: RStudio, PBC; RStudio2021; 2021.

[35] Xie Y. Knitr: A general-purpose package for dynamic report generation in r. 2021.

[36] Xie Y. Bookdown: Authoring books and technical documents with r markdown. 2021.

[37] Lamport L. LaTeX: A document preparation system. United States: Addison-Wesley; 1994.

[38] Anna Krystalli. Reproducible research data and project management in r. Reproducible Research; 2021.
